# The Role of X Chromosome in Alzheimer’s Disease Genetics

**DOI:** 10.1101/2024.04.22.24306094

**Authors:** Michael E. Belloy, Yann Le Guen, Ilaria Stewart, Joachim Herz, Richard Sherva, Rui Zhang, Victoria Merritt, Matthew S. Panizzon, Richard L. Hauger, the VA Million Veteran Program, J. Michael Gaziano, Mark Logue, Valerio Napolioni, Michael D. Greicius

## Abstract

**Importance:** The X chromosome has remained enigmatic in Alzheimer’s disease (AD), yet it makes up 5% of the genome and carries a high proportion of genes expressed in the brain, making it particularly appealing as a potential source of unexplored genetic variation in AD.

**Objectives:** Perform the first large-scale X chromosome-wide association study (XWAS) of AD. Primary analyses are non-stratified, while secondary analyses evaluate sex-stratified effects.

**Design:** Meta-analysis of genetic association studies in case-control, family-based, population-based, and longitudinal AD-related cohorts from the US Alzheimer’s Disease Genetics Consortium (ADGC) and Alzheimer’s Disease Sequencing Project (ADSP), the UK Biobank (UKB), the Finnish health registry (FinnGen), and the US Million Veterans Program (MVP). Risk for AD evaluated through case-control logistic regression analyses. Data were analyzed between January 2023 and March 2024.

**Setting:** Genetic data available from high-density single-nucleotide polymorphism (SNP) microarrays and whole-genome sequencing (WGS). Summary statistics for multi-tissue expression and protein quantitative trait loci (QTL) available from published studies, enabling follow-up genetic colocalization analyses.

**Participants:** 1,629,863 eligible participants were selected from referred and volunteer samples, of which 477,596 were excluded for analysis exclusion criteria. Number of participants who declined to participate in original studies was not available.

**Main Outcome and Measures:** Risk for AD (odds ratio; OR) with 95% confidence intervals (CI). Associations were considered at X-chromosome-wide (P-value<1e-5) and genome-wide (P-value<5e-8) significance.

**Results:** Analyses included 1,152,284 non-Hispanic White European ancestry subjects (57.3% females), including 138,558 cases. 6 independent genetic loci passed X-chromosome-wide significance, with 4 showing support for causal links between the genetic signal for AD and expression of nearby genes in brain and non-brain tissues. One of these 4 loci passed conservative genome-wide significance, with its lead variant centered on an intron of *SLC9A7* (OR=1.054, 95%-CI=[1.035, 1.075]) and colocalization analyses prioritizing both the *SLC9A7* and nearby *CHST7* genes.

**Conclusion and Relevance:** We performed the first large-scale XWAS of AD and identified the novel *SLC9A7* locus. *SLC9A7* regulates pH homeostasis in Golgi secretory compartments and is anticipated to have downstream effects on amyloid beta accumulation. Overall, this study significantly advances our knowledge of AD genetics and may provide novel biological drug targets.

**Key points:** *Question:* Does the X chromosome play a role in the genetics of Alzheimer’s Disease (AD)?

*Findings:* In a genetic meta-analysis across 1,152,284 individuals, several X chromosome loci were associated with AD. Four loci showed evidence of shared genetic associations between AD risk and regulation of nearby gene expression in brain tissue. The top association signal was intronic on *SLC9A7* and linked to its expression.

*Meaning:* We performed the first large-scale X chromosome-wide association study of AD and prioritized *SLC9A7* as a novel risk locus. This study significantly advances our knowledge of AD genetics and provides novel biological drug targets.

## Introduction

The X chromosome has remained enigmatic, not just in AD, but in the broader field of genome-wide association studies. It is typically excluded due to technical challenges and power limitations because of its complex inheritance pattern^1^. The X chromosome however makes up 5% of the genome and carries a high proportion of genes expressed in the brain. Additionally, it may contribute to the well-established higher prevalence of AD in women relative to men^2^. We thus set out to fill in this gap by performing the first meta-analysis of XWAS conducted on various publicly available AD-related cohorts, as well as multiple biobanks where AD phenotypes were available. To ensure maximal power, this study was designed as a large-scale discovery combining all available samples.

## Methods

An in-depth overview of all methodologies is provided in the **eMethods**. The current study followed STREGA reporting guidelines. Participants or their caregivers provided written informed consent in the original studies. The current study protocol was granted an exemption by the Stanford Institutional Review Board because the analyses were carried out on “de-identified, off-the-shelf” data; therefore, additional informed consent was not required.

### Data Ascertainment

Case-control, family-based, and longitudinal AD genetic cohorts from the ADGC and ADSP (release-3) were available through public repositories, with genetic data from SNP microarrays and WGS (**eTable1-2**)^3,4^. These cohorts contributed clinically diagnosed AD cases (40.0% pathology verified; **eTable3**). Analyses in UKB, FinnGen, and MVP used genetic data from SNP microarrays^5–8^. UKB data and FinnGen summary results (v10) were publicly available. UKB contributed health-registry-confirmed AD cases and proxy Alzheimer’s disease-and-dementia (ADD) cases; FinnGen contributed health-registry-confirmed AD cases; MVP contributed health-registry-confirmed and proxy ADD cases.

### Quality Control and Processing

ADGC and ADSP data underwent extensive quality control (QC) and imputation to the TOPMed reference panel (**eTable4-5**). Specific consideration was given to X-chromosome QC as in prior work (cf. **eMethods**)^9^. Genetic data processing for UKB, FinnGen, and MVP followed cohort-specific protocols^5–8^. Non-Hispanic White, European ancestry cases and controls, carrying XX or XY with concordant self-reported sex and ages >60 years (>18 and median=63 in FinnGen), were retained for analyses (**eFigure1; eTable3**). Variants were filtered using cohort-specific minor allele frequency (MAF) criteria, which on average correspond to MAF>0.05% (**eTable5**).

### X chromosome Considerations

X chromosome analyses considered non-pseudoautosomal regions. Genotype encoding was 0/2 in men (XY) and 0/1/2 in women (XX), following a random X chromosome inactivation (XCI) model in women. In UKB, most cases were proxy cases, i.e. family history of ADD in first-degree relatives. This proxy approach has been established to replicate AD autosomal genetic risk factors and be adaptable to XWAS^10^. To maximize power, the health-registry and proxy status were unified into a single phenotype for which association coefficients were adjusted onto a regular case-control scale (**eTable6-7**). After rescaling, UKB showed consistent coefficient distributions with ADGC+ADSP (**eFigure2**). A similar approach was used in MVP, but in line with MVP protocols, analyses were separated for health-registry and proxy phenotypes^7^.

### Statistical Analyses

XWAS evaluated case-control logistic regressions on AD risk, adjusting for sex, age, technical covariates, and genetic principal components (capturing population stratification) as applicable per dataset. Mixed models to include related subjects were used in ADGC, ADSP, UKB (LMM-BOLT v2.4)^11^, and FinnGen (Regenie)^6^. Association results across datasets were combined through fixed effects inverse-variance weighted meta-analyses. Primary analyses were non-stratified. Secondary analyses were sex-stratified, and conducted across ADGC, ADSP, and UKB. Association results were considered at the X-chromosome-wide (P-value<1e-5) and conservative genome-wide thresholds (P-value<5e-8). Sex effects were evaluated through heterogeneity tests and considered significant at P<0.05. Evidence for escape from XCI was evaluated by comparing variant beta coefficients derived from men and women-stratified XWAS (ratio=1 indicates escape; ratio=2 indicates no escape)^12^.

### Genetic Colocalization

To identify potentially causal genes in associated risk loci, statistical colocalization was evaluated between the local genetic association signal for AD and the genetic association signal for molecular traits such as expression levels of genes within that locus (R-v.4.2.1, *coloc*)^13^. We leveraged public datasets where quantitative trait loci (QTL) for expression and protein levels were available for the X chromosome in brain and non-brain tissues (cf. **eMethods**).

## Results

The study design is provided in **Figure1A**. 1,152,284 individuals (138,558 cases: 15,081 clinically diagnosed cases, 41,091 health-registry-confirmed cases, and 82,386 proxy cases) were included in the XWAS (**eTable3**). There was no sign of genomic inflation (**eFigure3**). We associated 2 rare (MAF<1%) lead variants in the *NLGN4X* and *MID1* loci, and 4 common lead variants in the *SLC9A7*, *ZNF280C*, *ARGRG4*, and *MTM1* loci (**Table1**; locus zoom and forest plots in **eFigures4-5**). All common variant loci showed colocalization for at least one nearby gene in brain tissue (**Table2; eTable8**). The overall top association signal (cross-cohort allele frequencies in **eTable9**), intronic on *SLC9A7*, passed conservative significance criteria and showed colocalization for several genes, most notably *SLC9A7* and *CHST7*. Colocalization plots for top prioritized genes are in **eFigures6-10**.

The *ZNF280C* and *ARGRG4* lead variants showed evidence for escape from XCI, while the *MID1* variant appeared female-specific (**eTable10**). Sex-stratified XWAS only revealed 1 X-chromosome-wide significant, female-specific rare variant association without colocalization support (**eFigure11**, **eTable11**) and indicated that evidence for escape from XCI was apparent only for a few common, small effect size variants (**eFigure12**).

**Figure 1.**
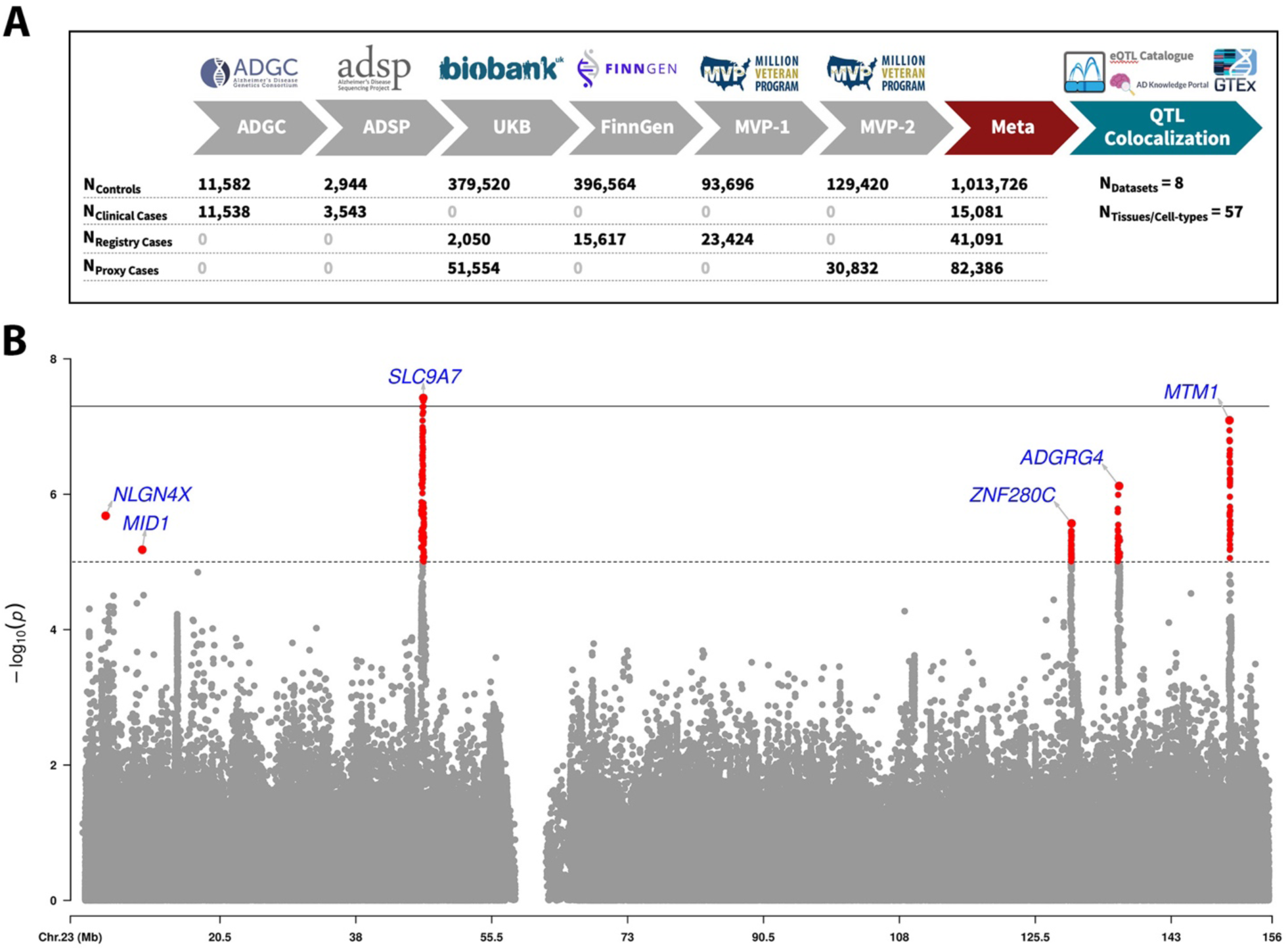
X chromosome-wide association study of Alzheimer’s disease. A) Overview of study design and sample sizes. To increase specificity to AD (rather than ADD), the XWAS meta-analysis was intersected to variants with association results in ADGC (which used only clinically confirmed cases and controls). **B)** Manhattan plot for the XWAS meta-analysis. The dotted line indicates X-chromosome-wide significance (P-value<1e-5) and full line indicates genome-wide significance (P-value<5e-8). Lead variants for independent loci are annotated with their nearest protein-coding gene (Gencode v42).

**Table 1.**
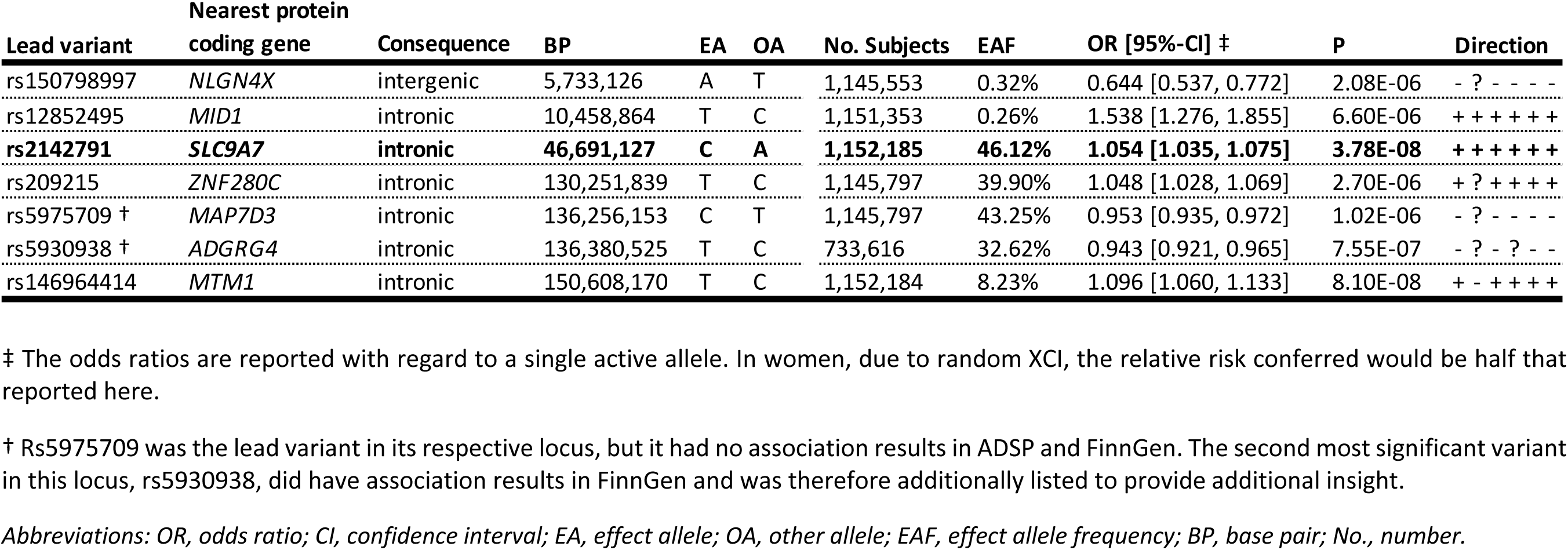
X chromosome-wide association study of Alzheimer’s disease: Associated lead variants. The Direction column indicates the association effect direction across meta-analyzed cohorts following the order of ADGC, ADSP, UKB, FinnGen, MVP-1 (using health registry status), and MVP-2 (using proxy status). A question mark indicates the variant was not available in the respective cohort. Variants are annotated using dbSNP153. Association signals passing genome-wide significance are bolded.

**Table 2.**
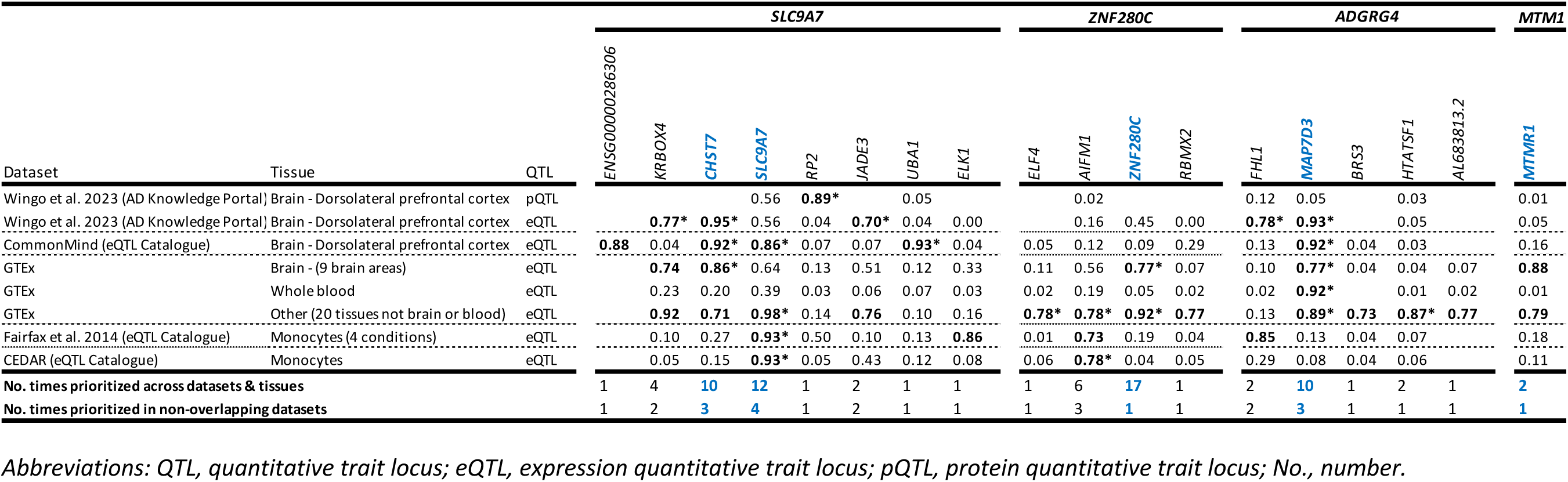
Genetic colocalization with quantitative trait locus data. Colocalization was evaluated for genes in each AD associated locus using a 2Mb window centered on the lead variant. Evidence for colocalization was considered at colocalization posterior probability (PP4)>0.7 (bolded). The table presents PP4 results and is restricted to genes and datasets/tissues where at least one colocalization reached PP4>0.7. As such, the table is partitioned into 4 common variant loci that showed colocalization support. Bolded entries with an asterisk (*) indicate the lead variant was also a significant QTL in the respective data/tissue. Missing entries indicate that no QTL data were available. The total number of times a gene was prioritized (PP4>0.7) is summarized to help identify the most likely causal gene per locus (blue bolded genes and numbers). Overlapping datasets were considered as those where subjects partially or fully overlapped (non-overlapping datasets are separated by dashed lines).

## Discussion

We performed an XWAS of AD in 1,152,284 individuals, making this the largest genetic association study of AD to date^14^. The top signal showed support for a causal link between the genetic regulation of *SLC9A7* or *CHST7* expression and AD risk. *CHST7* encodes a chondroitin 6-sulfotransferase that confers negatively charged sulfate groups to glycosaminoglycans, which may relate to promoting tau fibrillization and spreading^15^. Notably, *SLC9A7* (a.k.a *NHE7*) is a paralog of *SLC9A6* (a.k.a *NHE6*), previously implicated in experimental work as an X-linked AD modifying gene^16^. These are highly conserved genes that regulate pH homeostasis in Golgi secretory compartments and endosomes and might thus be expected to contribute to increased amyloid accumulation across aging when their expression levels are increased (a detailed background and rationale are provided in **Appendix-A**). In line with this expectation, QTL data support that the top risk allele is associated with increased expression of *SLC9A7* in brain tissue, increasing expression by 17-44% for an active allele (**eTable12**). Although the *SLC9A7* top variant has a small effect size (OR=1.054, 95%-CI=[1.035, 1.075]), given this relatively small effect on *SLC9A7* expression in the brain, it may be that more substantial reduction or pharmacological inhibition of *SLC9A7* would prove to be an effective therapeutic strategy for AD.

Despite this study’s formidable sample size, only the *SLC9A7* locus reached conservative significance criteria with a small effect size, suggesting the X chromosome contributes relatively little to AD prevalence. In addition, only 2 lead variants of small effect size indicated escape from XCI and only 1 rare lead variant appeared female-specific, such that these XWAS results have little bearing on sex-stratified AD prevalence. Similarly, sex-stratified XWAS did not reveal striking results, which would have been expected if the X chromosome played a significant role in the observations that 2/3 of AD patients across the lifespan are women^2^. Overall, our results suggest that while the X chromosome plays only a small role in the population prevalence of AD, the specific pathways highlighted here open the door to novel pathogenic pathways and associated drug targets.

### Limitations

This study focused on European ancestry individuals. When larger cross-ancestry samples become available, future studies should extend AD XWAS into these populations. Similarly, future, larger sex-stratified AD XWAS may help identify sex-specific risk genes and genes escaping from XCI. Lastly, this study did not provide conclusive insight into the causal gene at the *SLC9A7* locus, which future experimental studies should interrogate.

## Conclusion

We performed the first large-scale XWAS of AD and identified the novel *SLC9A7* risk locus. Overall, this study significantly advances our knowledge of the genetics of AD and may provide novel biological drug targets.

## Consent for publication

Not applicable.

## Availability of data and materials

Data used in the XWAS are available upon application to:

- dbGaP (https://www.ncbi.nlm.nih.gov/gap/)

- NIAGADS (https://www.niagads.org/)

- LONI (https://ida.loni.usc.edu/)

- AMP-AD knowledge portal / Synapse (https://www.synapse.org/)

- Rush (https://www.radc.rush.edu/)

- NACC (https://naccdata.org/)

- UKB (https://www.ukbiobank.ac.uk/)

- FinnGen (https://www.finngen.fi/en)

- MVP (https://www.mvp.va.gov/)

The specific data repository and identifier for ADGC and ADSP data are indicated in **eTable1** of the supplement.

The data, code, and phenotypes used to generate MVP results are accessible to researchers with MVP data access. Due to VA policy, MVP is currently only accessible to VA researchers with a funded MVP project, either through a VA Merit Award, career development award, or NIH R01. Additional information is available at https://genhub.va.gov/file/view/897656. GWAS summary results for the MVP cohort will be posted to dbGAP after publication.

Colocalization datasets are available from:

- AMP-AD knowledge portal / Synapse (https://www.synapse.org/; identifier: syn51150434)

- eQTL catalogue (https://www.ebi.ac.uk/eqtl/)

- GTEx (https://www.gtexportal.org/home/)

Summary statistics generated by this study will be deposited in both NIAGADS and the EMBL-EBI GWAS Catalog.

## Authors’ contributions

M.E.B., Y.L. and M.D.G. had full access to all the data in the study and take responsibility for the integrity of the data and the accuracy of the data analysis. M.E.B. performed data acquisition and analyses, designed analyses, designed study, wrote paper, and obtained funding. Y.L. performed data acquisition and analyses, designed analyses, designed study, and wrote paper. I.S. performed data acquisition and analyses. J.H. co-wrote the paper. M.W.L. and J.M.G were involved in data, funding, and resource acquisition. M.W.L, R.S., V.M., M.S.P., and R.L.H. were involved in conceptualization and study design. R.Z and R.S. analyzed and curated data. V.N. performed data acquisition and analyses, designed study, designed analyses, supervised analyses, and supervised work. M.D.G designed study, designed analyses, supervised analyses, supervised work, wrote paper, and obtained funding. All authors contributed to critical revision of the manuscript.

## Data Availability

Data used in the XWAS are available upon application to:
- dbGaP (https://www.ncbi.nlm.nih.gov/gap/)
- NIAGADS (https://www.niagads.org/)
- LONI (https://ida.loni.usc.edu/)
- AMP-AD knowledge portal / Synapse (https://www.synapse.org/)
- Rush (https://www.radc.rush.edu/)
- NACC (https://naccdata.org/)
- UKB (https://www.ukbiobank.ac.uk/)
- FinnGen (https://www.finngen.fi/en)
- MVP (https://www.mvp.va.gov/)
The specific data repository and identifier for ADGC and ADSP data are indicated in eTable1 of the supplement.
The data, code, and phenotypes used to generate MVP results are accessible to researchers with MVP data access. Due to VA policy, MVP is currently only accessible to VA researchers with a funded MVP project, either through a VA Merit Award, career development award, or NIH R01. Additional information is available at https://genhub.va.gov/file/view/897656. GWAS summary results for the MVP cohort will be posted to dbGAP after publication.
Colocalization datasets are available from:
- AMP-AD knowledge portal / Synapse (https://www.synapse.org/; identifier: syn51150434)
- eQTL catalogue (https://www.ebi.ac.uk/eqtl/)
- GTEx (https://www.gtexportal.org/home/)
Summary statistics generated by this study will be deposited in both NIAGADS and the EMBL-EBI GWAS Catalog.

https://www.ncbi.nlm.nih.gov/gap/

https://www.niagads.org/

https://ida.loni.usc.edu/

https://www.synapse.org/

https://www.radc.rush.edu/

https://naccdata.org/

https://www.ukbiobank.ac.uk/

https://www.finngen.fi/en

https://www.mvp.va.gov/

https://www.ebi.ac.uk/eqtl/

## Acknowledgements

Data for this study were prepared, archived, and distributed by the National Institute on Aging Alzheimer’s Disease Data Storage Site (NIAGADS) at the University of Pennsylvania (U24-AG041689), funded by the National Institute on Aging. The contents of this article do not represent the views of the National Institutes of Health, the U.S. Department of Veterans Affairs, or the United States Government.

## Acknowledgments for the use of ADSP WES and WGS data

The Alzheimer’s Disease Sequencing Project (ADSP) is comprised of two Alzheimer’s Disease (AD) genetics consortia and three National Human Genome Research Institute (NHGRI) funded Large Scale Sequencing and Analysis Centers (LSAC). The two AD genetics consortia are the Alzheimer’s Disease Genetics Consortium (ADGC) funded by NIA (U01 AG032984), and the Cohorts for Heart and Aging Research in Genomic Epidemiology (CHARGE) funded by NIA (R01 AG033193), the National Heart, Lung, and Blood Institute (NHLBI), other National Institute of Health (NIH) institutes and other foreign governmental and non-governmental organizations. The Discovery Phase analysis of sequence data is supported through UF1AG047133 (to Drs. Schellenberg, Farrer, Pericak-Vance, Mayeux, and Haines); U01AG049505 to Dr. Seshadri; U01AG049506 to Dr. Boerwinkle; U01AG049507 to Dr. Wijsman; and U01AG049508 to Dr. Goate and the Discovery Extension Phase analysis is supported through U01AG052411 to Dr. Goate, U01AG052410 to Dr. Pericak-Vance and U01 AG052409 to Drs. Seshadri and Fornage.

Sequencing for the Follow Up Study (FUS) is supported through U01AG057659 (to Drs. PericakVance, Mayeux, and Vardarajan) and U01AG062943 (to Drs. Pericak-Vance and Mayeux). Data generation and harmonization in the Follow-up Phase is supported by U54AG052427 (to Drs. Schellenberg and Wang). The FUS Phase analysis of sequence data is supported through U01AG058589 (to Drs. Destefano, Boerwinkle, De Jager, Fornage, Seshadri, and Wijsman), U01AG058654 (to Drs. Haines, Bush, Farrer, Martin, and Pericak-Vance), U01AG058635 (to Dr. Goate), RF1AG058066 (to Drs. Haines, Pericak-Vance, and Scott), RF1AG057519 (to Drs. Farrer and Jun), R01AG048927 (to Dr. Farrer), and RF1AG054074 (to Drs. Pericak-Vance and Beecham).

The ADGC cohorts include: Adult Changes in Thought (ACT) (UO1 AG006781, UO1 HG004610, UO1 HG006375, U01 HG008657), the Alzheimer’s Disease Centers (ADC) ( P30 AG019610, P30 AG013846, P50 AG008702, P50 AG025688, P50 AG047266, P30 AG010133, P50 AG005146, P50 AG005134, P50 AG016574, P50 AG005138, P30 AG008051, P30 AG013854, P30 AG008017, P30 AG010161, P50 AG047366, P30 AG010129, P50 AG016573, P50 AG016570, P50 AG005131, P50 AG023501, P30 AG035982, P30 AG028383, P30 AG010124, P50 AG005133, P50 AG005142, P30 AG012300, P50 AG005136, P50 AG033514, P50 AG005681, and P50 AG047270), the Chicago Health and Aging Project (CHAP) (R01 AG11101, RC4 AG039085, K23 AG030944), Indianapolis Ibadan (R01 AG009956, P30 AG010133), the Memory and Aging Project (MAP) ( R01 AG17917), Mayo Clinic (MAYO) (R01 AG032990, U01 AG046139, R01 NS080820, RF1 AG051504, P50 AG016574), Mayo Parkinson’s Disease controls (NS039764, NS071674, 5RC2HG005605), University of Miami (R01 AG027944, R01 AG028786, R01 AG019085, IIRG09133827, A2011048), the Multi-Institutional Research in Alzheimer’s Genetic Epidemiology Study (MIRAGE) (R01 AG09029, R01 AG025259), the National Cell Repository for Alzheimer’s Disease (NCRAD) (U24 AG21886), the National Institute on Aging Late Onset Alzheimer’s Disease Family Study (NIA- LOAD) (R01 AG041797), the Religious Orders Study (ROS) (P30 AG10161, R01 AG15819), the Texas Alzheimer’s Research and Care Consortium (TARCC) (funded by the Darrell K Royal Texas Alzheimer’s Initiative), Vanderbilt University/Case Western Reserve University (VAN/CWRU) (R01 AG019757, R01 AG021547, R01 AG027944, R01 AG028786, P01 NS026630, and Alzheimer’s Association), the Washington Heights-Inwood Columbia Aging Project (WHICAP) (RF1 AG054023), the University of Washington Families (VA Research Merit Grant, NIA: P50AG005136, R01AG041797, NINDS: R01NS069719), the Columbia University HispanicEstudio Familiar de Influencia Genetica de Alzheimer (EFIGA) (RF1 AG015473), the University of Toronto (UT) (funded by Wellcome Trust, Medical Research Council, Canadian Institutes of Health Research), and Genetic Differences (GD) (R01 AG007584). The CHARGE cohorts are supported in part by National Heart, Lung, and Blood Institute (NHLBI) infrastructure grant HL105756 (Psaty), RC2HL102419 (Boerwinkle) and the neurology working group is supported by the National Institute on Aging (NIA) R01 grant AG033193.

The CHARGE cohorts participating in the ADSP include the following: Austrian Stroke Prevention Study (ASPS), ASPS-Family study, and the Prospective Dementia Registry-Austria (ASPS/PRODEM-Aus), the Atherosclerosis Risk in Communities (ARIC) Study, the Cardiovascular Health Study (CHS), the Erasmus Rucphen Family Study (ERF), the Framingham Heart Study (FHS), and the Rotterdam Study (RS). ASPS is funded by the Austrian Science Fond (FWF) grant number P20545-P05 and P13180 and the Medical University of Graz. The ASPS-Fam is funded by the Austrian Science Fund (FWF) project I904),the EU Joint Programme - Neurodegenerative Disease Research (JPND) in frame of the BRIDGET project (Austria, Ministry of Science) and the Medical University of Graz and the Steiermärkische Krankenanstalten Gesellschaft. PRODEM-Austria is supported by the Austrian Research Promotion agency (FFG) (Project No. 827462) and by the Austrian National Bank (Anniversary Fund, project 15435. ARIC research is carried out as a collaborative study supported by NHLBI contracts (HHSN268201100005C, HHSN268201100006C, HHSN268201100007C, HHSN268201100008C, HHSN268201100009C, HHSN268201100010C, HHSN268201100011C, and HHSN268201100012C). Neurocognitive data in ARIC is collected by U01 2U01HL096812, 2U01HL096814, 2U01HL096899, 2U01HL096902, 2U01HL096917 from the NIH (NHLBI, NINDS, NIA and NIDCD), and with previous brain MRI examinations funded by R01-HL70825 from the NHLBI. CHS research was supported by contracts HHSN268201200036C, HHSN268200800007C, N01HC55222, N01HC85079, N01HC85080, N01HC85081, N01HC85082, N01HC85083, N01HC85086, and grants U01HL080295 and U01HL130114 from the NHLBI with additional contribution from the National Institute of Neurological Disorders and Stroke (NINDS). Additional support was provided by R01AG023629, R01AG15928, and R01AG20098 from the NIA. FHS research is supported by NHLBI contracts N01-HC- 25195 and HHSN268201500001I. This study was also supported by additional grants from the NIA (R01s AG054076, AG049607 and AG033040 and NINDS (R01 NS017950). The ERF study as a part of EUROSPAN (European Special Populations Research Network) was supported by European Commission FP6 STRP grant number 018947 (LSHG-CT-2006-01947) and also received funding from the European Community’s Seventh Framework Programme (FP7/2007-2013)/grant agreement HEALTH-F4- 2007-201413 by the European Commission under the programme "Quality of Life and Management of the Living Resources" of 5th Framework Programme (no. QLG2-CT-2002- 01254). High-throughput analysis of the ERF data was supported by a joint grant from the Netherlands Organization for Scientific Research and the Russian Foundation for Basic Research (NWO-RFBR 047.017.043). The Rotterdam Study is funded by Erasmus Medical Center and Erasmus University, Rotterdam, the Netherlands Organization for Health Research and Development (ZonMw), the Research Institute for Diseases in the Elderly (RIDE), the Ministry of Education, Culture and Science, the Ministry for Health, Welfare and Sports, the European Commission (DG XII), and the municipality of Rotterdam. Genetic data sets are also supported by the Netherlands Organization of Scientific Research NWO Investments (175.010.2005.011, 911-03-012), the Genetic Laboratory of the Department of Internal Medicine, Erasmus MC, the Research Institute for Diseases in the Elderly (014-93-015; RIDE2), and the Netherlands Genomics Initiative (NGI)/Netherlands Organization for Scientific Research (NWO) Netherlands Consortium for Healthy Aging (NCHA), project 050-060-810. All studies are grateful to their participants, faculty and staff. The content of these manuscripts is solely the responsibility of the authors and does not necessarily represent the official views of the National Institutes of Health or the U.S. Department of Health and Human Services.

The FUS cohorts include: the Alzheimer’s Disease Centers (ADC) ( P30 AG019610, P30 AG013846, P50 AG008702, P50 AG025688, P50 AG047266, P30 AG010133, P50 AG005146, P50 AG005134, P50 AG016574, P50 AG005138, P30 AG008051, P30 AG013854, P30 AG008017, P30 AG010161, P50 AG047366, P30 AG010129, P50 AG016573, P50 AG016570, P50 AG005131, P50 AG023501, P30 AG035982, P30 AG028383, P30 AG010124, P50 AG005133, P50 AG005142, P30 AG012300, P50 AG005136, P50 AG033514, P50 AG005681, and P50 AG047270), Alzheimer’s Disease Neuroimaging Initiative (ADNI) (U19AG024904), Amish Protective Variant Study (RF1AG058066), Cache County Study (R01AG11380, R01AG031272, R01AG21136, RF1AG054052), Case Western Reserve University Brain Bank (CWRUBB) (P50AG008012), Case Western Reserve University Rapid Decline (CWRURD) (RF1AG058267, NU38CK000480), CubanAmerican Alzheimer’s Disease Initiative (CuAADI) (3U01AG052410), Estudio Familiar de Influencia Genetica en Alzheimer (EFIGA) (5R37AG015473, RF1AG015473, R56AG051876), Genetic and Environmental Risk Factors for Alzheimer Disease Among African Americans Study (GenerAAtions) (2R01AG09029, R01AG025259, 2R01AG048927), Gwangju Alzheimer and Related Dementias Study (GARD) (U01AG062602), Hussman Institute for Human Genomics Brain Bank (HIHGBB) (R01AG027944, Alzheimer’s Association "Identification of Rare Variants in Alzheimer Disease"), Ibadan Study of Aging (IBADAN) (5R01AG009956), Mexican Health and Aging Study (MHAS) (R01AG018016), Multi-Institutional Research in Alzheimer’s Genetic Epidemiology (MIRAGE) (2R01AG09029, R01AG025259, 2R01AG048927), Northern Manhattan Study (NOMAS) (R01NS29993), Peru Alzheimer’s Disease Initiative (PeADI) (RF1AG054074), Puerto Rican 1066 (PR1066) (Wellcome Trust (GR066133/GR080002), European Research Council (340755)), Puerto Rican Alzheimer Disease Initiative (PRADI) (RF1AG054074), Reasons for Geographic and Racial Differences in Stroke (REGARDS) (U01NS041588), Research in African American Alzheimer Disease Initiative (REAAADI) (U01AG052410), Rush Alzheimer’s Disease Center (ROSMAP) (P30AG10161, R01AG15819, R01AG17919), University of Miami Brain Endowment Bank (MBB), and University of Miami/Case Western/North Carolina A&T African American (UM/CASE/NCAT) (U01AG052410, R01AG028786).

The four LSACs are: the Human Genome Sequencing Center at the Baylor College of Medicine (U54 HG003273), the Broad Institute Genome Center (U54HG003067), The American Genome Center at the Uniformed Services University of the Health Sciences (U01AG057659), and the Washington University Genome Institute (U54HG003079).

Biological samples and associated phenotypic data used in primary data analyses were stored at Study Investigators institutions, and at the National Cell Repository for Alzheimer’s Disease (NCRAD, U24AG021886) at Indiana University funded by NIA. Associated Phenotypic Data used in primary and secondary data analyses were provided by Study Investigators, the NIA funded Alzheimer’s Disease Centers (ADCs), and the National Alzheimer’s Coordinating Center (NACC, U01AG016976) and the National Institute on Aging Genetics of Alzheimer’s Disease Data Storage Site (NIAGADS, U24AG041689) at the University of Pennsylvania, funded by NIA This research was supported in part by the Intramural Research Program of the National Institutes of health, National Library of Medicine. Contributors to the Genetic Analysis Data included Study Investigators on projects that were individually funded by NIA, and other NIH institutes, and by private U.S. organizations, or foreign governmental or nongovernmental organizations.

An up to date acknowledgment statement can be found on the ADSP site: https://www.niagads.org/adsp/content/acknowledgement-statement.

Data collection and sharing for this project was funded by the Alzheimer’s Disease Neuroimaging Initiative (ADNI) (National Institutes of Health Grant U01 AG024904) and DOD ADNI (Department of Defense award number W81XWH-12-2-0012). ADNI is funded by the National Institute on Aging, the National Institute of Biomedical Imaging and Bioengineering, and through generous contributions from the following: AbbVie, Alzheimer’s Association; Alzheimer’s Drug Discovery Foundation; Araclon Biotech; BioClinica, Inc.; Biogen; Bristol-Myers Squibb Company; CereSpir, Inc.; Cogstate; Eisai Inc.; Elan Pharmaceuticals, Inc.; Eli Lilly and Company; EuroImmun; F. Hoffmann-La Roche Ltd and its affiliated company Genentech, Inc.; Fujirebio; GE Healthcare; IXICO Ltd.; Janssen Alzheimer Immunotherapy Research & Development, LLC.; Johnson & Johnson Pharmaceutical Research & Development LLC.; Lumosity; Lundbeck; Merck & Co., Inc.; Meso Scale Diagnostics, LLC.; NeuroRx Research; Neurotrack Technologies; Novartis Pharmaceuticals Corporation; Pfizer Inc.; Piramal Imaging; Servier; Takeda Pharmaceutical Company; and Transition Therapeutics. The Canadian Institutes of Health Research is providing funds to support ADNI clinical sites in Canada. Private sector contributions are facilitated by the Foundation for the National Institutes of Health (www.fnih.org). The grantee organization is the Northern California Institute for Research and Education, and the study is coordinated by the Alzheimer’s Therapeutic Research Institute at the University of Southern California. ADNI data are disseminated by the Laboratory for Neuro Imaging at the University of Southern California.

Additional information to include in an acknowledgment statement can be found on the LONI site: https://adni.loni.usc.edu/wp-content/uploads/how_to_apply/ADNI_Data_Use_Agreement.pdf.

The Alzheimer’s Disease Genetics Consortium (ADGC) supported sample preparation, whole exome sequencing and data processing through NIA grant U01AG032984. Sequencing data generation and harmonization is supported by the Genome Center for Alzheimer’s Disease, U54AG052427, and data sharing is supported by NIAGADS, U24AG041689. Samples from the National Centralized Repository for Alzheimer’s Disease and Related Dementias (NCRAD), which receives government support under a cooperative agreement grant (U24 AG021886) awarded by the National Institute on Aging (NIA), were used in this study. We thank contributors who collected samples used in this study, as well as patients and their families, whose help and participation made this work possible. NIH grants supported enrollment and data collection for the individual studies including: GenerAAtions R01AG20688 (PI M. Daniele Fallin, PhD); Miami/Duke R01 AG027944, R01 AG028786 (PI Margaret A. Pericak-Vance, PhD); NC A&T P20 MD000546, R01 AG28786-01A1 (PI Goldie S. Byrd, PhD); Case Western (PI Jonathan L. Haines, PhD); MIRAGE R01 AG009029 (PI Lindsay A. Farrer, PhD); ROS P30AG10161, R01AG15819, R01AG30146, TGen (PI David A. Bennett, MD); MAP R01AG17917, R01AG15819, TGen (PI David A. Bennett, MD). The NACC database is funded by NIA/NIH Grant U01 AG016976. NACC data are contributed by the NIA-funded ADCs: P30 AG019610 (PI Eric Reiman, MD), P30 AG013846 (PI Neil Kowall, MD), P30 AG062428-01 (PI James Leverenz, MD) P50 AG008702 (PI Scott Small, MD), P50 AG025688 (PI Allan Levey, MD, PhD), P50 AG047266 (PI Todd Golde, MD, PhD), P30 AG010133 (PI Andrew Saykin, PsyD), P50 AG005146 (PI Marilyn Albert, PhD), P30 AG062421-01 (PI Bradley Hyman, MD, PhD), P30 AG062422-01 (PI Ronald Petersen, MD, PhD), P50 AG005138 (PI Mary Sano, PhD), P30 AG008051 (PI Thomas Wisniewski, MD), P30 AG013854 (PI Robert Vassar, PhD), P30 AG008017 (PI Jeffrey Kaye, MD), P30 AG010161 (PI David Bennett, MD), P50 AG047366 (PI Victor Henderson, MD, MS), P30 AG010129 (PI Charles DeCarli, MD), P50 AG016573 (PI Frank LaFerla, PhD), P30 AG062429-01(PI James Brewer, MD, PhD), P50 AG023501 (PI Bruce Miller, MD), P30 AG035982 (PI Russell Swerdlow, MD), P30 AG028383 (PI Linda Van Eldik, PhD), P30 AG053760 (PI Henry Paulson, MD, PhD), P30 AG010124 (PI John Trojanowski, MD, PhD), P50 AG005133 (PI Oscar Lopez, MD), P50 AG005142 (PI Helena Chui, MD), P30 AG012300 (PI Roger Rosenberg, MD), P30 AG049638 (PI Suzanne Craft, PhD), P50 AG005136 (PI Thomas Grabowski, MD), P30 AG062715-01 (PI Sanjay Asthana, MD, FRCP), P50 AG005681 (PI John Morris, MD), P50 AG047270 (PI Stephen Strittmatter, MD, PhD).

This work was supported by grants from the National Institutes of Health (R01AG044546, P01AG003991, RF1AG053303, R01AG058501, U01AG058922, RF1AG058501 and R01AG057777). The recruitment and clinical characterization of research participants at Washington University were supported by NIH P50 AG05681, P01 AG03991, and P01 AG026276. This work was supported by access to equipment made possible by the Hope Center for Neurological Disorders, and the Departments of Neurology and Psychiatry at Washington University School of Medicine.

We thank the contributors who collected samples used in this study, as well as patients and their families, whose help and participation made this work possible. Members of the National Institute on Aging Late-Onset Alzheimer Disease/National Cell Repository for Alzheimer Disease (NIA-LOAD NCRAD) Family Study Group include the following: Richard Mayeux, MD, MSc; Martin Farlow, MD; Tatiana Foroud, PhD; Kelley Faber, MS; Bradley F. Boeve, MD; Neill R. Graff-Radford, MD; David A. Bennett, MD; Robert A. Sweet, MD; Roger Rosenberg, MD; Thomas D. Bird, MD; Carlos Cruchaga, PhD; and Jeremy M. Silverman, PhD.

This work was partially supported by grant funding from NIH R01 AG039700 and NIH P50 AG005136. Subjects and samples used here were originally collected with grant funding from NIH U24 AG026395, U24 AG021886, P50 AG008702, P01 AG007232, R37 AG015473, P30 AG028377, P50 AG05128, P50 AG16574, P30 AG010133, P50 AG005681, P01 AG003991, U01MH046281, U01 MH046290 and U01 MH046373. The funders had no role in study design, analysis or preparation of the manuscript. The authors declare no competing interests.

This work was supported by the National Institutes of Health (R01 AG027944, R01 AG028786 to MAPV, R01 AG019085 to JLH, P20 MD000546); a joint grant from the Alzheimer’s Association (SG-14-312644) and the Fidelity Biosciences Research Initiative to MAPV; the BrightFocus Foundation (A2011048 to MAPV). NIA-LOAD Family-Based Study supported the collection of samples used in this study through NIH grants U24 AG026395 and R01 AG041797 and the MIRAGE cohort was supported through the NIH grants R01 AG025259 and R01 AG048927. We thank contributors, including the Alzheimer’s disease Centers who collected samples used in this study, as well as patients and their families, whose help and participation made this work possible. Study design: HNC, BWK, JLH, MAPV; Sample collection: MLC, JMV, RMC, LAF, JLH, MAPV; Whole exome sequencing and Sanger sequencing: SR, PLW; Sequencing data analysis: HNC, BWK, KLHN, SR, MAK, JRG, ERM, GWB, MAPV; Statistical analysis: BWK, KLHN, JMJ, MAPV; Preparation of manuscript: HNC, BWK. The authors jointly discussed the experimental results throughout the duration of the study. All authors read and approved the final manuscript.

Data collection and sharing for this project was supported by the Washington Heights-Inwood Columbia Aging Project (WHICAP, PO1AG07232, R01AG037212, RF1AG054023) funded by the National Institute on Aging (NIA) and by the National Center for Advancing Translational Sciences, National Institutes of Health, through Grant Number UL1TR001873. This manuscript has been reviewed by WHICAP investigators for scientific content and consistency of data interpretation with previous WHICAP Study publications. We acknowledge the WHICAP study participants and the WHICAP research and support staff for their contributions to this study.

This work was supported by grants from the National Institutes of Health (R01AG044546, P01AG003991, RF1AG053303, R01AG058501, U01AG058922, RF1AG058501 and R01AG057777). The recruitment and clinical characterization of research participants at Washington University were supported by NIH P50 AG05681, P01 AG03991, and P01 AG026276. This work was supported by access to equipment made possible by the Hope Center for Neurological Disorders, and the Departments of Neurology and Psychiatry at Washington University School of Medicine.

We thank the contributors who collected samples used in this study, as well as patients and their families, whose help and participation made this work possible. Members of the National Institute on Aging Late-Onset Alzheimer Disease/National Cell Repository for Alzheimer Disease (NIA-LOAD NCRAD) Family Study Group include the following: Richard Mayeux, MD, MSc; Martin Farlow, MD; Tatiana Foroud, PhD; Kelley Faber, MS; Bradley F. Boeve, MD; Neill R. Graff-Radford, MD; David A. Bennett, MD; Robert A. Sweet, MD; Roger Rosenberg, MD; Thomas D. Bird, MD; Carlos Cruchaga, PhD; and Jeremy M. Silverman, PhD.

This work was supported by grants from the National Institutes of Health (R01AG044546, P01AG003991, RF1AG053303, R01AG058501, U01AG058922, RF1AG058501 and R01AG057777). The recruitment and clinical characterization of research participants at Washington University were supported by NIH P50 AG05681, P01 AG03991, and P01 AG026276. This work was supported by access to equipment made possible by the Hope Center for Neurological Disorders, and the Departments of Neurology and Psychiatry at Washington University School of Medicine.

We thank the contributors who collected samples used in this study, as well as patients and their families, whose help and participation made this work possible. Members of the National Institute on Aging Late-Onset Alzheimer Disease/National Cell Repository for Alzheimer Disease (NIA-LOAD NCRAD) Family Study Group include the following: Richard Mayeux, MD, MSc; Martin Farlow, MD; Tatiana Foroud, PhD; Kelley Faber, MS; Bradley F. Boeve, MD; Neill R. Graff-Radford, MD; David A. Bennett, MD; Robert A. Sweet, MD; Roger Rosenberg, MD; Thomas D. Bird, MD; Carlos Cruchaga, PhD; and Jeremy M. Silverman, PhD.

Mayo RNAseq Study- Study data were provided by the following sources: The Mayo Clinic Alzheimer’s Disease Genetic Studies, led by Dr. Nilufer Ertekin-Taner and Dr. Steven G. Younkin, Mayo Clinic, Jacksonville, FL using samples from the Mayo Clinic Study of Aging, the Mayo Clinic Alzheimer’s Disease Research Center, and the Mayo Clinic Brain Bank. Data collection was supported through funding by NIA grants P50 AG016574, R01 AG032990, U01 AG046139, R01 AG018023, U01 AG006576, U01 AG006786, R01 AG025711, R01 AG017216, R01 AG003949, NINDS grant R01 NS080820, CurePSP Foundation, and support from Mayo Foundation. Study data includes samples collected through the Sun Health Research Institute Brain and Body Donation Program of Sun City, Arizona. The Brain and Body Donation Program is supported by the National Institute of Neurological Disorders and Stroke (U24 NS072026 National Brain and Tissue Resource for Parkinson’s Disease and Related Disorders), the National Institute on Aging (P30 AG19610 Arizona Alzheimer’s Disease Core Center), the Arizona Department of Health Services (contract 211002, Arizona Alzheimer’s Research Center), the Arizona Biomedical Research Commission (contracts 4001, 0011, 05-901 and 1001 to the Arizona Parkinson’s Disease Consortium) and the Michael J. Fox Foundation for Parkinson’s Research ROSMAP- We are grateful to the participants in the Religious Order Study, the Memory and Aging Project. This work is supported by the US National Institutes of Health [U01 AG046152, R01 AG043617, R01 AG042210, R01 AG036042, R01 AG036836, R01 AG032990, R01 AG18023, RC2 AG036547, P50 AG016574, U01 ES017155, KL2 RR024151, K25 AG041906-01, R01 AG30146, P30 AG10161, R01 AG17917, R01 AG15819, K08 AG034290, P30 AG10161 and R01 AG11101.

Mount Sinai Brain Bank (MSBB)- This work was supported by the grants R01AG046170, RF1AG054014, RF1AG057440 and R01AG057907 from the NIH/National Institute on Aging (NIA). R01AG046170 is a component of the AMP-AD Target Discovery and Preclinical Validation Project. Brain tissue collection and characterization was supported by NIH HHSN271201300031C.

This study was supported by the National Institute on Aging (NIA) grants AG030653, AG041718, AG064877 and P30-AG066468.

We would like to thank study participants, their families, and the sample collectors for their invaluable contributions. This research was supported in part by the National Institute on Aging grant U01AG049508 (PI Alison M. Goate). This research was supported in part by Genentech, Inc. (PI Alison M. Goate, Robert R. Graham).

The NACC database is funded by NIA/NIH Grant U01 AG016976. NACC data are contributed by these NIA-funded ADCs: P30 AG013846 (PI Neil Kowall, MD), P50 AG008702 (PI Scott Small, MD), P50 AG025688 (PI Allan Levey, MD, PhD), P30 AG010133 (PI Andrew Saykin, PsyD), P50 AG005146 (PI Marilyn Albert, PhD), P50 AG005134 (PI Bradley Hyman, MD, PhD), P50 AG016574 (PI Ronald Petersen, MD, PhD), P30 AG013854 (PI M. Marsel Mesulam, MD), P30 AG008017 (PI Jeffrey Kaye, MD), P30 AG010161 (PI David Bennett, MD), P30 AG010129 (PI Charles DeCarli, MD), P50 AG016573 (PI Frank LaFerla, PhD), P50 AG005131 (PI Douglas Galasko, MD), P30 AG028383 (PI Linda Van Eldik, PhD), P30 AG010124 (PI John Trojanowski, MD, PhD), P50 AG005142 (PI Helena Chui, MD), P30 AG012300 (PI Roger Rosenberg, MD), P50 AG005136 (PI Thomas Grabowski, MD), P50 AG005681 (PI John Morris, MD), P30 AG028377 (Kathleen Welsh-Bohmer, PhD), and P50 AG008671 (PI Henry Paulson, MD, PhD).

Samples from the National Cell Repository for Alzheimer’s Disease (NCRAD), which receives government support under a cooperative agreement grant (U24 AG21886) awarded by the National Institute on Aging (NIA), were used in this study. We thank contributors who collected samples used in this study, as well as patients and their families, whose help and participation made this work possible.

The Alzheimer’s Disease Genetics Consortium supported the collection of samples used in this study through National Institute on Aging (NIA) grants U01AG032984 and RC2AG036528.

We acknowledge the generous contributions of the Cache County Memory Study participants. Sequencing for this study was funded by RF1AG054052 (PI: John S.K. Kauwe)

## Acknowledgments for the use of GWAS data distributed by NIAGADS

The NIA Genetics of Alzheimer’s Disease Data Storage Site (NIAGADS) is supported by a collaborative agreement from the National Institute on Aging, U24AG041689.

NG00047: The NIA supported this work through grants U01-AG032984, RC2-AG036528, U01-AG016976 (Dr Kukull); U24 AG026395, U24 AG026390, R01AG037212, R37 AG015473 (Dr Mayeux); K23AG034550 (Dr Reitz); U24-AG021886 (Dr Foroud); R01AG009956, RC2 AG036650 (Dr Hall); UO1 AG06781, UO1 HG004610 (Dr Larson); R01 AG009029 (Dr Farrer); 5R01AG20688 (Dr Fallin); P50 AG005133, AG030653 (Dr Kamboh); R01 AG019085 (Dr Haines); R01 AG1101, R01 AG030146, RC2 AG036650 (Dr Evans); P30AG10161, R01AG15819, R01AG30146, R01AG17917, R01AG15819 (Dr Bennett); R01AG028786 (Dr Manly); R01AG22018, P30AG10161 (Dr Barnes); P50AG16574 (Dr Ertekin-Taner, Dr Graff-Radford), R01 AG032990 (Dr Ertekin-Taner), KL2 RR024151 (Dr Ertekin-Taner); R01 AG027944, R01 AG028786 (Dr Pericak-Vance); P20 MD000546, R01 AG28786-01A1 (Dr Byrd); AG005138 (Dr Buxbaum); P50 AG05681, P01 AG03991, P01 AG026276 (Dr Goate); and P30AG019610, P30AG13846, U01-AG10483, R01CA129769, R01MH080295, R01AG017173, R01AG025259, R01AG33193, P50AG008702, P30AG028377, AG05128, AG025688, P30AG10133, P50AG005146, P50AG005134, P01AG002219, P30AG08051, MO1RR00096, UL1RR029893, P30AG013854, P30AG008017, R01AG026916, R01AG019085, P50AG016582, UL1RR02777, R01AG031581, P30AG010129, P50AG016573, P50AG016575, P50AG016576, P50AG016577, P50AG016570, P50AG005131, P50AG023501, P50AG019724, P30AG028383, P50AG008671, P30AG010124, P50AG005142, P30AG012300, AG010491, AG027944, AG021547, AG019757, P50AG005136 (Alzheimer Disease GeneticsConsortium [ADGC]). We thank Creighton Phelps, Stephen Synder, and Marilyn Miller from the NIA, who are ex-officio members of the ADGC. Support was also provided by the Alzheimer’s Association (IIRG-08-89720 [Dr Farrer] and IIRG-05-14147 [Dr Pericak-Vance]), National Institute of Neurological Disorders and Stroke grant NS39764, National Institute of Mental Health grant MH60451, GlaxoSmithKline, and the Office of Research and Development, Biomedical Laboratory Research Program, US Department of Veterans Affairs Administration. For the ADGC, biological samples and associated phenotypic data used in primary data analyses were stored at principal investigators’ institutions and at the National Cell Repository for Alzheimer’s Disease (NCRAD) at Indiana University, funded by the NIA. Associated phenotypic data used in secondary data analyses were stored at the National Alzheimer’s Coordinating Center and at the NIA Alzheimer’s Disease Data Storage Site at the University of Pennsylvania, funded by the NIA. Contributors to the genetic analysis data included principal investigators on projects ndividually funded by the NIA, other NIH institutes, or private entities.

## Acknowledgments for other GWAS and phenotype data

### NACC

The NACC database is funded by NIA/NIH Grant U01 AG016976. NACC data are contributed by the NIA-funded ADCs: P30 AG019610 (PI Eric Reiman, MD), P30 AG013846 (PI Neil Kowall, MD), P30 AG062428-01 (PI James Leverenz, MD) P50 AG008702 (PI Scott Small, MD), P50 AG025688 (PI Allan Levey, MD, PhD), P50 AG047266 (PI Todd Golde, MD, PhD), P30 AG010133 (PI Andrew Saykin, PsyD), P50 AG005146 (PI Marilyn Albert, PhD), P30 AG062421-01 (PI Bradley Hyman, MD, PhD), P30 AG062422-01 (PI Ronald Petersen, MD, PhD), P50 AG005138 (PI Mary Sano, PhD), P30 AG008051 (PI Thomas Wisniewski, MD), P30 AG013854 (PI Robert Vassar, PhD), P30 AG008017 (PI Jeffrey Kaye, MD), P30 AG010161 (PI David Bennett, MD), P50 AG047366 (PI Victor Henderson, MD, MS), P30 AG010129 (PI Charles DeCarli, MD), P50 AG016573 (PI Frank LaFerla, PhD), P30 AG062429-01(PI James Brewer, MD, PhD), P50 AG023501 (PI Bruce Miller, MD), P30 AG035982 (PI Russell Swerdlow, MD), P30 AG028383 (PI Linda Van Eldik, PhD), P30 AG053760 (PI Henry Paulson, MD, PhD), P30 AG010124 (PI John Trojanowski, MD, PhD), P50 AG005133 (PI Oscar Lopez, MD), P50 AG005142 (PI Helena Chui, MD), P30 AG012300 (PI Roger Rosenberg, MD), P30 AG049638 (PI Suzanne Craft, PhD), P50 AG005136 (PI Thomas Grabowski, MD), P30 AG062715-01 (PI Sanjay Asthana, MD, FRCP), P50 AG005681 (PI John Morris, MD), P50 AG047270 (PI Stephen Strittmatter, MD, PhD).

### MARS & LATC

We thank all Minority Aging Research Study and Latino Core participants and the Rush Alzheimer’s Disease Center staff. This database was funded by the NIH/NIA grants R01AG22018 (MARS) and P30AG 072975 (ADC).

### GenADA

The genotypic and associated phenotypic data used in the study “Multi-Site Collaborative Study for Genotype-Phenotype Associations in Alzheimer’s Disease (GenADA)” were provided by the GlaxoSmithKline, R&D Limited.

### ROSMAP

ROSMAP study data were provided by the Rush Alzheimer’s Disease Center, Rush University Medical Center, Chicago. Data collection was supported through funding by NIA grants P30AG10161, R01AG15819, R01AG17917, R01AG30146, R01AG36836, U01AG32984, U01AG46152, the Illinois Department of Public Health, and the Translational Genomics Research Institute.

### AddNeuroMed

The AddNeuroMed data are from a public-private partnership supported by EFPIA companies and SMEs as part of InnoMed (Innovative Medicines in Europe), an Integrated Project funded by the European Union of the Sixth Framework program priority FP6-2004-LIFESCIHEALTH-5. Clinical leads responsible for data collection are Iwona Kłoszewska (Lodz), Simon Lovestone (London), Patrizia Mecocci (Perugia), Hilkka Soininen (Kuopio), Magda Tsolaki (Thessaloniki), and Bruno Vellas (Toulouse), imaging leads are AndySimmons (London), Lars-Olad Wahlund (Stockholm) and Christian Spenger (Zurich) and bioinformatics leads are Richard Dobson (London) and Stephen Newhouse (London).

### ADNI

Data collection and sharing for this project was funded by the Alzheimer’s Disease Neuroimaging Initiative (ADNI) (National Institutes of Health Grant U01 AG024904) and DOD ADNI (Department of Defense award number W81XWH-12-2-0012). ADNI is funded by the National Institute on Aging, the National Institute of Biomedical Imaging and Bioengineering and through generous contributions from the following: AbbVie. Alzheimer’s Association; Alzheimer’s Drug Discovery Foundation; Araclon Biotech; BioClinica. Inc.; Biogen; Bristol-Myers Squibb Company; CereSpir. Inc.; Cogstate; Eisai Inc.; Elan Pharmaceuticals. Inc.; Eli Lilly and Company; EuroImmun; F. Hoffmann-La Roche Ltd and its affiliated company Genentech. Inc.; Fujirebio; GE HealtControlsare; IXICO Ltd.; Janssen Alzheimer Immunotherapy Research & Development. LLC.; Johnson & Johnson Pharmaceutical Research & Development LLC.; Lumosity; Lundbeck; Merck & Co. Inc.; Meso Scale Diagnostics. LLC.; NeuroRx Research; Neurotrack Technologies; Novartis Pharmaceuticals Corporation; Pfizer Inc.; Piramal Imaging; Servier; Takeda Pharmaceutical Company; and Transition Therapeutics. The Canadian Institutes of Health Research is providing funds to support ADNI clinical sites in Canada. Private sector contributions are facilitated by the Foundation for the National Institutes of Health. The grantee organization is the Northern California Institute for Research and Education, and the study is coordinated by the Alzheimer’s Therapeutic Research Institute at the University of Southern California. ADNI data are disseminated by the Laboratory for Neuro Imaging at the University of Southern California.

### NCRAD

Biological samples used in this study were stored at study investigators’ institutions and at the National Cell Repository for Alzheimer’s Disease (NCRAD) at Indiana University, which receives government support under a cooperative agreement grant (U24 AG21886) awarded by the National Institute on Aging (NIA). We thank contributors who collected samples used in this study, as well as patients and their families, whose help and participation made this work possible.

### UK Biobank

UK Biobank data were analyzed under Application Number 45420.

### VA Million Veteran Program: Core Acknowledgement

### MVP Program Office

- Sumitra Muralidhar, Ph.D., Program Director

US Department of Veterans Affairs, 810 Vermont Avenue NW, Washington, DC 20420

- Jennifer Moser, Ph.D., Associate Director, Scientific Programs

US Department of Veterans Affairs, 810 Vermont Avenue NW, Washington, DC 20420

- Jennifer E. Deen, B.S., Associate Director, Cohort & Public Relations

US Department of Veterans Affairs, 810 Vermont Avenue NW, Washington, DC 20420

### MVP Executive Committee

- Co-Chair: Philip S. Tsao, Ph.D.

VA Palo Alto Health Care System, 3801 Miranda Avenue, Palo Alto, CA 94304

- Co-Chair: Sumitra Muralidhar, Ph.D.

US Department of Veterans Affairs, 810 Vermont Avenue NW, Washington, DC 20420

- J. Michael Gaziano, M.D., M.P.H.

VA Boston Healthcare System, 150 S. Huntington Avenue, Boston, MA 02130

- Elizabeth Hauser, Ph.D.

Durham VA Medical Center, 508 Fulton Street, Durham, NC 27705

- Amy Kilbourne, Ph.D., M.P.H.

VA HSR&D, 2215 Fuller Road, Ann Arbor, MI 48105

- Michael Matheny, M.D., M.S., M.P.H.

VA Tennessee Valley Healthcare System, 1310 24^th^ Ave. South, Nashville, TN 37212

- Dave Oslin, M.D.

Philadelphia VA Medical Center, 3900 Woodland Avenue, Philadelphia, PA 19104

### MVP Co-Principal Investigators

- J. Michael Gaziano, M.D., M.P.H.

VA Boston Healthcare System, 150 S. Huntington Avenue, Boston, MA 02130

- Philip S. Tsao, Ph.D.

VA Palo Alto Health Care System, 3801 Miranda Avenue, Palo Alto, CA 94304

### MVP Core Operations

- Jessica V. Brewer, M.P.H., Director, Cohort Operations

VA Boston Healthcare System, 150 S. Huntington Avenue, Boston, MA 02130

- Mary T. Brophy M.D., M.P.H., Director, Biorepository

VA Boston Healthcare System, 150 S. Huntington Avenue, Boston, MA 02130

- Kelly Cho, M.P.H, Ph.D., Director, MVP Phenomics

VA Boston Healthcare System, 150 S. Huntington Avenue, Boston, MA 02130

- Lori Churby, B.S., Director, Regulatory Affairs

VA Palo Alto Health Care System, 3801 Miranda Avenue, Palo Alto, CA 94304

- Scott L. DuVall, Ph.D., Director, VA Informatics and Computing Infrastructure (VINCI)

VA Salt Lake City Health Care System, 500 Foothill Drive, Salt Lake City, UT 84148

- Saiju Pyarajan Ph.D., Director, Data and Computational Sciences

VA Boston Healthcare System, 150 S. Huntington Avenue, Boston, MA 02130

- Luis E. Selva, Ph.D., Director, MVP Biorepository Coordination

VA Boston Healthcare System, 150 S. Huntington Avenue, Boston, MA 02130

- Shahpoor (Alex) Shayan, M.S., Director, MVP PRE Informatics

VA Boston Healthcare System, 150 S. Huntington Avenue, Boston, MA 02130

- Stacey B. Whitbourne, Ph.D., Director, MVP Cohort Development and Management

VA Boston Healthcare System, 150 S. Huntington Avenue, Boston, MA 02130

- MVP Coordinating Centers

`o` MVP Coordinating Center, Boston - J. Michael Gaziano, M.D., M.P.H.

VA Boston Healthcare System, 150 S. Huntington Avenue, Boston, MA 02130

`o` MVP Coordinating Center, Palo Alto – Philip S. Tsao, Ph.D.

VA Palo Alto Health Care System, 3801 Miranda Avenue, Palo Alto, CA 94304

`o` MVP Information Center, Canandaigua – Brady Stephens, M.S.

Canandaigua VA Medical Center, 400 Fort Hill Avenue, Canandaigua, NY 14424

`o` Cooperative Studies Program Clinical Research Pharmacy Coordinating Center, Albuquerque – Todd Connor, Pharm.D.; Dean P. Argyres, B.S., M.S.

New Mexico VA Health Care System, 1501 San Pedro Drive SE, Albuquerque, NM 87108

## MVP Publications and Presentations Committee

- Co-Chair: Themistocles L. Assimes, M.D., Ph. D

VA Palo Alto Health Care System, 3801 Miranda Avenue, Palo Alto, CA 94304

- Co-Chair: Adriana Hung, M.D.; M.P.H

VA Tennessee Valley Healthcare System, 1310 24^th^ Ave. South, Nashville, TN 37212

- Co-Chair: Henry Kranzler, M.D.

Philadelphia VA Medical Center, 3900 Woodland Avenue, Philadelphia, PA 19104

## MVP Local Site Investigators

- Samuel Aguayo, M.D., Phoenix VA Health Care System

650 E. Indian School Road, Phoenix, AZ 85012

- Sunil Ahuja, M.D., South Texas Veterans Health Care System

7400 Merton Minter Boulevard, San Antonio, TX 78229

- Kathrina Alexander, M.D., Veterans Health Care System of the Ozarks

1100 North College Avenue, Fayetteville, AR 72703

- Xiao M. Androulakis, M.D., Columbia VA Health Care System

6439 Garners Ferry Road, Columbia, SC 29209

- Prakash Balasubramanian, M.D., William S. Middleton Memorial Veterans Hospital

2500 Overlook Terrace, Madison, WI 53705

- Zuhair Ballas, M.D., Iowa City VA Health Care System

601 Highway 6 West, Iowa City, IA 52246-2208

- Elizabeth S. Bast, M.D., M.P.H., Miami VA Health Care System

1201 NW 16th Street, 11 GRC, Miami FL 33125

- Jean Beckham, Ph.D., Durham VA Medical Center

508 Fulton Street, Durham, NC 27705

- Sujata Bhushan, M.D., VA North Texas Health Care System

4500 S. Lancaster Road, Dallas, TX 75216

- Edward Boyko, M.D., VA Puget Sound Health Care System

1660 S. Columbian Way, Seattle, WA 98108-1597

- David Cohen, M.D., Portland VA Medical Center

3710 SW U.S. Veterans Hospital Road, Portland, OR 97239

- Louis Dellitalia, M.D., Birmingham VA Medical Center

700 S. 19th Street, Birmingham AL 35233

- Gerald Wayne Dryden, Jr., M.D., Ph.D., Louisville VA Medical Center

800 Zorn Avenue, Louisville, KY 40206

- L. Christine Faulk, M.D., Robert J. Dole VA Medical Center

5500 East Kellogg Drive, Wichita, KS 67218-1607

- Joseph Fayad, M.D., VA Southern Nevada Healthcare System

6900 North Pecos Road, North Las Vegas, NV 89086

- Daryl Fujii, Ph.D., VA Pacific Islands Health Care System

459 Patterson Rd, Honolulu, HI 96819

- Saib Gappy, M.D., John D. Dingell VA Medical Center

4646 John R Street, Detroit, MI 48201

- Frank Gesek, Ph.D., White River Junction VA Medical Center

163 Veterans Drive, White River Junction, VT 05009

- Michael Godschalk, M.D., Richmond VA Medical Center

1201 Broad Rock Blvd., Richmond, VA 23249

- Jennifer Greco, M.D., Sioux Falls VA Health Care System

2501 W 22nd Street, Sioux Falls, SD 57105

- Todd W. Gress, M.D., Ph.D., Hershel “Woody” Williams VA Medical Center

1540 Spring Valley Drive, Huntington, WV 25704

- Samir Gupta, M.D., M.S.C.S., VA San Diego Healthcare System

3350 La Jolla Village Drive, San Diego, CA 92161

- Salvador Gutierrez, M.D., Edward Hines, Jr. VA Medical Center

5000 South 5th Avenue, Hines, IL 60141

- Mark Hamner, M.D., Ralph H. Johnson VA Medical Center

109 Bee Street, Mental Health Research, Charleston, SC 29401

- John Harley, M.D., Ph.D., Cincinnati VA Medical Center

3200 Vine Street, Cincinnati, OH 45220

- Daniel J. Hogan, M.D., Bay Pines VA Healthcare System

10,000 Bay Pines Blvd Bay Pines, FL 33744

- Adriana Hung, M.D., M.P.H., VA Tennessee Valley Healthcare System

1310 24th Avenue, South Nashville, TN 37212

- Robin Hurley, M.D., W.G. (Bill) Hefner VA Medical Center

1601 Brenner Ave, Salisbury, NC 28144

- Pran Iruvanti, D.O., Ph.D., Hampton VA Medical Center

100 Emancipation Drive, Hampton, VA 23667

- Frank Jacono, M.D., VA Northeast Ohio Healthcare System

10701 East Boulevard, Cleveland, OH 44106

- Darshana Jhala, M.D., Philadelphia VA Medical Center

3900 Woodland Avenue, Philadelphia, PA 19104

- Seema Joshi, M.D., F.A.C.P., ABOIM; VA Eastern Kansas Health Care System

4101 S 4th Street Trafficway, Leavenworth, KS 66048

- Scott Kinlay, M.B.B.S., Ph.D., VA Boston Healthcare System

150 S. Huntington Avenue, Boston, MA 02130

- Michael Landry, Ph.D., Southeast Louisiana Veterans Health Care System

2400 Canal Street, New Orleans, LA 70119

- Peter Liang, M.D., M.P.H., VA New York Harbor Healthcare System

423 East 23rd Street, New York, NY 10010

- Suthat Liangpunsakul, M.D., M.P.H., Richard Roudebush VA Medical Center

1481 West 10th Street, Indianapolis, IN 46202

- Jack Lichy, M.D., Ph.D., Washington DC VA Medical Center

50 Irving St, Washington, D. C. 20422

- Tze Shien Lo, M.D., Fargo VA Health Care System

2101 N. Elm, Fargo, ND 58102

- C. Scott Mahan, M.D., Charles George VA Medical Center

1100 Tunnel Road, Asheville, NC 28805

- Ronnie Marrache, M.D., VA Maine Healthcare System Center, Augusta, ME 04330
- Stephen Mastorides, M.D., James A. Haley Veterans’ Hospital

13000 Bruce B. Downs Blvd, Tampa, FL 33612

- Kristin Mattocks, Ph.D., M.P.H., Central Western Massachusetts Healthcare System

421 North Main Street, Leeds, MA 01053

- Paul Meyer, M.D., Ph.D., Southern Arizona VA Health Care System

3601 S 6th Avenue, Tucson, AZ 85723

- Jonathan Moorman, M.D., Ph.D., James H. Quillen VA Medical Center

Corner of Lamont & Veterans Way, Mountain Home, TN 37684

- Providencia Morales, R.N., Northern Arizona VA Health Care System

500 Highway 89 North, Prescott, AZ 86313

- Timothy Morgan, M.D., VA Long Beach Healthcare System

5901 East 7th Street Long Beach, CA 90822

- Maureen Murdoch, M.D., M.P.H., Minneapolis VA Health Care System

One Veterans Drive, Minneapolis, MN 55417

- Eknath Naik, M.D., Ph.D., West Palm Beach VA Medical Center,

7305 North Military Trail, West Palm Beach, FL 33410-6400

- James Norton, Ph.D., VA Health Care Upstate New York

113 Holland Avenue, Albany, NY 12208

- Olaoluwa Okusaga, M.D., Michael E. DeBakey VA Medical Center

2002 Holcombe Blvd, Houston, TX 77030

- Michael K. Ong, M.D., VA Greater Los Angeles Health Care System

11301 Wilshire Blvd, Los Angeles, CA 90073

- Kris Ann Oursler, M.D., Salem VA Medical Center

1970 Roanoke Blvd, Salem, VA 24153

- Ismene Petrakis, M.D., VA Connecticut Healthcare System

950 Campbell Avenue, West Haven, CT 06516

- Samuel Poon, M.D., Manchester VA Medical Center

718 Smyth Road, Manchester, NH 03104

- Amneet S. Rai, Pharm.D., VA Sierra Nevada Health Care System

975 Kirman Avenue, Reno, NV 89502

- Michael Rauchman, M.D., St. Louis VA Health Care System

915 North Grand Blvd, St. Louis, MO 63106

- Richard Servatius, Ph.D., Syracuse VA Medical Cente

800 Irving Avenue, Syracuse, NY 13210

- Satish Sharma, M.D., Providence VA Medical Center

830 Chalkstone Avenue, Providence, RI 02908

- River Smith, Ph.D., Eastern Oklahoma VA Health Care System

1011 Honor Heights Drive, Muskogee, OK 74401

- Peruvemba Sriram, M.D., N. FL/S. GA Veterans Health System

1601 SW Archer Road, Gainesville, FL 32608

- Patrick Strollo, Jr., M.D., VA Pittsburgh Health Care System

University Drive, Pittsburgh, PA 15240

- Neeraj Tandon, M.D., Overton Brooks VA Medical Center

510 East Stoner Ave, Shreveport, LA 71101

- Philip Tsao, Ph.D., VA Palo Alto Health Care System

3801 Miranda Avenue, Palo Alto, CA 94304-1290

- Gerardo Villareal, M.D., New Mexico VA Health Care System

1501 San Pedro Drive, S.E. Albuquerque, NM 87108

- Jessica Walsh, M.D., VA Salt Lake City Health Care System

500 Foothill Drive, Salt Lake City, UT 84148

- John Wells, Ph.D., Edith Nourse Rogers Memorial Veterans Hospital

200 Springs Road, Bedford, MA 01730

- Jeffrey Whittle, M.D., M.P.H., Clement J. Zablocki VA Medical Center

5000 West National Avenue, Milwaukee, WI 53295

- Mary Whooley, M.D., San Francisco VA Health Care System

4150 Clement Street, San Francisco, CA 94121

- Peter Wilson, M.D., Atlanta VA Medical Center

1670 Clairmont Road, Decatur, GA 30033

- Junzhe Xu, M.D., VA Western New York Healthcare System

3495 Bailey Avenue, Buffalo, NY 14215-1199

- Shing Shing Yeh, Ph.D., M.D., Northport VA Medical Center

79 Middleville Road, Northport, NY 11768

- Andrew W. Yen, M.D., VA Northern California Health Care System

10535 Hospital Way, Mather, CA 95655

## References

1. Sun L, Wang Z, Lu T, Manolio TA, Paterson AD. eXclusionarY: 10 years later, where are the sex chromosomes in GWASs? The American Journal of Human Genetics. 2023;110(6):903–912. doi:10.1016/j.ajhg.2023.04.009

2. Alzheimer’s Association. Alzheimer’s disease facts and figures. Alzheimer’s and Dementia. 2023;19(4):1598-1695. doi:10.1002/alz.13016

3. NIAGADS. NG00067 – ADSP Umbrella. Published 2021. Accessed November 1, 2021. https://dss.niagads.org/datasets/ng00067/

4. Kuzma A, Valladares O, Cweibel R, et al. NIAGADS: The NIA Genetics of Alzheimer’s Disease Data Storage Site. Alzheimer’s & Dementia. 2016;12(11):1200–1203. doi:10.1016/j.jalz.2016.08.018

5. Bycroft C, Freeman C, Petkova D, et al. The UK Biobank resource with deep phenotyping and genomic data. Nature. 2018;562:203–209. doi:10.1038/s41586-018-0579-z

6. Kurki MI, Karjalainen J, Palta P, et al. FinnGen provides genetic insights from a well-phenotyped isolated population. Nature. 2023;613(7944):508–518. doi:10.1038/S41586-022-05473-8

7. Sherva R, Zhang R, Sahelijo N, et al. African ancestry GWAS of dementia in a large military cohort identifies significant risk loci. Mol Psychiatry. 2023;28(3):1293–1302. doi:10.1038/s41380-022-01890-3

8. Hunter-Zinck H, Shi Y, Li M, et al. Genotyping Array Design and Data Quality Control in the Million Veteran Program. The American Journal of Human Genetics. 2020;106(4):535–548. doi:10.1016/J.AJHG.2020.03.004

9. Le Guen Y, Napolioni V, Belloy ME, et al. Common X-Chromosome Variants Are Associated with Parkinson Disease Risk. Ann Neurol. 2021;90(1):22–34. doi:10.1002/ana.26051

10. Jansen IE, Savage JE, Watanabe K, et al. Genome-wide meta-analysis identifies new loci and functional pathways influencing Alzheimer’s disease risk. Nat Genet. 2019;51(3):404–413. doi:10.1038/s41588-018-0311-9

11. Loh P ru, Kichaev G, Gazal S, Schoech AP, Price AL. Mixed-model association for biobank-scale datasets. Nat Genet. 2018;50:906–908. doi:10.1038/s41588-018-0144-6

12. Sidorenko J, Kassam I, Kemper KE, et al. The effect of X-linked dosage compensation on complex trait variation. Nat Commun. 2019;10(1). doi:10.1038/s41467-019-10598-y

13. Giambartolomei C, Vukcevic D, Schadt EE, et al. Bayesian Test for Colocalisation between Pairs of Genetic Association Studies Using Summary Statistics. PLoS Genet. 2014;10(5):1–15. doi:10.1371/journal.pgen.1004383

14. Andrews SJ, Renton AE, Fulton-Howard B, Podlesny-Drabiniok A, Marcora E, Goate AM. The complex genetic architecture of Alzheimer’s disease: novel insights and future directions. EBioMedicine. 2023;90. doi:10.1016/J.EBIOM.2023.104511

15. Stopschinski BE, Holmes BB, Miller GM, et al. Specific glycosaminoglycan chain length and sulfation patterns are required for cell uptake of tau versus alfa- synuclein and beta-amyloid aggregates. Journal of Biological Chemistry. 2018;293(27):10826–10840. doi:10.1074/jbc.RA117.000378

16. Pohlkamp T, Xian X, Wong CH, et al. NHE6 depletion corrects ApoE4-mediated synaptic impairments and reduces Amyloid plaque load. Elife. 2021;10. doi:10.7554/eLife.72034

