## Appendix-A for "The Role of X Chromosome in Alzheimer’s Disease Genetics"

The need to maintain the pH of the cytoplasm and intracellular compartments within close parameters is essential to the functioning of the cell and the regulation of intracellular transport mechanisms. Many if not most of the biological functions of the cell have evolved to take place at or near a narrowly defined pH optimum and any deviation from this optimum results in progressively impaired functions and ultimately death of the cell and the organism. Enzymes have evolved to function at a pH optimum that resides usually around 7.4 in the cytoplasm. However, enzymes like lysosomal proteases or lipid metabolic enzymes have evolved to be most active at an acidic pH that is closer to 5. Indeed, activation of these enzymes often occurs only once they have progressed into heavily acidified compartments in order to prevent their premature activation in the biosynthetic or secretory pathway where they could prematurely degrade newly synthesized molecules. The acidification of these intracellular organelles is mediated by the vacuolar ATPase (vATPase) a.k.a. the proton pump. This is an energy intensive process by which the vATPase establishes an electrochemical proton gradient across diverse cellular membranes which not only regulates the activity and rate of catalytic events but also drives the appropriate sorting of intracellular cargo-containing compartments (reviewed in (Vasanthakumar and Rubinstein, 2020). For instance, alkalinization of intracellular compartments of the Golgie and the endolysosomal pathway with weak bases like ammonium chloride or chloroquine prevents or delays the processing of endocytosed cargo and the timely recycling of endocytic receptors from early endosomes (Thorens and Vassalli, 1986). Therefore, the kinetics at which acidification of specific compartments occurs is of the utmost importance for the coordinated and regulated functioning of the cell.

The electrochemical gradient that is generated by the vATPase also drives other ion exchange mechanisms that are themselves not dependent upon the consumption of ATP. Such mechanisms involve for instance the chloride/proton antiporter, chloride channels and the sodium hydrogen exchangers (NHEs) (Jentsch and Pusch, 2018; Flessner and Orlowski, 2021). The concerted functions of these ion exchangers in conjunction with the vATPase then regulate the ultimate pH and ion composition of the respective compartments in which they reside.

Nine different sodium hydrogen antiporters regulate pH homeostasis in virtually all tissues of the body in this manner (Flessner and Orlowski, 2021). While NHE 1-4 are primarily involved in ion transport at the plasma membrane, NHE 5 to 9 primarily regulate proton, sodium and potassium homeostasis across intracellular membranes. NHE5 and NHE6 control proton exchange in recycling and early endosomes, while NHE9 has been localized to late endosomes, phagosomes and recycling endosomes. NHE8 appears to be required for perinuclear vesicle fusion and sorting. NHE7, by contrast, localizes to Golgi compartments where it has been implicated in the exchange of protons with the cytoplasm and the acidification of secretory compartments, although its precise functions there require further investigations.

Human genetic defects in *NHE6*, *NHE7* and *NHE9* have been described and found to cause neurodevelopmental syndromes that include autism, epilepsy, mental disability and selective neuronal loss (Flessner and Orlowski, 2021). These findings emphasize the importance of intracellular compartmental pH and ion homeostasis especially for the development and function of the human brain. The critical importance of early endosomal pH regulation is underscored by loss of function mutations in the X-chromosomal *NHE6*, which is the cause for Christianson syndrome. The resulting accelerated and unchecked acidification of the early endosomal compartment results in the premature activation of lysosomal enzymes which in turn mediate the aberrant degradation of various neuronal proteins including neurotrophins and neurotrophin receptors (Ouyang et al., 2013).

At the other end of the spectrum, i.e. in the aging brain, reduced metabolism and impaired energy production would be predicted to adversely impact vATPase function and consequently result in the delayed acidification of the endolysosomal compartment, leading to impaired autophagy and lysosomal degradation of cellular waste products, including amyloid and tau. Such a model of AD pathogenesis is supported by numerous studies in mice and humans that have revealed profound impairments of vATPase-mediated proton translocation activity in mouse models of AD (Lee et al., 2022) as well as the prominent enlargement of endosomes that has been proposed to be the result of impaired endosomal acidification kinetics (Pohlkamp et al., 2021). Consequently, genetic disruption of *NHE6*, the primary proton leak channel in the early endosome, was found to greatly delay amyloid accumulation in an AD mouse model (Pohlkamp et al., 2021). Similarly, pharmacological or genetic inhibition of NHE6 function in cortical neurons completely abolished the endosomal recycling delay and intracellular sequestration of ApoE and excitatory neurotransmitter receptors in the presence of ApoE4 (Xian et al., 2018). Conversely, missense mutations in *NHE7* lead to Golgi alkalinization (Khayat et al., 2019) where NHE7 has been proposed to mediate proton influx from the cytosol in exchange for sodium ions (Milosavljevic et al., 2014). Consistent with the proposed models and the conclusions of these earlier studies, our current finding now further suggests that a genetic polymorphism that results in a modest increase of NHE7 protein expression is neurodevelopmentally neutral, but by disrupting Golgi pH homeostasis also appears to increase risk for late-onset AD. This finding therefore supports proposed therapeutic interventions where partial pharmacological inhibition of NHEs, i.e. intracellular proton leak channels, during aging to support ailing proton pump activity might stave off or prevent the manifestation of AD (Xian et al., 2018; Pohlkamp et al., 2021).

### References


Flessner R, Orlowski J (2021) Na+/H+ Exchangers. In: Encyclopedia of Molecular Pharmacology (Offermanns S, Rosenthal W, eds), pp 1047-1062. Cham: Springer International Publishing.

Jentsch TJ, Pusch M (2018) CLC Chloride Channels and Transporters: Structure, Function, Physiology, and Disease. Physiol Rev 98:1493-1590.

Khayat W, Hackett A, Shaw M, Ilie A, Dudding-Byth T, Kalscheuer VM, Christie L, Corbett MA, Juusola J, Friend KL, Kirmse BM, Gecz J, Field M, Orlowski J (2019) A recurrent missense variant in SLC9A7 causes nonsyndromic X-linked intellectual disability with alteration of Golgi acidification and aberrant glycosylation. Hum Mol Genet 28:598-614.

Lee JH, Yang DS, Goulbourne CN, Im E, Stavrides P, Pensalfini A, Chan H, Bouchet-Marquis C, Bleiwas C, Berg MJ, Huo C, Peddy J, Pawlik M, Levy E, Rao M, Staufenbiel M, Nixon RA (2022) Faulty autolysosome acidification in Alzheimer's disease mouse models induces autophagic build-up of Abeta in neurons, yielding senile plaques. Nat Neurosci 25:688-701.

Milosavljevic N, Monet M, Lena I, Brau F, Lacas-Gervais S, Feliciangeli S, Counillon L, Poet M (2014) The intracellular Na(+)/H(+) exchanger NHE7 effects a Na(+)-coupled, but not K(+)-coupled proton-loading mechanism in endocytosis. Cell Rep 7:689-696.

Ouyang Q, Lizarraga SB, Schmidt M, Yang U, Gong J, Ellisor D, Kauer JA, Morrow EM (2013) Christianson syndrome protein NHE6 modulates TrkB endosomal signaling required for neuronal circuit development. Neuron 80:97-112.

Pohlkamp T, Xian X, Wong CH, Durakoglugil MS, Werthmann GC, Saido TC, Evers BM, White CL, 3rd, Connor J, Hammer RE, Herz J (2021) NHE6 depletion corrects ApoE4-mediated synaptic impairments and reduces amyloid plaque load. Elife 10.

Thorens B, Vassalli P (1986) Chloroquine and ammonium chloride prevent terminal glycosylation of immunoglobulins in plasma cells without affecting secretion. Nature 321:618-620.

Vasanthakumar T, Rubinstein JL (2020) Structure and Roles of V-type ATPases. Trends Biochem Sci 45:295-307.

Xian X, Pohlkamp T, Durakoglugil MS, Wong CH, Beck JK, Lane-Donovan C, Plattner F, Herz J (2018) Reversal of ApoE4-induced recycling block as a novel prevention approach for Alzheimer's disease. Elife 7.
