## Supplement for "The Role of X Chromosome in Alzheimer’s Disease Genetics"

### eMethods

In the current study, we used data from a variety of cohorts and sequencing projects related to AD[1–23]. All available genetic/phenotypic data were jointly harmonized with the purpose of performing phenotype/covariate harmonization. Details are provided below.

#### ADGC & ADSP Phenotype Ascertainment

*Cohorts and Phenotype Ascertainment*

Details on phenotype ascertainment are described elsewhere[1–5,7]. Briefly, all individuals with a diagnosis of AD met National Institute of Neurological and Communicative Disorders and Stroke/Alzheimer's Disease and Related Disorders Association (NINCDS-ADRDA) criteria for definite, probable, or possible late-onset AD[6], or met Diagnosis and Statistical Manual of Mental Disorders IV-V (DSMIV-V) criteria[8–10], or had a clinical dementia rating (CDR® Dementia Staging Instrument[11]) > 0.5. Some cohorts verified AD diagnoses through neuropathology, using Braak staging[12], CERAD scoring[22], or National Institute on Aging Reagan (NIA-Reagan) 1997 criteria[13]. Cognitively normal subjects did not have AD according to the above clinical AD criteria, did not have a diagnosis of mild-cognitive impairment (MCI), and had a CDR of 0 and/or Mini-Mental State Examination (MMSE[14]) > 25. In MIRAGE, control status was evaluated through a Modified Telephone Interview of Cognitive Status score ≥ 86 (a telephone version of the MMSE)[15].

Further, the National Alzheimer's Coordinating Center (NACC), Rush University Religious Orders Study/Memory and Aging Project (ROSMAP), and Alzheimer's Disease Neuroimaging Initiative (ADNI), are longitudinal cohorts that provide detailed information regarding clinical status (control, MCI, demented) and presumed disease etiology at repeated examinations. Additionally, deceased subjects are assessed for neuropathology. Where possible, in NACC, a final diagnosis of MCI or possible/probable/definite AD was obtained using NIA Alzheimer's Association (NIA-AA) 2011 criteria[16,17]. In all three cohorts, AD diagnoses were verified by neuropathology as middle or high AD likelihood following NIA-Reagan 1997 criteria (moderate to frequent neuritic plaques and Braak stage III-VI)[13]. In concordance with the category "possible AD dementia with evidence of the AD pathophysiological process" from the NIA-AA 2011 criteria[16], we attributed possible AD diagnoses to subjects who met clinical criteria for non-AD dementia but also met AD neuropathological criteria. In concordance with the NIA-AA 2011/2012 framework[17,18], we also evaluated neuropathology in MCI subjects to verify presumed AD etiology. Controls were not re-evaluated based on neuropathology data. Subjects that reverted from dementia to control status during longitudinal follow-up were excluded. Additional cohort-specific details are listed below.

*NACC*

Genotyping waves 1 through 7 from the Alzheimer's Disease Centers (ADC1-7) and a subset of the ADSP projects include subjects ascertained and evaluated by the clinical and neuropathological cores of 32 NIA-funded ADCs. NACC coordinates the collection of these phenotypes, implements diagnoses (cognitively normal, cognitively impaired but not MCI, MCI, demented; and presumed disease etiology), and then provides all data to researchers under the form of the Minimum Data Set (MDS), Uniform Data Set (UDS)[19,20,23], and Neuropathology data set (NP)[21]. The MDS represents an older subset of the NACC data and only contains cross-sectional data, while the more recent UDS provides longitudinal phenotypes and covariates. Since 2015, the UDS was updated to incorporate the NIA-AA 2011 criteria for MCI and AD[17,24]. In the current study, we used the UDS and NP for which data was collected between September 2005 and March 2022, to determine phenotypes for subjects in ADC1-7, ADSP WES/WGS, and ADGC Exome arrays.

Subjects that had a diagnosis of Down syndrome, central nervous system neoplasm, bipolar disorder, schizophrenia, alcohol-induced dementia, or substance-abuse-induced dementia, were excluded. Subjects carrying mutations of dominantly inherited AD or frontotemporal lobar degeneration (FTLD) were also excluded. Subjects with a final diagnosis of MCI or dementia, for which the etiology was unknown, not due to AD, or only secondary due to AD (and without AD neuropathological information), were excluded. Subjects with a final diagnosis of "cognitively impaired but not MCI", but having no other neurological disorder, were kept as controls, considering that this more consistently matched control criteria in many of the other cohorts considered in this study.

*ROSMAP*

In ROSMAP, subjects were diagnosed at each visit: as possible/probable AD according to NINCDS-ADRDA criteria[6]; as MCI when judged to have cognitive impairment but not meeting dementia criteria according to the clinician; or as control when there was no cognitive impairment or the subject did not meet dementia criteria[25,26]. At time of death, a final clinical diagnosis was made by an expert neurologist, followed by a case conference consensus review (blinded to postmortem data)[27].

*ADNI*

In ADNI, subjects were diagnosed at regular visits: as possible/probable AD according to NINCDS-ADRDA criteria[6]; as MCI according to Petersen/Winblad criteria; or as control when not demented, not MCI, CDR = 0, and MMSE > 28. Neuropathology assessments followed the NACC NP framework.

*Phenotype Harmonization*

The available sample contained many subjects that were genotyped multiple times across different studies. This largely reflected efforts from the ADGC, ADSP, and AMP-AD, to perform next-generation sequencing (NGS) on existing cohort samples for the purpose of rare variant discovery and AD gene prioritization. In other instances, participants were recruited in different studies at different times. Therefore, to handle potential duplicate discordance and phenotype heterogeneity, we implemented a cross-sample phenotype harmonization procedure aiming to standardize pathology-verified diagnoses where possible, share unique missing information across all duplicate entries of a given subject, resolve longitudinal changes in diagnosis, and flag subjects with unresolvable duplicate discordance for exclusion.

Duplicate samples were identified by determining genetic cryptic relatedness (cf. below), but for sample cross-referencing did not include known identical twins in LOAD and ROSMAP samples. First, duplicate samples were flagged as discordant if their age-at-death information differed by more than 2 years or if pathology measures (Braak or neuritic plaque density) differed. Across all cohorts, where possible, AD diagnoses were verified by neuropathology as middle or high AD likelihood following NIA-Reagan 1997 criteria (moderate to frequent neuritic plaques and Braak stage III-VI)[13]. Additionally, when only either neuritic plaque or Braak information was available and in line with NIA-Reagan 1997 middle or high AD likelihood criteria, and/or the cohort/project demographics provided a diagnosis of definite AD, the subject was considered to have pathology-verified AD status. Cognitively normal (CN) subjects with evidence of AD pathology were kept as CN. Further, if at least one entry across duplicate samples indicated a diagnosis of Down syndrome, central nervous system neoplasm, bipolar disorder, schizophrenia, alcohol-induced dementia, substance-abuse-induced dementia, neurological (not including Parkinson's disease), or systemic disease despite being cognitively normal, or carrying mutations of dominantly inherited AD or frontotemporal lobar degeneration (FTLD), then all duplicate samples were marked as such and flagged for exclusion. Extending on the above, all genetic samples were checked for the presence of known pathogenic mutations on *APP, PSEN1, PSEN2,* and *MAPT*, whereby carriers and their duplicate samples were flagged for exclusion.

Then, duplicate samples with differing age entries (i.e. longitudinal changes) were evaluated. Reversions from AD or dementia to MCI status, or from MCI to cognitively normal (CN) status, were permitted, but reversions from AD or non-AD dementia to CN status were flagged for exclusion. "Reversions" from AD to non-AD dementia status were permitted, unless pathology (cf. above) indicated the presence of AD pathology, thereby marking the subject as AD. Vice versa, "conversions" from non-AD dementia to AD status were permitted, unless pathology (cf. above) indicated no presence of AD pathology, thereby marking the subject as non-AD dementia. All other types of conversions were directly permitted. Then, duplicate samples for which the diagnoses at the oldest shared age entries differed, or for which diagnoses differed but age was consistent (i.e. apparent cross-sectional discordances), were evaluated. Discordances between AD and non-AD dementia status were resolved based on pathology (cf. above) or flagged as discordant if no pathology data was available. Discordances between CN and AD status, or CN and non-AD dementia status, were resolved as respectively AD or non-AD dementia when those dementia diagnoses corresponded to a unique age-at-onset (of symptoms) without other available age information (i.e. indicating that a conversion likely occurred after the subject was lost to follow-up in the cohort that last observed a CN status), or, were flagged as discordant if duplicate entries shared the same age-at-examination and age-at-last-exam. Discordances between CN and MCI status, or MCI and AD status, or MCI and non-AD dementia status, were resolved as respectively MCI, AD, or non-AD dementia (i.e. keeping the most severe diagnosis).

Finally, once all clinical diagnostic and pathological data were unified across duplicate entries, pathological criteria were applied once more to obtain the final diagnoses. Where possible, AD diagnoses were verified by neuropathology as middle or high AD likelihood following NIA-Reagan 1997 criteria (moderate to frequent neuritic plaques and Braak stage III-VI)[13]. In concordance with the category "possible AD dementia with evidence of the AD pathophysiological process" from the NIA-AA 2011 criteria[16], we attributed possible AD diagnoses to subjects who met clinical criteria for non-AD dementia but also met AD neuropathological criteria. In concordance with the NIA-AA 2011/2012 framework[17,18], we also evaluated neuropathology in MCI subjects to verify presumed AD etiology and considered subjects as cases if AD pathology, following NIA-Reagan 1997 criteria (cf. above), was present (i.e. marking high likelihood of AD etiology). Controls were not re-evaluated based on neuropathology data.

Beyond cross-referencing clinical diagnostic and pathological data across subjects, other covariates were considered for cross-referencing or sharing in case of missingness across duplicate entries. These included age-at-onset of cognitive symptoms, age-at-examination providing clinical diagnosis, at-at-last exam, age-at-death, sex, race, ethnicity, *APOE* genotype provided from demographics, *APOE* genotype provided from whole-genome sequencing, and *APOE* genotype provided from whole-exome sequencing. Duplicate entries with discordant sex or race information were flagged for exclusion.

**ADGC and ADSP Genetic Data Quality Control and Processing**

*Ascertainment of Genetic Data*

Genotypes were available from high-density single-nucleotide polymorphism (SNP) genotyping microarrays (Illumina or Affymetrix) for ADGC or whole genome sequencing (WGS) for ADSP (**eTable 1-2**). Genotype samples had their genetic variants lifted to hg38 using liftOver if not released in hg38 and annotated using dbSNP153 variant identifiers[28].

*ADGC Autosomal Quality Control*

Autosomal variants were extracted from the SNP array data and further processed in several stages. In each cohort/platform/array, variants were excluded based on genotyping rate (<95%), MAF<1%, and Hardy-Weinberg equilibrium in controls ($p<10^{-6}$) using PLINK v1.9[29]. As in our prior work[30], information derived from the gnomAD v.3.1 database[31] was used to filter out SNPs that met one of the following exclusion criteria: (i) located in a low complexity region, (ii) located within common structural variants (MAF > 1%), (iii) multiallelic SNPs with MAF > 1% for at least two alternate alleles, (iv) located within a common insertion/deletion, (v) having any flag different than PASS in gnomAD, (vi) having potential probe polymorphisms.

*ADSP Autosomal Quality Control*

The ADSP WGS data (NG00067.v5) were joint called by the ADSP following the SNP/Indel Variant Calling Pipeline and data management tool used for the analysis of genome and exome sequencing for the Alzheimer's Disease Sequencing Project (VCPA)[32]. The current analyses of ADSP WGS were restricted to bi-allelic variants, to which we applied the Variant Quality Score Recalibration (VSQR) quality control filter ("PASS" variants; GATK v4.1)[33]. Variants with a genotyping rate less than 80%, deviating from Hardy Weinberg Equilibrium (HWE) in the full sample or in controls ($p<10^{-6}$), and a minor allele count less than 10, were excluded. Consistent with the methodology detailed in Belloy et al. 2022[34], we then applied several filters to remove artifactual variants: (i) variants that represented sequencing center or platform artifacts as identified by Fisher exact testing in controls ($p<10^{-5}$), (ii) variants reported in gnomAD v3.1[31] to have a "non-PASS", falling in a low complexity region, or showing more than 10% allele frequency devation between our European ancestry control participants in ADSP and non-Finnish European participants in gnomAD, and (iii) duplicate discordance variants that show discrepancies across several 100 technical duplicates present in ADSP.

*Genetic Relationship Determination using King*

Across all cohorts, the relatedness of subjects (after QC indicated above) was evaluated through identity-by-descent (IBD) analysis (using directly genotyped non-palindromic SNPs shared across all genetic datasets with a call rate > 95% & minor allele frequency (MAF)>1%)[35]. This outcome was used for duplicate tracking across samples, which in turn was used to enable phenotype harmonization (cf. above).

*Ancestry Determination*

Individual ancestries were determined using SNPweights v.2.1 with populations from the 1000 Genomes Consortium as a reference[36,37]. By applying an ancestry percentage cut-off ≥ 75%, the samples were stratified into the five super populations, South-Asians (SAS), East-Asians (EAS), Amerindians (AMR), Africans (AFR) and Europeans (EUR) (**eFigure 1**). When multiple samples were available for a single unique individual, the ancestry was inferred from the sample with the highest genetic coverage.

*Restriction to European ancestry for XWAS*

XWAS were focused on the European ancestry subsample. Two main reasons explain the more extreme population structure on the X-chromosome compared to autosomes: (i) the X-chromosome has a smaller effective population size and thus the rate of genetic drift of X-linked loci is amplified, (ii) local adaptation will lead to higher levels of differentiation between geographically isolated populations[38]. As such, to better control for population structure, we restricted our analyses to European ancestry participants.

*Relationship Determination and Principal Component Analysis using GENESIS*

For ADGC and ADSP data respectively, the relatedness of subjects and principal components capturing population substructure were determined using IBD and principal component analyses (PCA) as implemented through the R package GENESIS (R v3.6.0)[39]. Specifically, this approach first uses an R-implementation of KING-robust to determine kinship coefficients that take into account ancestry divergence. The derived pairwise kinship coefficients are then used to perform a PCA in related samples (PC-AiR) providing accurate ancestry inference not confounded by family structure. The latter output is then used to estimate kinship coefficients using PC-Relate, which accounts for population structure (ancestry) among sample individuals through the use of ancestry representative principal components (PCs) to provide accurate relatedness estimates due only to recent family (pedigree) structure. For each respective data set, these analyses were performed on pruned SNPs ($R^2 < 0.5$, call rate > 95%, MAF > 1%, and excluding palindromic SNPs) in non-Hispanic White European ancestry individuals.

*ADGC X chromosome Quality Control and TOPMed Imputation*

The X chromosome variants underwent a similar harmonization pipeline as the autosomes. We excluded multi-allelic SNPs, SNPs within structural variations, and potential probe polymorphism SNPs. Additionally, our analysis excluded the pseudoautosomal regions of the X chromosome and used only the European ancestry participants as derived above. Several steps were performed to avoid spurious findings: (i) variants with less than 95% genotyping rate and (ii) individuals with more than 5% genotype missingness were excluded. (iii) Reported sex was checked using PLINK1.9 --*check-sex* flag[29], with 0.4 max value for females and 0.94 min value for men, and all individuals with a discordant sex label were excluded. (iv) Heterozygous SNPs in males were set as missing in males, while (v) SNPs with differential missingness between AD cases and controls were removed ($p<10^{-5}$ per cohort/platform/array). (vi) HWE was tested in female controls and SNPs with $p<10^{-5}$ were removed (per cohort/platform/array). (vii) Any monomorphic SNPs that remained were removed. (viii) Differential missingness and differential MAF between males and females were both tested and SNPs with $p<10^{-5}$, for either one of the tests, were excluded (per cohort/platform/array). Finally, as for the autosomes and based on gnomAD v3.1[31] information, we filtered variants (ix) located in a low complexity region, (x) located within common structural variants (MAF > 1%), (xi) multiallelic SNPs with MAF>1% for at least two alternate alleles, (xii) located within a common insertion/deletion, (xiii) having any flag different than PASS in gnomAD v.3.1, (xiv) having potential probe polymorphisms (xv) more than 10% MAF difference with gnomAD frequency in non-Finnish Eurpeans. The remaining SNPs were checked for consistency with the TOPMed panel, flipping of palindromic SNPs, and were imputed on the TOPMed Imputation server[40,41], which uses Minimac 4 for imputation. The following parameters were selected: reference panel TOPMed-r2 (2022), phasing with Eagle v2.4, r-square imputation score cut off 0.3.

*ADSP X chromosome Quality Control*

The X chromosome variants underwent a similar harmonization pipeline as the autosomes. Our analysis excluded the pseudoautosomal regions of the X chromosome and used only the European ancestry participants. The ADSP WGS data for X chromosome (NG00067.v5) were joint called by the ADSP following the SNP/Indel Variant Calling Pipeline and data management tool used for the analysis of genome and exome sequencing for the Alzheimer's Disease Sequencing Project (VCPA)[32]. The current analyses of ADSP WGS were restricted to bi-allelic variants, to which we applied the Variant Quality Score Recalibration (VSQR) quality control filter ("PASS" variants; GATK v4.1)[33]. Variants with a genotyping rate of less than 80% and a minor allele count of less than 2 were excluded. Consistent with the methodology detailed in

Belloy et al. 2022[34], we then applied several filters to remove artifactual variants: (i) variants that represent sequencing center or platform artifacts as identified by Fisher exact testing in controls ($p<10^{-5}$), (ii) variants reported in gnomAD v3.1 to have a "non-PASS", falling in a low complexity region, or showing more than 10% allele frequency devation between our European ancestry control participants in ADSP and non-Finnish European participants in gnomAD, and (iii) duplicate discordance variants that show discrepancies across several 100 technical duplicates present in ADSP.

Several additional steps were performed to avoid spurious findings: (i) Heterozygous SNPs in males were set as missing in males, (ii) variants with a genotyping rate less than 80% in controls, cases, men, or women were excluded, (iii) variants with differential missingness between AD cases and controls were removed ($p<10^{-10}$ for the full sample). (iv) HWE was tested in female and male controls using the Plink --hardy command that allows joint sex evaluation and variants with $p<10^{-5}$ were removed (for the full sample). (v) Differential missingness between males and females was tested and SNPs with $p<10^{-20}$ were excluded (for the full sample).

**ADGC & ADSP Statistical Analyses**

*Case-control XWAS*

All association analyses with AD risk were adjusted for sex (in non-stratified XWAS), array type, the first 5 genetic prinicipal components (PC-AiRs), and *APOE**4/2 dosage. Age adjustment in case-control analyses was not performed, given that the current AD genetic samples often showed younger ages for cases than controls due to the use of age-at-onset information (**eTable3**), which violates the assumption for age adjustment (which is that older age is associated with increased AD incidence). In prior work, we showed that age adjustment in such scenarios leads to significantly decreased power for genetic association analyses[30]. Adjustment for *APOE* genotypes is relevant given the established interactions with sex[42], which may notably be relevant to the X chromosome and could lead to increased model noise if not accounted for. Additionally, the case-control clinical cohorts are enriched for *APOE**4 cases compared to population-based studies[42], which may further exacerbate any potential confounding effects.

Cohorts from ADGC were pooled into a mega-analysis. LMM-BOLT was used in both ADGC and ADSP[43], using autosomal data to derive genetic relationship matrices to allow the inclusion of related subjects. Resultant betas were converted to traditional odds ratios using the transformation approach as detailed in the LMM-BOLT manual. Across ADGC and ADSP, subjects were unrelated down to 1$^{st}$ degree.

*Age information*

For cases that only had age-at-death (AAD) available, the final ages used for regression analysis were subtracted by 10 years to approximate age-at-onset (AAO). This reflects expected mean delays between AAO and AAD for AD patients[44], and is consistent with the derived age covariate for AD cohorts provided by the Alzheimer's Disease Genetics Consortium (ADGC) on NIAGADS[45]. In cohorts that provide conversion information but not AAO, age-at-examination (AAE) was used and followed a prioritization of age-at-MCI-diagnosis > age-at-dementia-diagnosis (incident) > age-at-dementia diagnosis (prevalent). This was done to most closely approximate AAO. For the remaining control samples, age-at-last-examination (AAL) was used. After implementing these criteria, samples were filtered to have a minimal age of 60 years. Some samples were censored at ages 90+, for which we assumed the age was 90.

**UKB Phenotype ascertainment**

Detailed descriptions of all the variables and field provided by UKB are provided elsewhere[46].

In the first round of phenotype ascertainment, we derived health-registry-confirmed AD status and related age information for the individuals directly. Subjects were assumed to be controls if they had no other diagnosis inferred from health registry information relevant to dementia status. We specifically considered the following data fields and entries: *Diagnoses_main_ICD10* [G300,G301,G308,G309,F000,F001,F002,F009], *Diagnoses_secondary_ICD10* [G300,G301,G308, G309,F000,F001,F002,F009], *Date_of_first_in_patient_diagnosis_main_ICD10* [if date provided], *Date_of_first_in_patient_diagnosis_ICD10* [if date provided], *Source_of_alzheimers_disease_ report* [0,1,11,12,2,21,22 = selft report, hospital admission, death record], *Date_of_alzheimers_disease_ report* [if date provided], *Source_of_all_cause_dementia_report* [0,1,11,12,2,21,22 = selft report, hospital admission, death record], *Source_of_frontotemporal_dementia_report* [any entry], *Source_of_vascular_ dementia_report* [any entry], and *Date_of_all_cause_dementia_report* [if data provided]. The above fields were used to determine dementia status, allowing us to differentiate between late-onset AD individuals (LOAD), early-onset AD (EOAD), vascular dementia, frontotemporal dementia, and other all-cause dementia participants. For the health-registry AD phenotype, cases were restricted to all LOAD individuals. The above fields were further used to determine the earliest available age at which a dementia occurrence or report was made. Age information for controls was available from the variables: *Age_when_attended_assessment_centre* [oldest age entry retrieved] and *Age_at_death*.

We then identified the proxy ADD case and control status and related age by accessing the following fields and entries: *Illnesses_of_father*, *Illnesses_of_mother*, *Illnesses_of_sibblings*, *Fathers_age*, *Fathers_age_at_death*, *Mothers_age*, and *Mothers_age_at_death* (where it should be noted that age and sex info was not available for siblings). The youngest reported age was used for proxy ADD cases, while the oldest reported age was used for proxy controls. Proxy status was ignored if subjects were adopted.

Both health-registry-confirmed AD status and proxy ADD status were then combined into a single phenotype (cf. **eTables6-7**). Notably, all subjects were ages >60y in at least the subject or one parent.

**UKB Genetic Data Quality Control and Processing**

A Detailed description of all the UKB genetic data and processing is provided elsewhere[46]. Specifically, we accessed SNP array data imputed to the Haplotype Reference Consortium (HRC) and UK10K haplotype resource. We further filtered to subjects with consent, passing sex check QC, no heterozygosity outliers, having age information available, and belonging to a white ethnic background (field *Ethnic_background* [1001,1002,1003]). We then identified a homogenous ancestry cluster within this group using "aberrant" on the first 20 genetic PCs, as in Schwartzentruber et al. 2021[47].

**UKB Statistical Analyses**

All association analyses with the AD phenotype were adjusted for sex (in non-stratified XWAS), array type, assessment center, the first 20 genetic principal components provided by UKB, and *APOE**4/2 dosage. LMM-BOLT was used (as was done for ADGC and ADSP)[43], using autosomal data to derive genetic relationship matrices to allow the inclusion of related subjects. Resultant betas were converted to traditional odds ratios using the transformation approach as detailed in the LMM-BOLT manual. Additionally, since the UKB XWAS leveraged the proxy phenotype, an additional correction factor was needed to rescale beta coefficients onto a regular case-control scale. This correction is detailed in **eTables6-7**.

**FinnGen**

Ascertainment of FinnGen phenotype and genotype data is described in detail elsewhere[48]. Summary statistics were available from version 10 (v10) of the publicly released set of genetic summary statistics made available by FinnGen here: https://www.finngen.fi/en/access_results. Documentation on Genetic data processing and statistical analyses is provided here: https://finngen.gitbook.io/documentation. FinnGen made us of Regenie to include related individuals in the genetic association analyses[49]. Information about phenotypes and endpoints is provided here: https://www.finngen.fi/en/researchers/clinical-endpoints, https://r10.risteys.finngen.fi/. We specifically leveraged the "Alzheimer's disease, wide definition" phenotype.

**MVP Phenotype Ascertainment**

Generation of phenotypes for MVP mirrors methods as described in Sherva et al. 2023[50], but was updated based on the more recent MVP 2022_1 data release. Briefly, MVP phenotype data were generated from VA electronic medical records. Alzheimer's disease and related dementia (ADRD) cases were identified on the basis of International Classification of Disease (ICD) 9 and 10 codes. Information on proxy cases (parental dementia) was obtained from the MVP Baseline Survey[51].

**MVP Genetic Data Quality Control and Processing**

Generation and quality control of the MVP genetic data is described in detail elsewhere[52]. Briefly, the genetic data were genotyped using the MVP 1.0 custom Axiom array13, phasing was performed by SHAPEIT4 v 4.1.3, and imputation was performed with MINIMAC4 based on the TopMed imputation reference panel[40]. Variants were then filtered to imputation scores > 0.4 and allele frequencies >= 0.1% in the full dataset. The subset of European-ancestry MVP participants, as determined by the genetically informed Harmonized Ancestry and Race/Ethnicity (HARE) method[53], was then extracted for XWAS and variants were subsequently filtered to allele frequencies >= 0.05%.

**MVP Statistical Analyses**

As in Sherva et al. 2023, our ADRD case-control analysis was independent of the cohort used for the proxy analysis. The MVP case-control XWAS (MVP-1) included ICD-identified cases with onset after age 60. The controls were all over age 65 without any dementia or mild cognitive impairment ICD codes and without a history of AD medication usage. The MVP proxy XWAS (MVP-2) was performed separately using a set of controls without any report of parental dementia. Only survey data for Veteran participants over age 45 at last visit were included in proxy analyses (age for parents was not available). The proxy analyses were focused on maternal phenotypes only. This represents the majority of proxy samples since paternal phenotypes cannot be used for men and there was a relative paucity of women. In both the MVP-1 and MVP-2 cohorts, Plink was used to conduct case-control logistic regression analyses on unrelated subjects. For the proxy XWAS, a correction factor was needed to rescale beta coefficients onto a regular case-control scale. This correction is detailed in **eTables6-7**.

### General Statistical Analyses

*Meta-analysis.*

AD XWAS meta-analyses were conducted using genome-wide, fixed effects inverse-variance weighted meta-analysis as implemented in GWAMA[54].

*Sex heterogeneity.*

Sex heterogeneity tests were evaluated as: Z-value = $(Beta_{men} - Beta_{women})/\sqrt{(SE_{men}^2 + SE_{women}^2)}$. P-values were then determined using the normal distribution with a two-sided hypothesis in R using the following formulation: P-value = 2*pnorm(q=Z-value, lower.tail=FALSE).

*Escape from X chromosome inactivation.*

XCI escape status with regard to AD was evaluated by dividing XWAS beta coefficients from men by beta coefficients from women (similar as in Sidorenko et al. 2019[55]), where a ratio close to 2 suggests no escape from XCI (men beta coefficients for a single active X genotype are double compared to those in women where the X genotype undergoes random XCI) and a ratio close to 1 suggests escape from XCI (the beta coefficients in women become consistent with those in men if there is escape from XCI). We further identified if there was any prior support for XCI at each identified locus by consulting 2 published research articles, containing summaries of genes with prior reported XCI status in addition to novel findings[56,57].

*Genetic Colocalization.*

QTL resources with X chromosome genetic data were available for various tissues from GTEx[58], brain tissue from Wingo et al. 2023[59], and brain tissue (CommonMind, Braineac2), monocytes (CEDAR, Fairfax et al. 2014), microglia (Young et al. 2019), and T cells (Kasela et al. 2017), uniformly processed by the eQTL Catalogue[60]. Colocalization was considered for all genes in each associated locus using a 2Mb window centered on the lead XWAS variant. Evidence for colocalization was considered at colocalization posterior probability (PP4)>0.7 (as in Bellenguez et al. 2022)[61]. Additionally, colocalizations with PP4>0.7 were annotated to indicate whether the QTL passed significane criteria in the respective data/tissues (FDR correction in GTEx and Wingo et al. 2023; P<1e-5 in the eQTL Catalogue which corresponds on average to FDR corrected QTL P-values (no FDR corrected P-values were provided)). Genetic colocalization analyses were restricted to variants seen in 95% of the full XWAS meta-analysis and for which MAF did not deviate >10% across the XWAS and QTL data. We used the "coloc.abf" function from the *coloc* package (R-v.4.2.1)[62], providing sample size, P-value, and MAF. We did not use beta coefficients and standard errors due to potential concerns for variants with MAF close to 50%.

### eFigures.

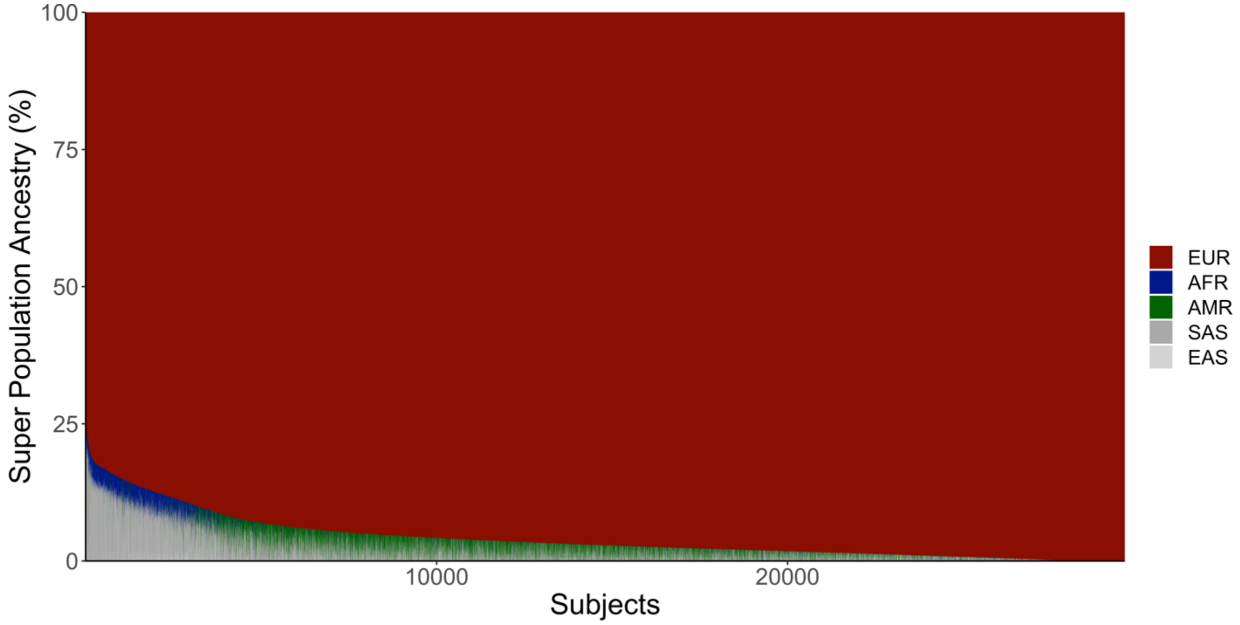

**eFigure 1. Admixture plot across the five major super populations, for European ancestry case-control participants included in ADGC and ADSP.**

*Abbreviations: EUR, European; AFR, African; AMR; Amerindian; SAS, South Asian; EAS; East Asian.*

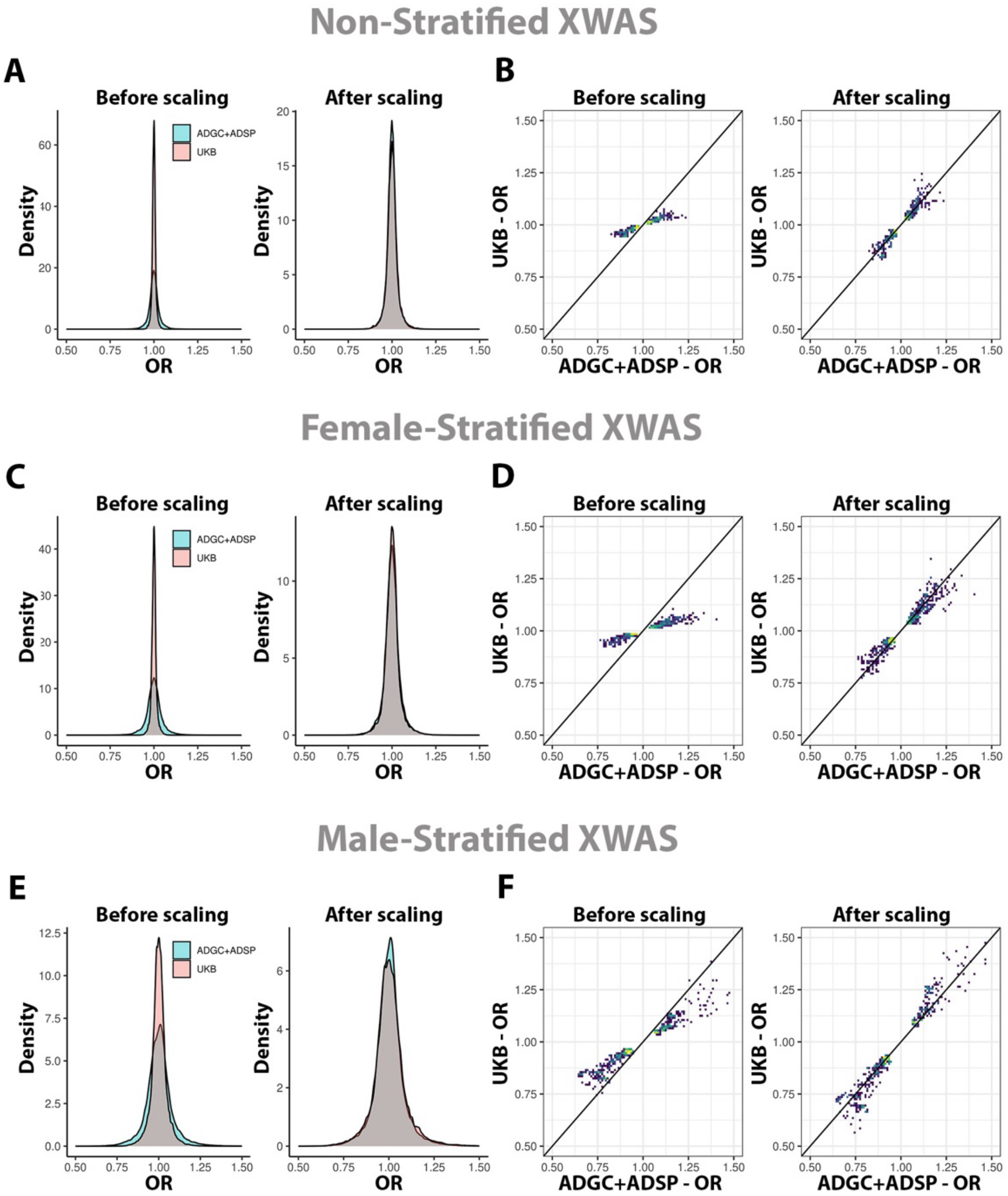

**eFigure 2. UKB beta coefficient adjustment onto a regular case-control scale.** Rescaling for (**A-B**) non-stratified, (**C-D**) female-stratified, and (**E-F**) male-stratified AD XWAS. **A,C,E)** Density plots show beta coefficients for all variants intersecting across ADGC+ADSP and UKB, before and after rescaling. **B,D,F)** Scatter density plots show beta coefficients for prioritized variants intersecting across ADGC+ADSP and UKB, before and after rescaling. Intensity increases from dark blue to bright yellow. A line with slope=1 is plotted for reference. Variants had allele frequencies >= 1%, P<0.1 in both ADGC+ADSP and UKB, concordant effect directions across ADGC+ADSP and UKB, and P<0.01 in ADGC+ADSP+UKB (these variants are more likely to include true associations).

*Abbreviations: OR; Odds Ratio.*

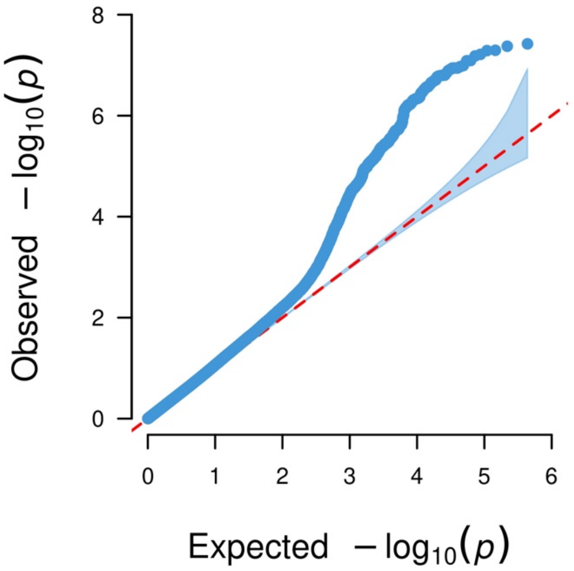

**eFigure 3. QQplot for the non-stratified AD XWAS meta-analysis**. The inflation factor (λ=1.0671) and sample size-adjusted inflation factor ($\lambda_{1,000}$=1.0003) showed no sign of inflation.

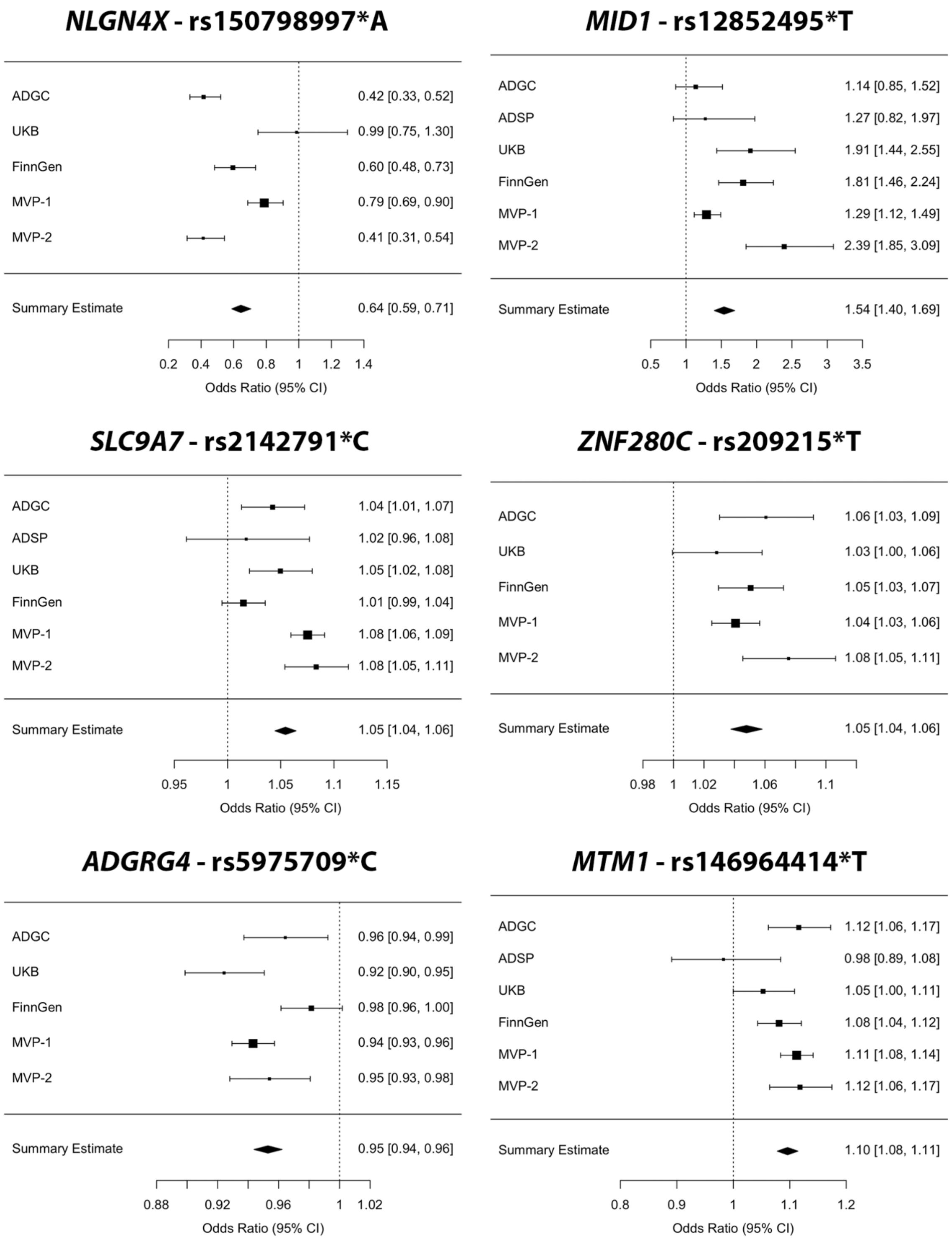

**eFigure 4. Forest plots for all 6 lead variants from the non-stratified AD XWAS.** Panels are marked by locus name and reported variant rsID. The odds ratios are reported with regard to a single active allele. In women, due to random XCI, the relative risk conferred would be half that reported here.

### *NLGN4X* - rs150798997*A

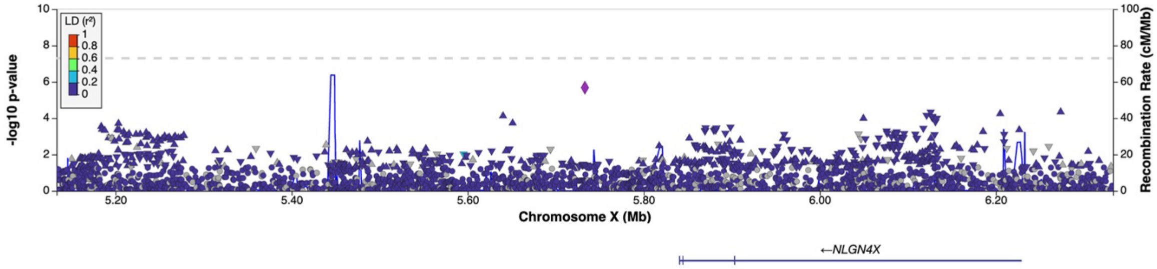

### *MID1* - rs12852495*T

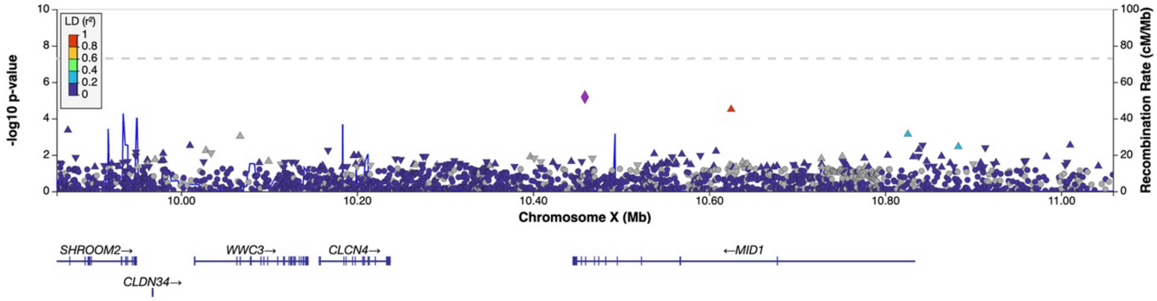

### *SLC9A7* - rs2142791*C

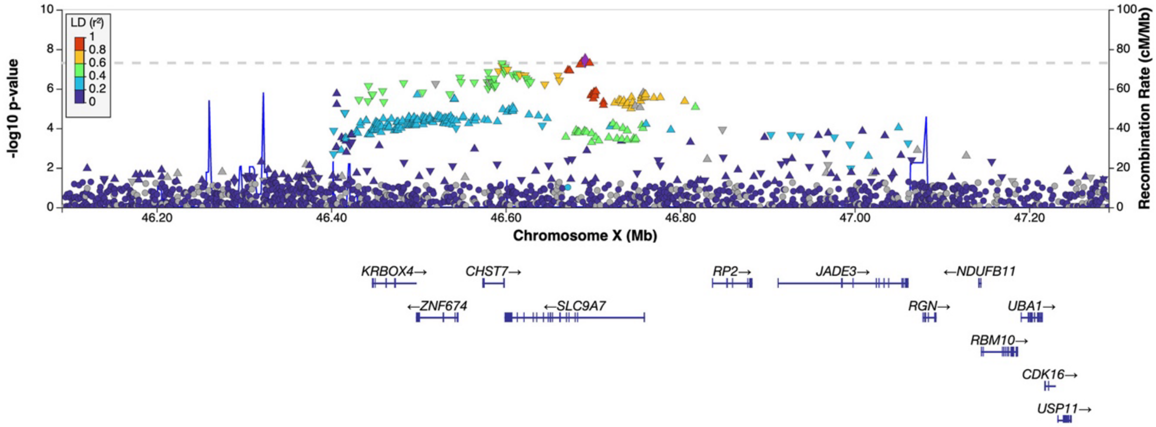

### *ZNF280C* - rs209215*T

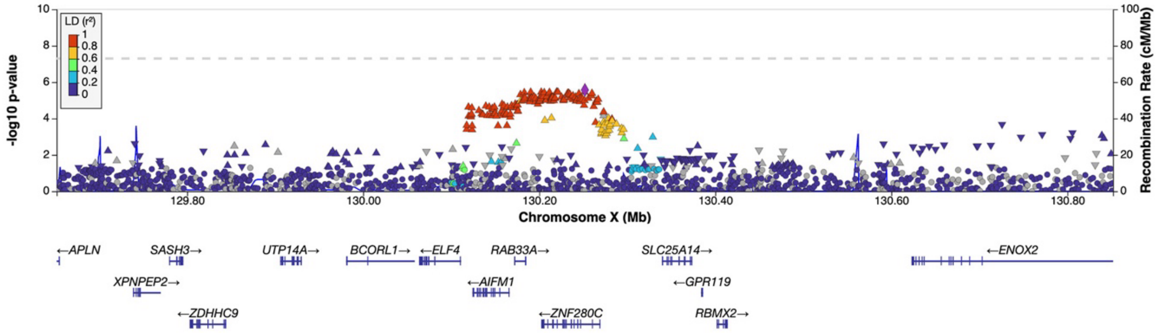

**eFigure 5 (part 1). Locus zoom plots for all 6 lead variants from the non-stratified AD XWAS**. Purple markers indicate the lead variant per locus. Linkage Disequilibrium (LD) is shown for European ancestry.

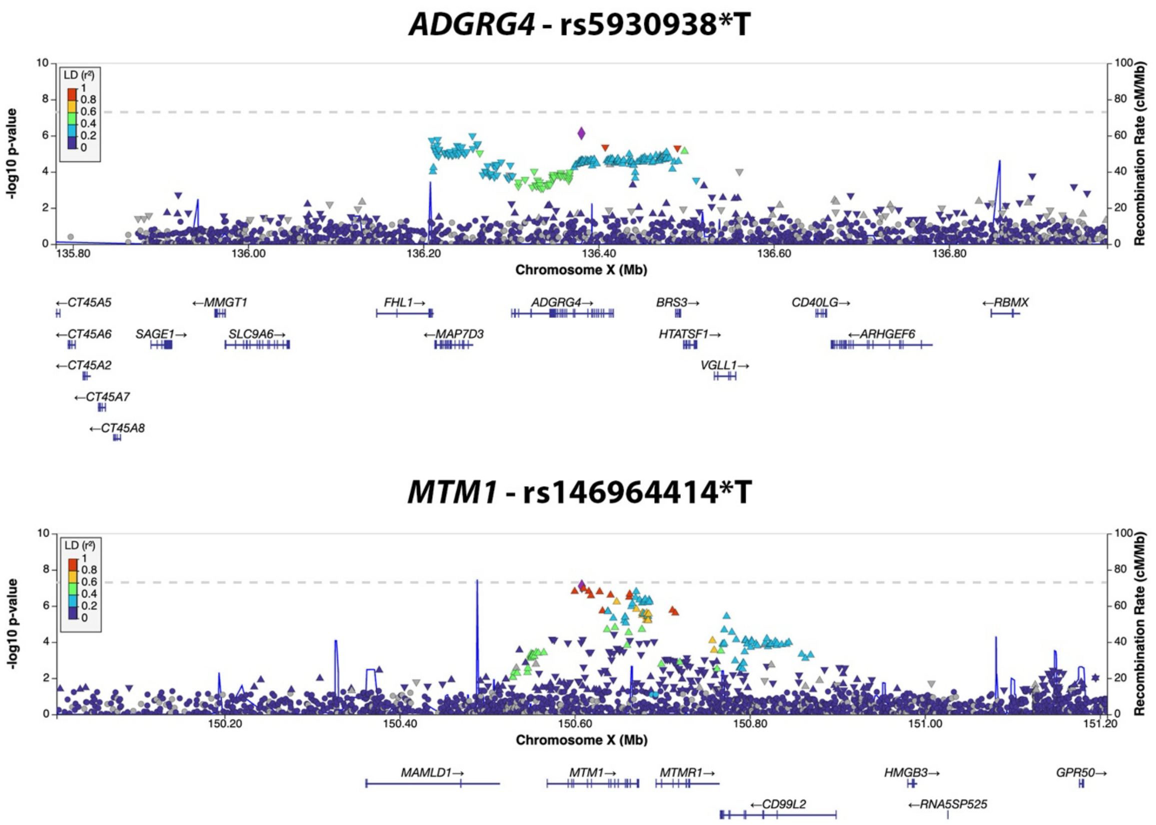

**eFigure 5 (part 2). Locus zoom plots for all 6 lead variants from the non-stratified AD XWAS**. Purple markers indicate the lead variant per locus. Linkage Disequilibrium (LD) is shown for European ancestry

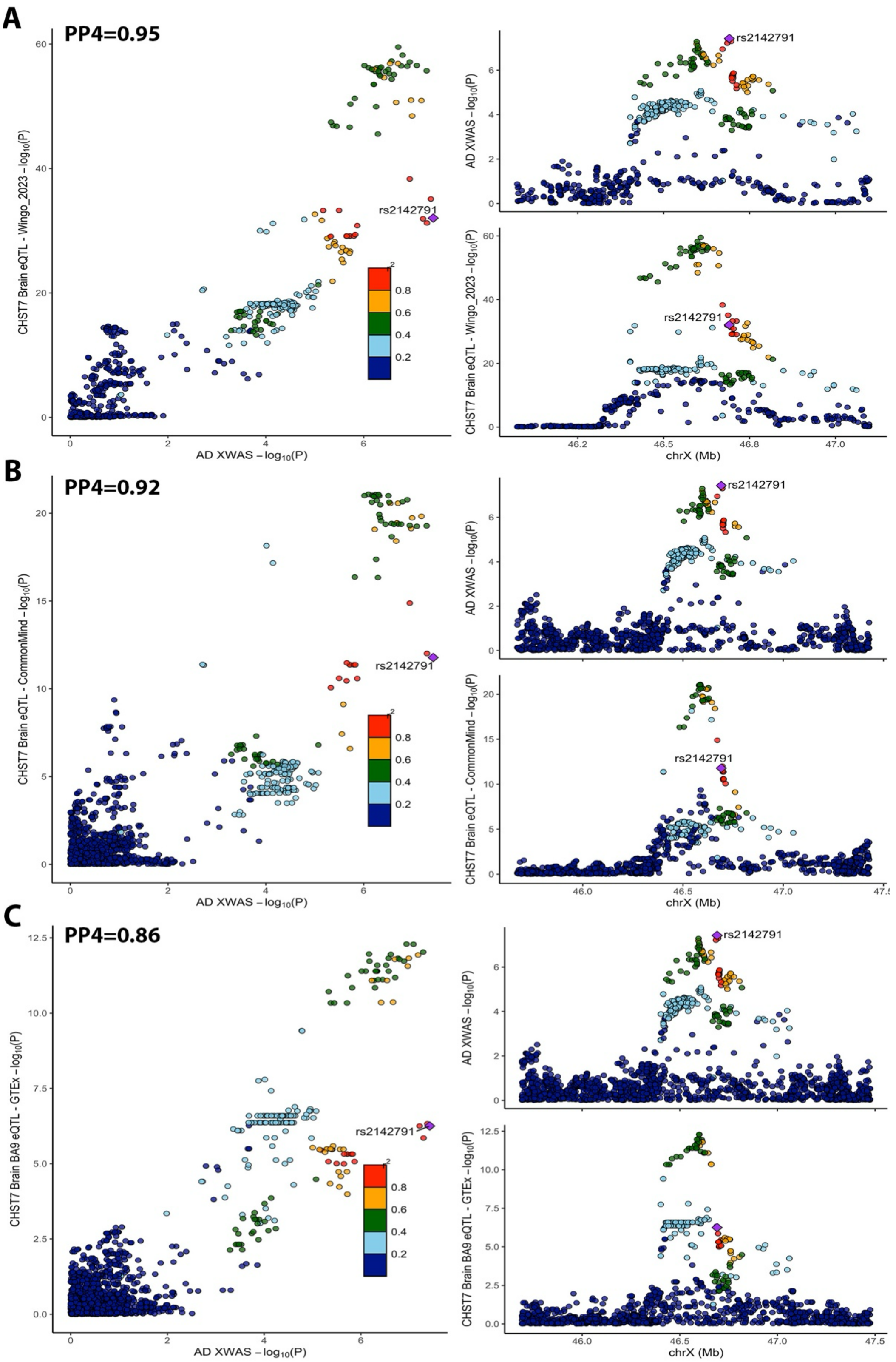

**eFigure 6. *CHST7* Colocalization plots.** For GTEx, only the colocalization with best PP4 in brain (that also had a significant QTL) is visualized. For other brain datasets, colocalizations with PP4>0.7 are visualized.

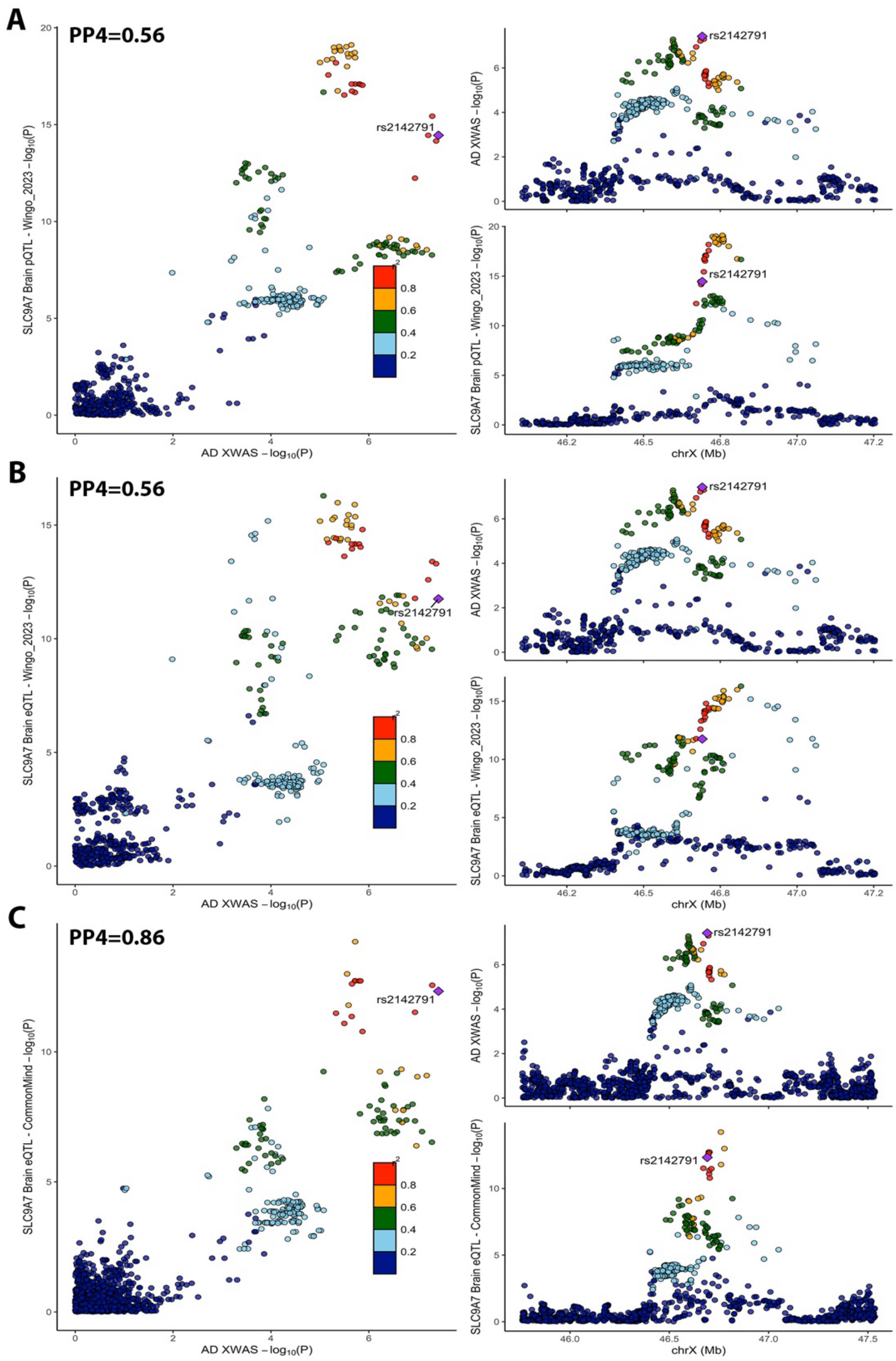

**eFigure 7 (part1).** ***SLC9A7*** **colocalization plots.** For GTEx, only the colocalization with best PP4 in brain and non-brain tissues are respectively visualized. All other brain colocalizations with a relaxed PP4>0.4 threshold are visualized, as well as the colocalization with best PP4 in monocytes.

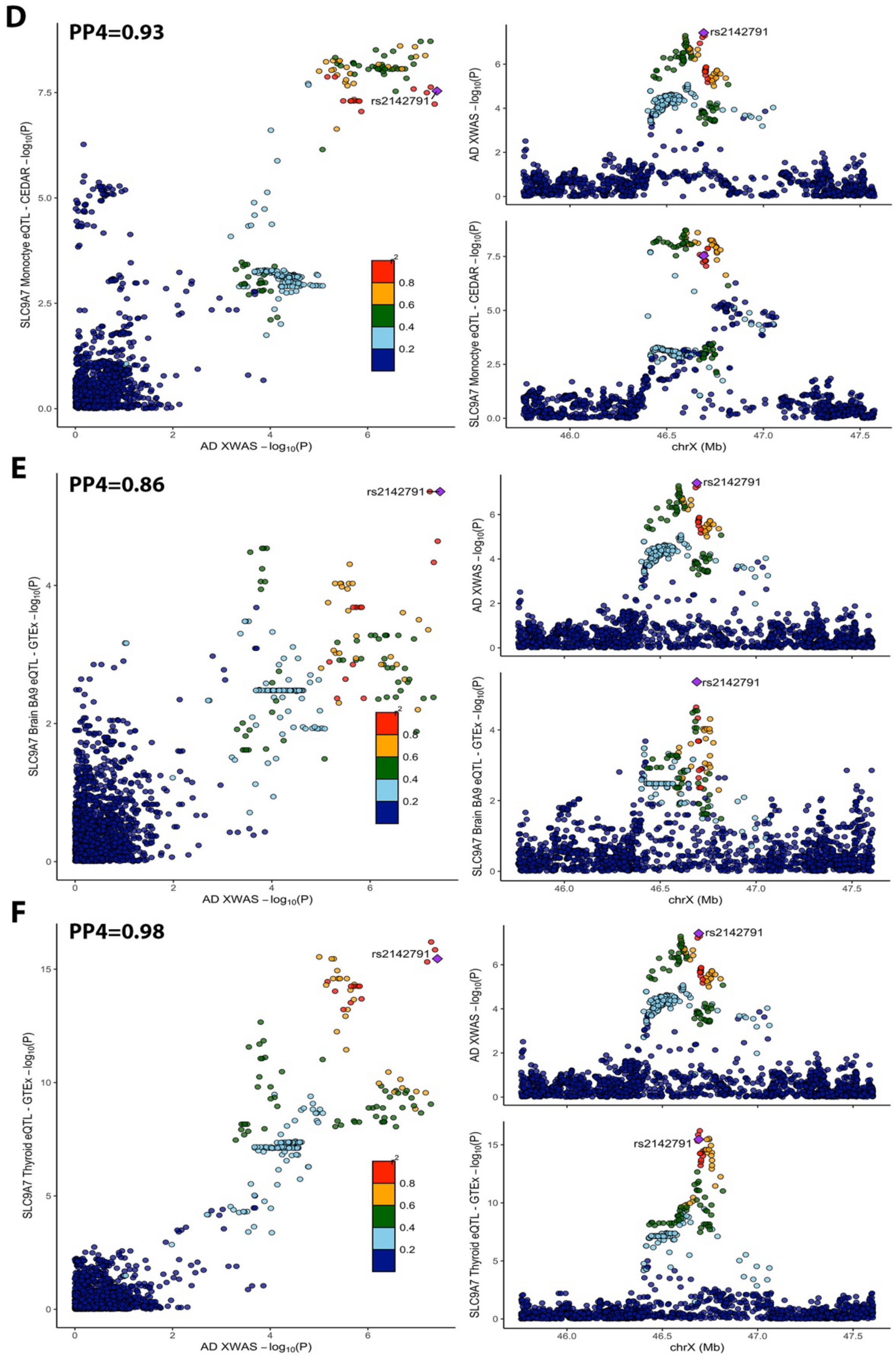

**eFigure 7 (part2). *SLC9A7* colocalization plots.** For GTEx, only the colocalization with best PP4 in brain and non-brain tissues are respectively visualized. All other brain colocalizations with a relaxed PP4>0.4 threshold are visualized, as well as the colocalization with best PP4 in monocytes.

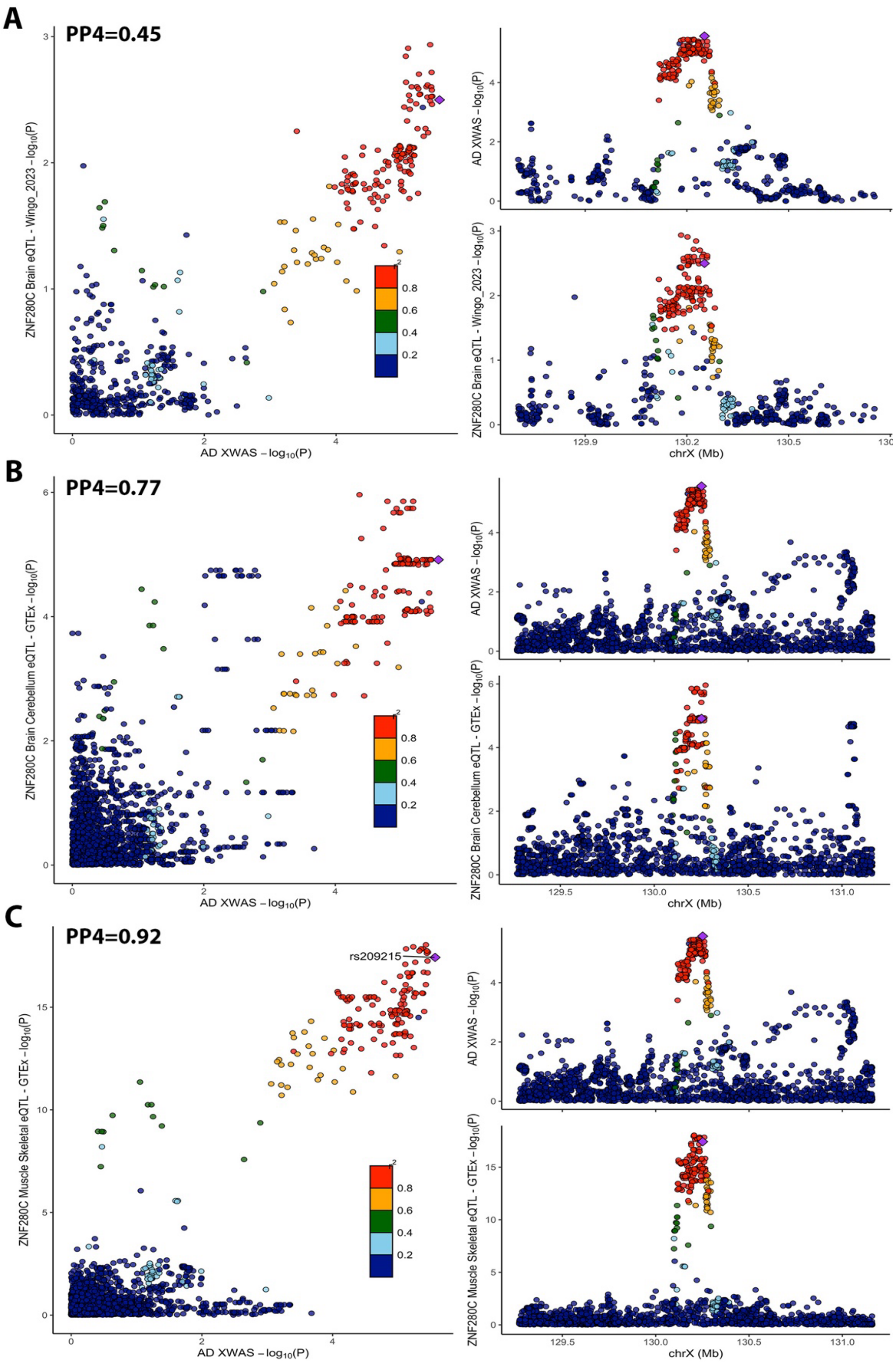

**eFigure 8. *ZNF280C* colocalization plots.** For GTEx, only the colocalizations with best PP4 in brain and non-brain tissues are respectively visualized. Additionally the colocalization in Brain eQTL data from Wingo et al. 2023, passing relaxed PP4>0.4 threshold, is visualized.

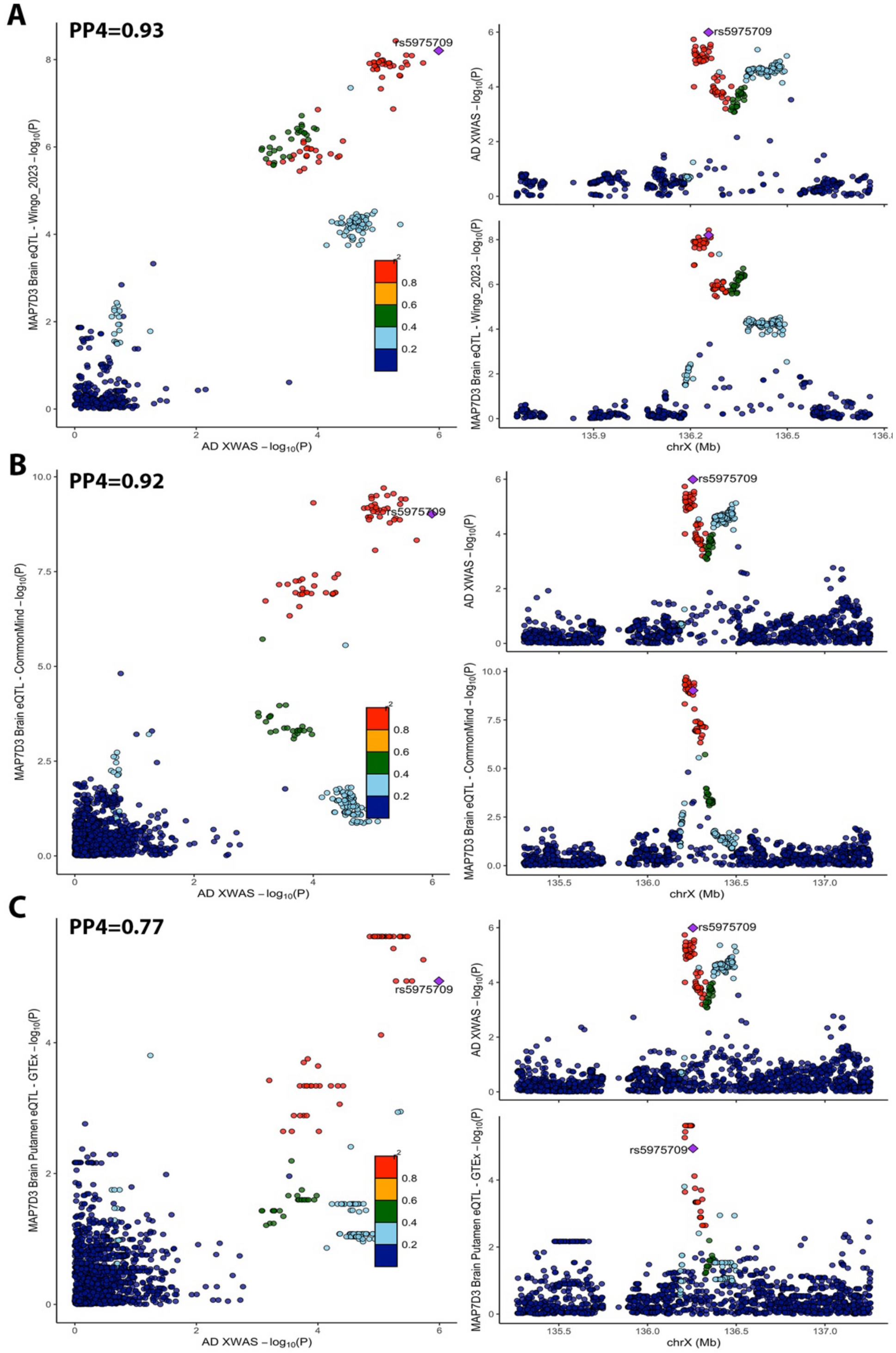

**eFigure 9. *MAP7D3* colocalization plots (part 1).** For GTEx, only the colocalizations with best PP4 in brain and non-brain tissues are respectively visualized. For other brain datasets, colocalizations with PP4>0.7 are visualized.

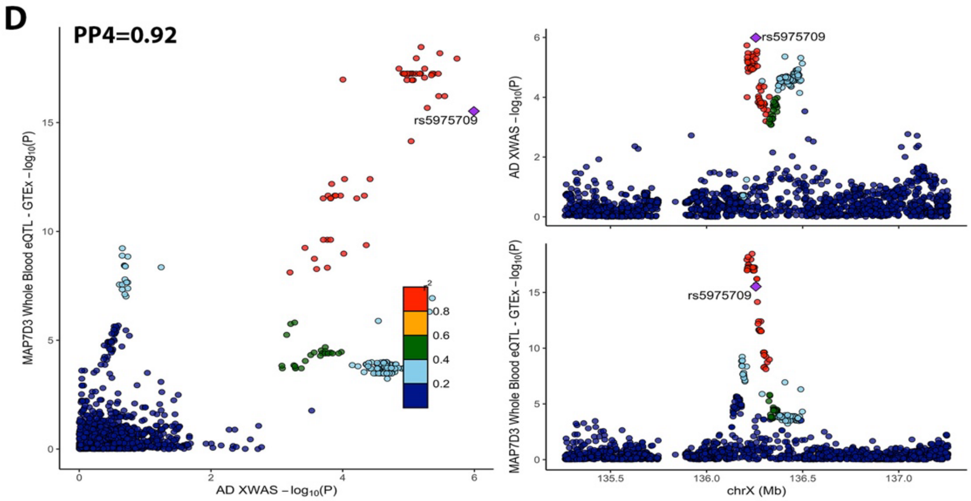

**eFigure 9. *MAP7D3* colocalization plots (part 2).** For GTEx, only the colocalizations with best PP4 in brain and non-brain tissues are respectively visualized. For other brain datasets, colocalizations with PP4>0.7 are visualized.

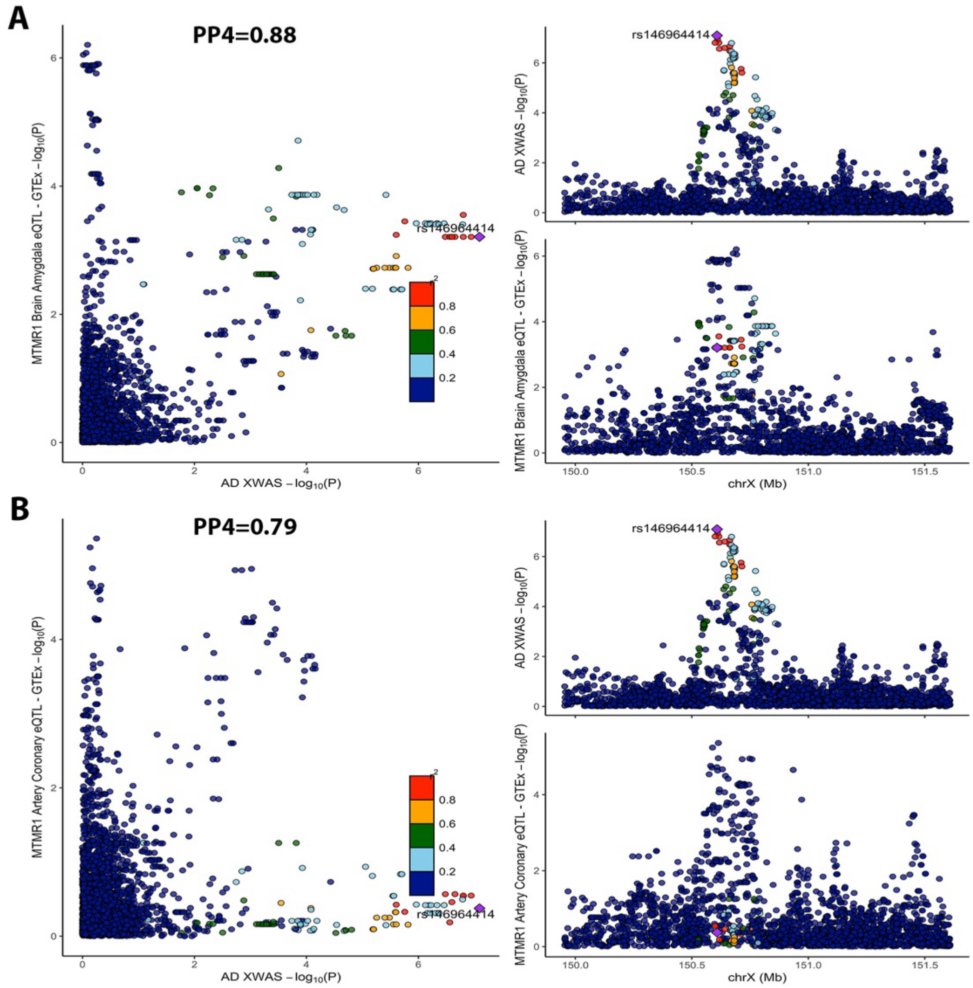

**eFigure 10.** ***MTMR1*** **colocalization plots.** The two colocalizations with PP4>0.7 are visualized. Although PP4 values were >0.7, visually, the plots suggest there is uncertainty in the colocalization with a second independent QTL signal appearing to be present.

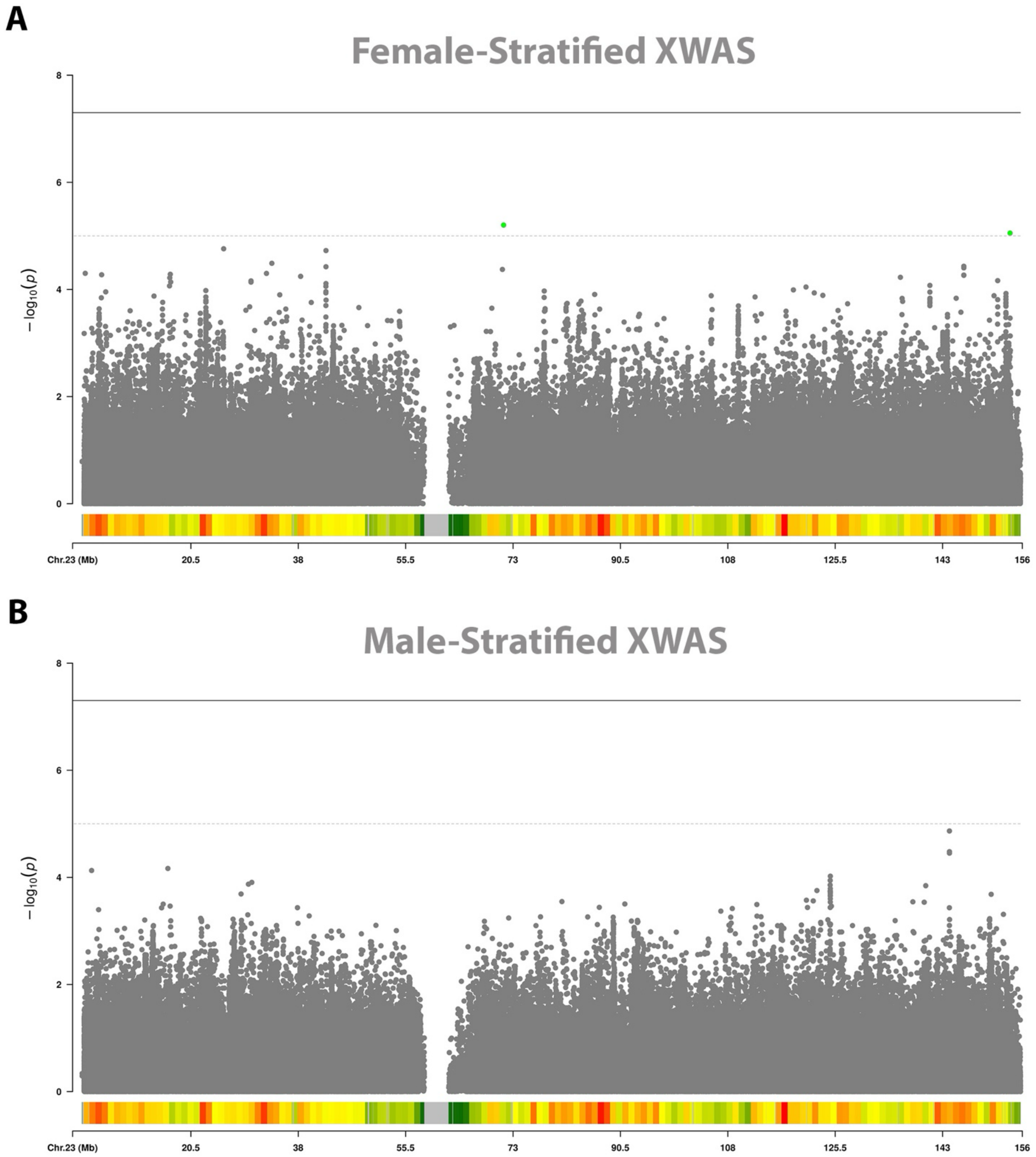

**eFigure 11. Sex-stratified AD XWAS across ADGC, ADSP, and UKB. A)** Female-stratified AD XWAS Manhattan plot. **B)** Male-stratified AD XWAS Manhattan plot. The dotted line indicates X-chromosome-wide significance (P-value<1e-5) and full line genome-wide significance (P-value<5e-8). Green dots indicate variants passing the X-chromosome-wide threshold. The histogram at the bottom indicates variant density (increasing density from green, to yellow, to orange, and red).

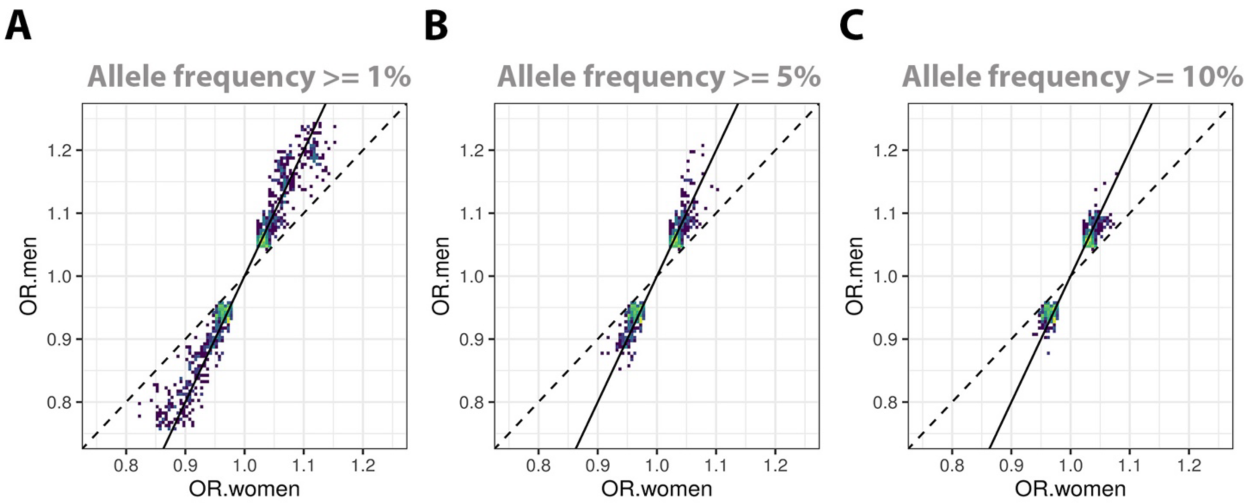

**eFigure 12. Evaluation of escape from XCI in sex-stratified AD XWAS across ADGC, ADSP, and UKB.** Scatter density plots compare beta coefficients from men and women for prioritized variants. Intensity increases from dark blue to bright yellow. Lines with slope=1 (dashed) and slope=2 (solid) are plotted for reference and respectively indicate expectations for escape and no escape from XCI (similar as in Sidorenko et al. 2019[55]; for escape from XCI, the expectation is that beta coefficients from men and women are consistent). The prioritized variants had P<0.1 in both men and women, concordant effect directions across men and women, and allele frequencies **(A)** >= 1%, **(B)** >= 5%, **(C)** >= 10% (these variants are more likely to include true associations and notably include local association signals on the *MID1*, *ZNF280C*, and *ADGRG4* loci). Overall, some common variants fall on or close to slope=1, suggesting they escape XCI with regard to AD. Theoretically, it would be the most specific to evaluate sex-specificity in data with clinically confirmed cases only (ADGC+ADSP), but given the small effect sizes or low frequencies of the lead variants, it was reasoned that the best evaluation would be based on the largest available sample size (ADGC+ADSP+UKB).

*Abbreviations: OR.women; Odds Ratio in women; OR.men; Odds Ratio in men.*

### eTables.

**eTable 1. Overview of genotyping platforms across all available AD-related genetic data.**

| Cohort/Study | Genotyping Platform | Cohort-Platform ID | Sample count | Data Repository |
|---|---|---|---|---|
| ACT | Illumina Human 660W-Quad | ACT | 2790 | NIAGADS (NG00034) / dbGaP (phs000234) |
| ADC1 | Illumina Human 660W-Quad | ADC1 | 2731 | NIAGADS (NG00022) / NACC |
| ADC2 | Illumina Human 660W-Quad | ADC2 | 928 | NIAGADS (NG00023) / NACC |
| ADC3 | Illumina Human OmniExpress | ADC3 | 1526 | NIAGADS (NG00024) / NACC |
| ADC4 | Illumina Human OmniExpress | ADC4 | 1054 | NIAGADS (NG00068) / NACC |
| ADC5 | Illumina Human OmniExpress | ADC5 | 1224 | NIAGADS (NG00069) / NACC |
| ADC6 | Illumina Human OmniExpress | ADC6 | 1333 | NIAGADS (NG00070) / NACC |
| ADC7 | Illumina Infinium Human OmniExpressExome | ADC7 | 1462 | NIAGADS (NG00071) / NACC |
| ADDNEUROMED | Illumina Human 610-Quad | ADM_Q | 315 | Synapse AddNeuroMed (syn4907804) |
| | Illumina Human OmniExpress | ADM_O | 329 | Synapse AddNeuroMed (syn4907804) |
| ADNI | Illumina Human 610-Quad | ADNI_1 | 757 | LONI ADNI |
| | Illumina Human OmniExpress | ADNI_2 | 361 | LONI ADNI |
| | Illumina Global Screening Array (GSA) | ADNI_3 | 327 | LONI ADNI |
| | Illumina Omni 2.5 | ADNI_O25 | 812 | LONI ADNI |
| | Whole Genome Sequencing - Illumina | ADNI_WGS | 812 | LONI ADNI |
| ADNI-DOD | Illumina Human OmniExpress | ADNI_DOD | 204 | LONI ADNIDOD |
| ADSP WGS | Whole Genome Sequencing | ADSP_WGS | 16906 | NIAGADS DSS (NG00067.v5) / NACC |
| GenADA | Affymetrix 500K | GSK | 1571 | dbGaP (phs000219) |
| NIA-LOAD | Illumina Human 610-Quad | LOAD | 5220 | NIAGADS (NG00020) |
| MAYO | Illumina Human Hap300 | MAYO_1 | 2099 | Synapse AMP-AD (syn5591675) / NIAGADS (NG00029) |
| MAYO2 | Illumina Omni 2.5 | MAYO_2 | 314 | Synapse AMP-AD (syn5550404) |
| MIRAGE | Illumina Human CNV370-Duo | MIRAGE_370 | 397 | NIAGADS (NG00031) |
| | Illumina Human 610-Quad | MIRAGE_610 | 1105 | NIAGADS (NG00031) |
| OHSU | Illumina Human CNV370-Duo | OHSU | 647 | NIAGADS (NG00017) |
| ROSMAP | Affymetrix GeneChip 6.0 - Broad Institute | ROSMAP_1B | 1126 | RADC Rush (contact:) / Synapse AMP-AD |
| | Affymetrix GeneChip 6.0 - TGen | ROSMAP_1T | 582 | RADC Rush (contact:) / Synapse AMP-AD |
| | Illumina Human OmniExpress 12 - Chop | ROSMAP_2C | 382 | RADC Rush (contact:) / Synapse AMP-AD |

| | | | | |
|---|---|---|---|---|
| | Illumina Multi-Ethnic - BU | ROSMAP_3BU | 494 | RADC Rush (contact:) |
| TARCC | Affymetrix 6.0 | TARCC | 625 | NIAGADS (NG00097) |
| | Illumina Multi-Ethnic – BU | TARCC_full | 2718 | TARCC (contact:) |
| TGEN2 | Affymetrix 6.0 | TGEN | 1599 | NIAGADS (NG00028) |
| UPITT | Illumina Human Omni1-Quad | UPITT | 2440 | NIAGADS (NG00026) |
| | Illumina Human 1M-Duo, Illumina 1M | UVM_A | 1153 | NIAGADS (NG00042) |
| UM/VU/MSSM | Affymetrix 6.0 | UVM_B | 864 | NIAGADS (NG00042) |
| | Illumina Human 550K. Illumina Human 610-Quad | UVM_C | 445 | NIAGADS (NG00042) |
| WASHU | Illumina Human 610-Quad | WASHU_1 | 670 | NIAGADS (NG00030) |
| WASHU2 | Illumina Human OmniExpress | WASHU_2 | 235 | NIAGADS (NG00087) |
| WHICAP | Illumina Human OmniExpress | WHICAP | 647 | NIAGADS (NG00093) |

**eTable 2. Overview of ADSP available through NIAGADS DSS (NG00067).**

| Study | Accession Number | Related Datasets |
|---|---|---|
| Accelerating Medicines Partnership- Alzheimer's Disease (AMP-AD) | sa000011 | NG00067 – ADSP Umbrella |
| Cache County Study | sa000014 | NG00067 – ADSP Umbrella |
| University of Pittsburgh- Kamboh WGS | sa000012 | NG00067 – ADSP Umbrella |
| CurePSP and Tau Consortium PSP WGS | sa000016 | NG00067 – ADSP Umbrella |
| NIH, CurePSP and Tau Consortium PSP WGS | sa000015 | NG00067 – ADSP Umbrella |
| UCLA Progressive Supranuclear Palsy | sa000017 | NG00067 – ADSP Umbrella |
| NACC Genentech WGS | sa000013 | NG00067 – ADSP Umbrella |
| Alzheimer's Disease Sequencing Project (ADSP) | sa000001 | NG00067 – ADSP Umbrella |
| Alzheimer's Disease Neuroimaging Initiative (ADNI) | sa000002 | NG00067 – ADSP Umbrella |
| Alzheimer's Disease Genetics Consortium: African Americans (ADGC AA) | sa000003 | NG00067 – ADSP Umbrella |
| The Familial Alzheimer Sequencing (FASe) project | sa000004 | NG00067 – ADSP Umbrella |
| Brkanac – Family-based genome scan for AAO of LOAD | sa000005 | NG00067 – ADSP Umbrella |
| HIHG Miami Families with AD | sa000006 | NG00067 – ADSP Umbrella |
| Washington Heights/Inwood Columbia Aging Project (WHICAP) | sa000007 | NG00067 – ADSP Umbrella |
| Charles F. and Joanne Knight Alzheimer's Disease Research Center (Knight ADRC) | sa000008 | NG00067 – ADSP Umbrella |
| Corticobasal degeneration Study (CBD) | sa000009 | NG00067 – ADSP Umbrella |
| Progressive Supranuclear Palsy Study (PSP) | sa000010 | NG00067 – ADSP Umbrella |

**eTable 3. Overview of participant demographics.**

| Dataset | | Diagnosis | | Pathology | | Sex | Age |
|---|---|---|---|---|---|---|---|
| Name | Participants after QC (N) | Type | (N (%)) | Available (N (%)) | AD Path. (N (%)) | Female (N (%)) | Age (Mean (SD)) |
| **ADGC §** | 23,120 | CN | 11,582 (50.1 %) | 1,359 (11.7 %) | 237 (17.4 %) | 6,930 (59.8 %) | 77.8 (8.9) |
| | | clinical-AD | 11,538 (49.9 %) | 4,018 (34.8 %) | 4,018 (100 %) | 6,940 (60.1 %) | 74.3 (7.7) |
| **ADSP §** | 6,487 | CN | 2,944 (45.4 %) | 595 (20.2 %) | 43 (7.2 %) | 1,789 (60.8 %) | 81.6 (6.6) |
| | | clinical-AD | 3,543 (54.6 %) | 2,009 (56.7 %) | 2,009 (100 %) | 2,020 (57.0 %) | 76.7 (8.3) |
| **UKB ¶** | 433,124 | CN | 379,520 (87.6 %) | 0 (0.0 %) | 0 (0.0 %) | 207,233 (54.6 %) | 75.3 (9.9) |
| | | registry-AD | 2,050 (0.5 %) | 0 (0.0 %) | 0 (0.0 %) | 1,054 (51.4 %) | 70.2 (5.3) |
| | | proxy-ADD | 51,554 (11.9 %) | 0 (0.0 %) | 0 (0.0 %) | 39,155 (75.9 %) | 84.2 (6.3) |
| **FinnGen ‡** | 412,181 | CN | 396,564 (96.2 %) | 0 (0.0 %) | 0 (0.0 %) | 223,435 (53.3 %) | 63.0 (-) ‡ |
| | | registry-AD | 15,617 (3.8 %) | 0 (0.0 %) | 0 (0.0 %) | 6,875 (44.0 %) | 78.3 (-) ‡ |
| **MVP-1** | 117,120 | CN | 93,696 (80.0 %) | 0 (0.0 %) | 0 (0.0 %) | 3,514 (3.8 %) | 77.3 (7.3) |
| | | registry-ADD | 23,424 (20.0 %) | 0 (0.0 %) | 0 (0.0 %) | 665 (3.8 %) | 81.6 (7.9) |
| **MVP-2 †** | 160,252 | CN | 129,420 (80.8 %) | 0 (0.0 %) | 0 (0.0 %) | 129,420 (100 %) | 70.6 (11.6) |
| | | proxy-ADD | 30,832 (19.2 %) | 0 (0.0 %) | 0 (0.0 %) | 30,832 (100 %) | 73.6 (8.5) |
| **Meta-analysis** | 1,152,284 | CN | 1,013,726 (88.0 %) | 1,954 (0.2 %) | 280 (14.3) | 572,321 (56.5 %) | 70.1 (-) |
| | | Any-AD/ADD | 138,558 (12.0 %) | 6,027 (4.3 %) | 6,027 (100 %) | 87,541 (63.2 %) | 79.4 (-) |

*Abbreviations: CN, cognitively normal; AD, Alzheimer's disease; ADD, Alzheimer's disease-and-dementia; N, number; QC, quality control; SD, standard deviation.*

§ Across ADGC and ADSP, 40% of clinically diagnosed cases were additionally verified to have Alzheimer's disease pathology.

¶ In UKB, reported sex for cases was based on the sex-specificity of proxy and health registry case status, and for controls based on the sex status of the most informative (i.e. older age) parent versus offspring. When there was no sex-specificity, the sex counts were divided.

‡ Age information was not directly available in FinnGen. For controls, it was inferred from a recent research article on FinnGen[48], and for cases, it was determined using the FinnGen endpoint browser (https://r10.risteys.finngen.fi/)

† In MVP-2, analyses were focussed on maternal phenotypes (representing the majority of samples since paternal phenotypes cannot be used for men in proxy XWAS and there further was a relative paucity of women). Age for parents was not available, but all subjects were at least 45 years old at last visit, such that on average parents would be expected to be at least 60 years of age. The reported age in the table is the subject/offspring age and thus does not directly reflect parental age.

**eTable 4. Overview of variant counts in ADGC cohorts with SNP arrays**.

| Cohort-Platform ID | No. variants pre-QC | No. variants post-QC | No. variants after imputation (Rsq>=0.3) |
|---|---|---|---|
| ACT | 13355 | 8044 | 1517800 |
| ADC1 | 11227 | 6486 | 1494962 |
| ADC2 | 12051 | 7147 | 943391 |
| ADC3 | 14784 | 9015 | 1207463 |
| ADC4 | 14172 | 8535 | 1047484 |
| ADC5 | 14260 | 8589 | 1102801 |
| ADC6 | 14001 | 8406 | 1154153 |
| ADC7 | 14188 | 8558 | 1186710 |
| ADM_Q | 10129 | 5975 | 476723 |
| ADM_O | 0 | - | - |
| ADNI_1 | 17681 | 8693 | 850366 |
| ADNI_2 | 17707 | 10559 | 624153 |
| ADNI_3 | 31770 | 15601 | 626454 |
| ADNI_O25 | 55208 | 29503 | 932111 |
| ADNI_DOD | 17502 | 10484 | 416377 |
| GSK | 27380 | 3801 | 809771 |
| LOAD | 14927 | 8665 | 1688085 |
| MAYO_1 | 8906 | 5071 | 1277082 |
| MAYO_2 | 54563 | 22622 | 617560 |
| MIRAGE_370 | 8457 | 4883 | 583471 |
| MIRAGE_610 | 14565 | 8433 | 926878 |
| MTC | 14841 | 9061 | 822239 |
| OHSU | 11208 | 5857 | 770890 |
| ROSMAP_1B | 26992 | 15509 | 993056 |
| ROSMAP_1T | 0 | - | - |
| ROSMAP_2C | 14976 | 9052 | 653947 |
| ROSMAP_3BU | 17790 | 8796 | 653626 |
| TARCC | 23913 | 13150 | 819368 |
| TARCC_full | 50968 | 23661 | 1376958 |
| TGEN | 27380 | 14346 | 1067005 |
| UPITT | 15569 | 9020 | 1402980 |
| UVM_A | 10950 | 6535 | 1073412 |
| UVM_B | 23946 | 14476 | 985311 |
| UVM_C | 17230 | 10487 | 733848 |
| WASHU_1 | 5259 | 3189 | 679337 |
| WASHU_2 | 9559 | 6073 | 482874 |
| WHICAP | 14132 | 8471 | 759760 |

**eTable 5. Overview of variant counts across datasets after quality control and intersection with ADGC.**

| Dataset | No. variants prior to meta-analysis | No. variant intersecting with ADGC in meta-analysis |
|---|---|---|
| ADGC § | 437,105 | 437,105 |
| ADSP ¶ | 1,178,129 | 315,098 |
| UKB ‡ | 745,199 | 407,347 |
| FinnGen ‡ | 611,423 | 360,539 |
| MVP-1 † | 583,938 | 427,641 |
| MVP-2 † | 583,938 | 427,641 |

§ ADGC imputed cohorts were merged and variants filtered to genotyping rate >50% and minor allele count > 20, equivalent to minor allele frequencies >= 0.043%.

¶ ADSP variants (N=8,873,418) were filtered to genotyping rate >20% and minor allele count > 2 (equivalent to minor allele frequencies >= 0.015%), followed by standard, sex-specific, and ADSP-specific quality control.

‡ UKB and Finngen variants underwent cohort-specific QC and were then filtered to imputation scores > 0.3 and effect allele frequencies >= 0.05%

† MVP variants underwent cohort-specific QC and were then filtered to imputation scores > 0.4 and allele frequencies >= 0.1% in the full dataset including all ancestries. The subset of European ancestry indivdiuals was then extracted for XWAS and variants were subsequently filtered to allele frequencies >= 0.05%.

**eTable 6. Phenotype scoring and rescaling approach for the UKB non-stratified AD XWAS.** In the AD XWAS in UKB, any subject with a direct AD case status or a first-degree relative with ADD case status was attributed a diagnostic/phenotypic value of 1, while other individuals above the ages of 60 in either offspring or parents were attributed the value of 0. A phenotypic weight/score was then determined for cases based on the respective combination of proxy status, subject sex, and X chromosome inheritance pattern, while modeling random escape from X chromosome inactivation. This score represents the anticipated reduction in estimated beta coefficients, such that the correction factor (1/score) allows rescaling onto a regular case-control effect scale. The final beta coefficient adjustment was determined by averaging correction factors across all cases.

| | **Phenotype in XWAS** | **Phenotype Score** | **correction factor (1/score)** | **Rationale** |
|---|---|---|---|---|
| **Sex-non-specific** | | | | |
| Woman self-AD | 1 | 1 | 1 | Direct association so no correction factor. Parental info is not considered in case of self-AD |
| Woman with mother AD | 1 | 0.25 | 4 | Women have 1 X chromosome from two of the mother X chromosomes, sharing 50% of their genetic information with the mother. There is X chromosome inactivation in the mother, thus phenotye score = (1/2)/2. |
| Woman with father AD | 1 | 0.5 | 2 | Women have 1 X chromsome from father, sharing 50% of their genetic information with the father. There is no X inactivation in the father, thus phenotype score = 1/2. |
| Woman with mother AD & father AD | 1 | 0.75 | 1.33 | Combination of above scores. |
| Man self-AD | 1 | 1 | 1 | Direct association so no correction factor. Parental info is not considered in case of self-AD |
| Man with mother AD | 1 | 0.5 | 2 | Men have only 1 X chromosome from mother, sharing 100% of their genetic information with the mother. There is X chromosome inactivation in mother, thus phenotype score = 1/2. |
| Man with father AD | - | - | - | Paternal phenotype in men is not considered, since men don't inherit X chromsome from their father. |
| Man with sibling AD | 1 | 0.375 | 2.67 | If sibling sex is not known, which is the case in UK Biobank, we take the average of brother/sister scores. |
| Man with brother AD | 1 | 0.5 | 2 | Brothers inherit their X chromosome from their mother, such that the brothers share 50% genetic information. The X chromosome in the brother is active, thus phenotye score = 1/2. |
| Man with sister AD | 1 | 0.25 | 4 | The man shares 50% of his X chromosome genetic information with the sister. The sister also has an X chromosome from the father that the man doesn't. There is X chromsome inactivation in the sister, thus phenotye score = ((1/2)+0)/2. |
| Woman with sibling AD | 1 | 0.4375 | 2.29 | If sibling sex is not known, which is the case in UK Biobank, we take the average of brother/sister scores. |
| Woman with brother AD | 1 | 0.5 | 2 | The woman shares 50% of her X chromosome genetic information with the brother. The X chromosome in the brother is active, thus the phenotype score = 1/2. |
| Woman with sister AD | 1 | 0.375 | 2.67 | Both the woman and sister inherit the same X chromosome from their father (100%) and share 50% of their mother's X chromosome genetic information. There is X chromosome inactivation in the sister, thus phenotype score = (1+1/2)/2. |
| Woman with mother AD & sibling AD | 1 | 0.6875 | 1.45 | Combination of above scores. |
| Woman with father AD & sibling AD | 1 | 0.9375 | 1.07 | Combination of above scores. |
| Woman with father AD & mother AD & sibling AD | 1 | 1 | 1 | Combination of above scores, capped at 1 |
| Man with mother AD & sibling AD | 1 | 0.875 | 1.14 | Combination of above scores. |

**eTable 7. Phenotype scoring and rescaling approach for the UKB sex-stratified AD XWAS.** Compared to eTable6, additional subjects were excluded when sex-specificity was not guaranteed (e.g. both parents had AD or one parent had AD but a sibling had AD while their sex was unknown). Notably, the beta coefficient derived in men following the current approach corresponds to an XCI model, such that the beta coefficients for a single dosage represent a 50% probability of being active. As such, the men beta coefficients are subsequently multiplied by 2 to obtain male-specific beta coefficients without XCI (corresponding to a genotype encoding of 0/1).

| | Phenotype in XWAS | Phenotype Score | correction factor (1/score) | Rationale |
|---|---|---|---|---|
| **Woman-specific** | | | | |
| Woman self-AD | 1 | 1 | 1 | Direct association so no correction factor. Parental info is not considered in case of self-AD |
| Man with mother AD | 1 | 0.5 | 2 | Men have only 1 X chromosome from mother, sharing 100% of their genetic information with the mother. There is X chromosome inactivation in mother, thus phenotype score = 1/2. |
| Woman with mother AD | 1 | 0.25 | 4 | Women have 1 X chromosome from two of the mother X chromosomes, sharing 50% of their genetic information with the mother. There is X chromosome inactivation in the mother, thus phenotye score = (1/2)/2. |
| no AD | 0 | 0 | - | Controls have no AD and ages>60. Parents and siblings also don't have AD. Either parents or offspring are >60y of age. |
| **Man-specific** | | | | |
| Man self-AD | 1 | 1 | 1 | Direct association so no correction factor. Parental info is not considered in case of self-AD |
| Man with father AD | - | - | - | Paternal phenotype in men is not considered, since men don't inherit X chromsome from their father. |
| Woman with father AD | 1 | 0.5 | 2 | Women have 1 X chromsome from father, sharing 50% of their genetic information with the father. There is no X inactivation in the father, thus phenotype score = 1/2. |
| no AD | 0 | 0 | - | Controls have no AD and ages>60. Parents and siblings also don't have AD. Either parents or offspring are >60y of age. |

**eTable 8. Genetic colocalization with quantitative trait locus data: Extension of Table 2 without collapsing results from overlapping tissues.** Evidence for colocalization was considered at colocalization posterior probability (PP4)>0.7 (bolded). The table presents PP4 results and is restricted to genes and datasets/tissues where at least one colocalization reached PP4>0.7. Bolded entries with an asterisk (*) indicate the lead variant was also a significant QTL in the respective data/tissue. Missing entries indicate that no QTL data were available.

| Locus | Gene with COLOC support (PP4 > 0.7) | Number of times prioritized (PP4 > 0.7) | Number of times prioritized (PP4 > 0.7) - unique data | pQTL_Brain_Wingo2023 | eQTL_Brain_Wingo2023 | eQTL_Brain_CommonMind | eQTL_monocytes_naive_Fairfax2014 | eQTL_monocytes_IFN24_Fairfax2014 | eQTL_monocytes_LPS2_Fairfax2014 | eQTL_monoctyes_CEDAR | eQTL_GTEx_Brain_Amygdala | eQTL_GTEx_Brain_Anterior_cingulate_cortex_BA24 | eQTL_GTEx_Brain_Caudate_basal_ganglia | eQTL_GTEx_Brain_Cerebellum | eQTL_GTEx_Brain_Frontal_Cortex_BA9 | eQTL_GTEx_Brain_Hypothalamus | eQTL_GTEx_Brain_Putamen_basal_ganglia | eQTL_GTEx_Whole_Blood | eQTL_GTEx_Adipose_Subcutaneous | eQTL_GTEx_Adipose_Visceral_Omentum | eQTL_GTEx_Adrenal_Gland | eQTL_GTEx_Artery_Aorta | eQTL_GTEx_Artery_Coronary | eQTL_GTEx_Artery_Tibial | eQTL_GTEx_Breast_Mammary_Tissue | eQTL_GTEx_Cells_Cultured_fibroblasts | eQTL_GTEx_Esophagus_Gastroesophageal_Junction | eQTL_GTEx_Esophagus_Mucosa | eQTL_GTEx_Esophagus_Muscularis | eQTL_GTEx_Heart_Left_Ventricle | eQTL_GTEx_Lung | eQTL_GTEx_Muscle_Skeletal | eQTL_GTEx_Nerve_Tibial | eQTL_GTEx_Pancreas | eQTL_GTEx_Pituitary | eQTL_GTEx_Skin_Not_Sun_Exposed_Suprapubic | eQTL_GTEx_Skin_Sun_Exposed_Lower_leg | eQTL_GTEx_Small_Intestine_Terminal_Ileum | eQTL_GTEx_Stomach | eQTL_GTEx_Testis | eQTL_GTEx_Thyroid |
|---|---|---|---|---|---|---|---|---|---|---|---|---|---|---|---|---|---|---|---|---|---|---|---|---|---|---|---|---|---|---|---|---|---|---|---|---|---|---|---|---|---|
| ***SLC9A7*** | ***ENSG00000286306*** | **1** | **1** | | | **0.88** | | | | | | | | | | | | | | | | | | | | | | | | | | | | | | | | | | | |
| | ***KRBOX4*** | **4** | **2** | | **0.77*** | 0.04 | 0.10 | 0.06 | 0.08 | 0.05 | 0.05 | 0.22 | 0.20 | **0.74** | 0.06 | 0.08 | 0.10 | 0.23 | 0.00 | 0.03 | 0.05 | 0.03 | 0.05 | 0.03 | 0.03 | 0.03 | 0.36 | 0.07 | **0.92** | 0.03 | 0.03 | 0.03 | 0.04 | 0.24 | 0.13 | 0.46 | 0.23 | 0.07 | 0.06 | 0.08 | **0.76** |
| | ***CHST7*** | **10** | **3** | | **0.95*** | **0.92*** | 0.27 | 0.18 | 0.16 | 0.15 | **0.79** | **0.87** | **0.93** | **0.81*** | **0.86*** | **0.79*** | **0.79*** | 0.20 | 0.00 | 0.07 | 0.16 | **0.71** | 0.32 | 0.13 | 0.03 | 0.40 | 0.04 | 0.04 | 0.10 | 0.07 | 0.05 | 0.07 | 0.11 | 0.22 | 0.04 | 0.03 | 0.06 | 0.05 | 0.06 | 0.07 | 0.59 |
| | ***SLC9A7*** | **12** | **4** | 0.56 | 0.56 | **0.86*** | **0.92*** | **0.93*** | **0.93*** | **0.93*** | 0.08 | 0.15 | 0.05 | 0.05 | 0.64 | 0.06 | 0.05 | 0.39 | 0.02 | **0.96*** | 0.13 | 0.63 | 0.49 | 0.57 | **0.79** | **0.90*** | 0.58 | 0.66 | 0.56 | **0.94*** | 0.02 | 0.69 | 0.60 | 0.64 | 0.05 | 0.10 | 0.67 | **0.77** | **0.85** | 0.08 | **0.98*** |
| | ***RP2*** | **1** | **1** | **0.89*** | 0.04 | 0.07 | 0.05 | 0.50 | 0.06 | 0.05 | 0.13 | 0.05 | 0.07 | 0.04 | 0.10 | 0.07 | 0.05 | 0.03 | 0.03 | 0.03 | 0.09 | 0.04 | 0.05 | 0.02 | 0.03 | 0.00 | 0.03 | 0.03 | 0.08 | 0.04 | 0.03 | 0.04 | 0.03 | 0.06 | 0.04 | 0.02 | 0.14 | 0.06 | 0.04 | 0.05 | 0.02 |
| | ***JADE3*** | **2** | **2** | | **0.70*** | 0.07 | 0.04 | 0.06 | 0.10 | 0.43 | 0.10 | 0.06 | 0.10 | 0.05 | 0.11 | 0.08 | 0.51 | 0.06 | 0.06 | 0.09 | 0.08 | 0.66 | 0.08 | 0.04 | 0.32 | 0.06 | 0.11 | **0.76** | 0.18 | 0.11 | 0.26 | 0.02 | 0.03 | 0.03 | 0.12 | 0.04 | 0.04 | 0.21 | 0.03 | 0.06 | 0.54 |
| | ***UBA1*** | **1** | **1** | 0.05 | 0.04 | **0.93*** | 0.13 | 0.07 | 0.09 | 0.12 | 0.12 | 0.07 | 0.05 | 0.05 | 0.05 | 0.06 | 0.09 | 0.07 | 0.03 | 0.03 | 0.05 | 0.03 | 0.05 | 0.03 | 0.05 | 0.03 | 0.04 | 0.03 | 0.03 | 0.09 | 0.04 | 0.04 | 0.00 | 0.04 | 0.05 | 0.10 | 0.03 | 0.05 | 0.04 | 0.05 | 0.00 |
| | ***ELK1*** | **1** | **1** | | 0.00 | 0.04 | 0.11 | **0.86** | 0.06 | 0.08 | 0.05 | 0.04 | 0.33 | 0.05 | 0.10 | 0.18 | 0.09 | 0.03 | 0.02 | 0.04 | 0.16 | 0.03 | 0.04 | 0.03 | 0.03 | 0.02 | 0.03 | 0.11 | 0.03 | 0.03 | 0.03 | 0.02 | 0.02 | 0.06 | 0.07 | 0.04 | 0.03 | 0.05 | 0.03 | 0.05 | 0.04 |
| ***ZNF280C*** | ***ELF4*** | **1** | **1** | | | 0.05 | 0.01 | 0.01 | 0.16 | 0.06 | 0.05 | 0.06 | 0.03 | 0.11 | 0.03 | 0.03 | 0.04 | 0.02 | 0.10 | 0.06 | 0.02 | 0.02 | 0.07 | 0.00 | **0.78*** | 0.02 | 0.04 | 0.01 | 0.30 | 0.37 | 0.02 | 0.59 | 0.03 | 0.04 | 0.05 | 0.02 | 0.02 | 0.04 | 0.02 | 0.09 | 0.03 |
| | ***AIFM1*** | **6** | **3** | 0.02 | 0.16 | 0.12 | 0.14 | **0.73** | 0.50 | **0.78*** | 0.04 | 0.09 | 0.07 | 0.56 | 0.05 | 0.07 | 0.08 | 0.19 | **0.74*** | 0.21 | 0.02 | 0.11 | 0.03 | 0.36 | 0.02 | 0.17 | 0.26 | 0.23 | 0.12 | 0.02 | 0.13 | 0.00 | 0.66 | **0.78** | 0.20 | **0.78*** | **0.74*** | 0.06 | 0.07 | 0.60 | 0.25 |
| | ***ZNF280C*** | **17** | **1** | | 0.45 | 0.09 | 0.07 | 0.04 | 0.19 | 0.04 | 0.42 | 0.21 | 0.10 | **0.77*** | 0.30 | 0.33 | 0.04 | 0.05 | **0.83*** | **0.86*** | 0.66 | **0.87*** | 0.03 | **0.89*** | 0.67 | **0.84*** | **0.79*** | **0.85*** | **0.89*** | 0.69 | **0.83*** | **0.92*** | **0.86*** | 0.48 | **0.79*** | **0.87*** | **0.84*** | 0.11 | **0.71** | 0.16 | **0.85*** |
| | ***RBMX2*** | **1** | **1** | | 0.00 | 0.29 | 0.04 | 0.04 | 0.04 | 0.05 | 0.04 | 0.04 | 0.04 | 0.05 | 0.07 | 0.04 | 0.04 | 0.02 | 0.00 | 0.00 | 0.02 | 0.00 | 0.02 | 0.00 | 0.04 | 0.00 | 0.03 | 0.02 | 0.06 | **0.77** | 0.02 | 0.01 | 0.00 | 0.02 | 0.04 | 0.01 | 0.02 | 0.03 | 0.02 | 0.06 | 0.04 |
| ***ADGRG4*** | ***FHL1*** | **2** | **2** | 0.12 | **0.78*** | 0.13 | **0.85** | 0.04 | 0.04 | 0.29 | 0.05 | 0.12 | 0.04 | 0.10 | 0.09 | 0.06 | 0.04 | 0.02 | 0.07 | 0.03 | 0.08 | 0.05 | 0.13 | 0.00 | 0.03 | 0.02 | 0.12 | 0.11 | 0.02 | 0.07 | 0.02 | 0.03 | 0.09 | 0.04 | 0.04 | 0.03 | 0.02 | 0.04 | 0.03 | 0.00 | 0.00 |
| | ***MAP7D3*** | **10** | **3** | 0.05 | **0.93*** | **0.92*** | 0.13 | 0.07 | 0.04 | 0.08 | 0.06 | 0.07 | 0.19 | 0.04 | 0.09 | 0.04 | **0.77*** | **0.92*** | 0.07 | 0.02 | **0.72*** | 0.02 | 0.06 | 0.02 | 0.04 | **0.89*** | 0.02 | 0.02 | 0.02 | **0.72** | **0.74** | **0.87*** | 0.10 | 0.23 | 0.04 | 0.39 | 0.05 | 0.46 | 0.45 | **0.89*** | 0.01 |
| | ***BRS3*** | **1** | **1** | | | 0.04 | 0.04 | 0.04 | 0.04 | 0.04 | 0.04 | 0.04 | | | | 0.04 | | | | | | | | | 0.03 | | | | | | | | | 0.05 | 0.03 | | | | | **0.73** | 0.27 |
| | ***HTATSF1*** | **2** | **1** | 0.03 | 0.05 | 0.03 | 0.03 | 0.07 | 0.05 | 0.06 | 0.04 | 0.04 | 0.04 | 0.04 | 0.04 | 0.04 | 0.04 | 0.01 | 0.59 | 0.02 | 0.02 | 0.09 | 0.03 | **0.87*** | 0.02 | 0.02 | 0.02 | 0.02 | 0.02 | 0.02 | 0.04 | 0.02 | 0.08 | 0.03 | 0.03 | 0.02 | **0.86*** | 0.04 | 0.05 | 0.68 | 0.02 |
| | ***AL683813.2*** | **1** | **1** | | | | | | | | 0.05 | 0.04 | 0.05 | 0.03 | 0.05 | 0.04 | 0.07 | 0.02 | 0.02 | 0.03 | 0.09 | 0.13 | 0.04 | 0.02 | 0.02 | 0.02 | 0.02 | 0.02 | 0.02 | 0.02 | 0.14 | **0.77** | 0.02 | 0.02 | 0.04 | 0.02 | 0.02 | 0.05 | 0.02 | 0.21 | 0.02 |
| ***MTM1*** | ***MTMR1*** | **2** | **1** | 0.01 | 0.05 | 0.16 | 0.18 | 0.12 | 0.12 | 0.11 | **0.88** | 0.36 | 0.08 | 0.63 | 0.11 | 0.08 | 0.57 | 0.01 | 0.00 | 0.00 | 0.43 | 0.00 | **0.79** | 0.00 | 0.00 | 0.00 | 0.00 | 0.00 | 0.00 | 0.08 | 0.00 | 0.44 | 0.00 | 0.00 | 0.00 | 0.00 | 0.00 | 0.63 | 0.00 | 0.00 | 0.00 |

**eTable 9. Frequency in cases and controls for *SLC9A7* lead variant across cohorts.**

| Dataset | Allele Frequency Controls | Allele Frequency Cases |
|---|---|---|
| ADGC | 45.75% | 46.99% |
| ADSP | 46.49% | 46.58% |
| UKB † | 45.82% | 46.13% |
| FinnGen | 46.53% | 47.08% |
| MVP-1 | 45.44% | 47.28% |
| MVP-2 † | 45.61% | 46.42% |

† Note that the use of proxy cases causes a dilution of case allele frequencies (which relates to the need to adjust beta coefficients from proxy GWAS and XWAS[63]).

### eTable 10. Evaluation of sex-specific effects and escape from X chromosome inactivation (XCI) for lead variants from non-stratified AD XWAS.

XCI escape was evaluated by dividing beta coefficients from men by beta coefficients from women similar as in Sidorenko et al. 2019[55], where a ratio close to 2 suggests no escape from XCI and a ratio close to 1 suggests escape from XCI. Sex heterogeneity and XCI escape were evaluated for the beta coefficients obtained after meta-analyzing sex-stratified results across ADGC+ADSP+UKB. Theoretically, it would be the most specific to evaluate sex-specificity in data with clinically confirmed cases only (ADGC+ADSP), but given the small effect sizes or low frequencies of the lead variants, it was reasoned that the best evaluation would be based on the largest available sample size (ADGC+ADSP+UKB).

We further identified if there was any prior support for XCI at each locus, focusing on prioritized genes for common variants (cf. Table 2) or nearest genes for rare variants (*NLGN4X* & *MID1*). We consulted 4 resources across 2 published research articles:

- [1a]: Tukiainen et al. 2017 [56] Suppl.Table.1, which reviews XCI status reports from 2 prior papers.
- [1b]: Tukiainen et al. 2017 [56] Suppl.Table.13, which summarizes XCI status reports from their analyses.
- [1a]: Garieri et al. 2018 [57] Dataset S1, which reviews if XCI escape status was reported from 7 prior papers (inactive genes were not listed).
- [1b]: Garieri et al. 2018 [57] Dataset S3, which summarizes XCI status reports from their analyses, and 5 genes reported in their manuscript.

| Locus | non-stratified lead variant | EA | Women - No. subjects | Men - No. subjects | Women - EAF | Men - EAF | Women - Beta | Women - SE | Women - P-value | Men - Beta | Men - SE | Men - P-value | Sex heterogeneity P-value | Men / Women Beta Ratio | Appears to escape XCI in AD? | Prior XCI reports |
|---|---|---|---|---|---|---|---|---|---|---|---|---|---|---|---|---|
| *NLGN4X* | rs150798997 | A | 407,823 | 272,186 | 0.34% | 0.33% | -0.273 | 0.130 | 0.035 | -0.499 | 0.242 | 0.039 | 0.410 | 1.83 | No | Summary: variable escape. [1a] - NLGN4X: variable escape; [1b] - NLGN4X: variable escape; [2a] - NLGN4X: escape; [2b] - NLGN4X: inactive |
| *MID1* | rs12852495 | T | 411,223 | 274,586 | 0.30% | 0.29% | **0.408** | **0.140** | **3.69E-03** | **-0.138** | **0.251** | **0.583** | **0.058** | **-0.34** | **Women specific** | Summary: variable escape. [1a] - MID1: variable escape & inactive; [1b] - MID1: inactive; [2a] - MID1: escape; [2b] - MID1: escape |
| *SLC9A7* | rs2142791 | C | 411,732 | 274,909 | 45.90% | 45.93% | 0.022 | 0.014 | 0.100 | 0.039 | 0.027 | 0.149 | 0.587 | 1.73 | No | Summary: variable escape (appears mostly inactive). [1a] - SLC9A7: inactive; [1a] - CHST7: inactive; [2b] - SLC9A7 - inactive & escape; [2b] - CHST7: escape |
| *ZNF280C* | rs209215 | T | 407,927 | 272,326 | 39.41% | 39.33% | **0.030** | **0.015** | **0.037** | **0.030** | **0.029** | **0.302** | **0.992** | **0.99** | **Yes** | Summary: variable escape. [1a] - ZNF280C: variable escape; [1b] - ZNF280C: inactive; [2b] - ZNF280C: inactive |
| *ADGRG4* | rs5930938 | C | 407,927 | 272,326 | 33.00% | 32.95% | **-0.055** | **0.015** | **2.95E-04** | **-0.066** | **0.030** | **0.031** | **0.759** | **1.19** | **Yes** | Summary: variable escape. [1a] - MAP7D3: inactive; [1b] - MAP7D3: variable escape & inactive; [2a] - MAP7D3: escape; [2b] - MAP7D3: inactive |
| *MTM1* | rs146964414 | T | 411,736 | 274,904 | 8.03% | 8.07% | 0.035 | 0.024 | 0.157 | 0.086 | 0.047 | 0.070 | 0.338 | 2.48 | No | Summary: variable escape. [1a] - MTMR1: inactive; [1b] - MTMR1: inactive; [2a] - MTMR1: escape; [2b] - MTMR1: escape |

**eTable 11. Evaluation of sex-specific effects and escape from XCI for lead variants from female-stratified AD XWAS.**

The approach here is consistent as reported in **eTable10**. Notably, the *TAF1* lead variant showed no significant sex heterogeneity and would suggest escape from XCI, but prior literature suggests *TAF1* does not escape XCI. If there is no sex heterogeneity, this variant should likely have been identified in the larger non-stratified AD XWAS. Overall, this variant would thus appear to be a false positive. The *IRAK1* lead variant in contrast appears female-specific.

| Locus | Women-stratified lead variant | EA | Women - No. subjects | Men - No. subjects | Women - EAF | Men - EAF | Women - Beta | Women - SE | Women - P-value | Men - Beta | Men - SE | Men - P-value | Sex heterogeneity P-value | Men / Women Beta Ratio | Appears to escape XCI in AD? | Prior XCI reports |
|---|---|---|---|---|---|---|---|---|---|---|---|---|---|---|---|---|
| *TAF1* | rs757400922 | C | 409,578 | 273,645 | 0.35% | 0.35% | 0.612 | 0.135 | 6.29E-06 | 0.496 | 0.302 | 0.101 | 0.726 | 0.81 | Yes | Summary: inactive. [1a] - TAF1: inactive; [1b] - TAF1: inactive; [2b] - TAF1: inactive |
| *IRAK1* | rs200796773 | TA | 411,419 | 274,654 | 0.27% | 0.27% | **0.599** | **0.135** | **8.88E-06** | **-0.253** | **0.240** | **0.291** | **1.95E-03** | **-0.42** | **Women-specfic** | Summary: variable escape (appears mostly inactive). [1a] - IRAK1: inactive & variable escape; [1b] - IRAK1: inactive; [2b] - IRAK1: inactive |

**eTable 12. Effect sizes for *SLC9A7* lead variant on *SLC9A7* expression in brain tissue.** The table reports the effect size only for datasets where colocalization with *SLC9A7* expression in brain tissue showed PP4>0.6. CommondMind QTL data was accessed through processed data from the eQTL Catalogue, which used conditional quantile normalization followed by inverse normal transformation[60]. The GTEx effect estimate represents the allelic fold change, i.e. the magnitude of expression change associated with a given genetic variant[64]. Given that *SLC9A7* expression is mainly reported to not escape XCI and similarly did not show signs of XCI escape with regard to AD (cf. **eTable10**), the reported effect sizes in the current table should be considered to reflect the effect of a genotype that has 50% probability of being active (following an XCI model). The effect sizes should thus be doubled to reflect the effect of a fully active genotype.

| Dataset | SLC9A7 - PP4 | SLC9A7 eQTL - effect estimate [95%CI] |
|---|---|---|
| eQTL_Brain_Frontal_Cortex_BA9 | 0.86 | 0.083813 [0.049139, 0.112831] |
| CommonMind_dorsolateral_prefontal_cortex | 0.64 | 0.219761 [0.161608, 0.277915] |

### References

1. Kunkle BW, Grenier-boley B, Sims R, et al. Genetic meta-analysis of diagnosed Alzheimer's disease identifies new risk loci and implicates Aβ, tau, immunity and lipid processing. *Nat Genetic*. 2019;51(3):413-430.

2. Bis JC, Jian X, Chen BWK, et al. Whole exome sequencing study identifies novel rare and common Alzheimer's-Associated variants involved in immune response and transcriptional regulation. *Mol Psychiatry*. 2020;25:1859-1875. doi:10.1038/s41380-018-0112-7

3. Beecham GW, Bis JC, Martin ER, et al. The Alzheimer's disease sequencing project: Study design and sample selection. *Neurol Genet*. 2017;3:e194. doi:10.1212/NXG.0000000000000194

4. Wang M, Beckmann ND, Roussos P, et al. The Mount Sinai cohort of large-scale genomic, transcriptomic and proteomic data in Alzheimer's disease. *Sci Data*. 2018;5:180185. doi:10.1038/sdata.2018.185

5. De Jager PL, Ma Y, McCabe C, et al. A multi-omic atlas of the human frontal cortex for aging and Alzheimer's disease research. *Sci Data*. 2018;5:180142. doi:10.1038/sdata.2018.142

6. Mckhann G, Drachman D, Folstein M, Katzman R, Price D, Mckhann G. Clinical Diagnosis of Alzheimer's Disease: Report of the NINCDS-ADRDA Work Group Under the Auspices of Derpartment of Health and Human Services Task Force on Alzheimer's Disease. *Neurology*. 1984;34:27-28. doi:10.1212/01.wnl.0000400650.92875.cf

7. NIAGADS. NG00067 – ADSP Umbrella. Published 2021. Accessed November 1, 2021. https://dss.niagads.org/datasets/ng00067/

8. Bell CC. DSM-IV: Diagnostic and Statistical Manual of Mental Disorders. *JAMA*. 1994;272(10):828-829.

9. American Psychiatric Association. Diagnostic and Statistical Manual of Mental Disorders 5th edn. *American Psychiatric Association*. Published online 2013.

10. American Psychiatric Association. Diagnostic and Statistical Manual of Mental Disorders 4th edn. *American Psychiatric Association*. Published online 1994.

11. Hughes CP, Berg L, Danziger WL, Coben LA, Martin RL. A new clinical scale for the staging of dementia. *British Journal of Psychiatry*. 1982;140(6):566-572. doi:10.1192/bjp.140.6.566

12. Braak H, Braak E. Neuropathological stageing of Alzheimer-related changes. *Acta Neuropathol*. 1991;82:239-259. doi:10.1109/ICINIS.2015.10

13. Hyman BT, Trojanowski JQ. Editorial on Consensus Recommendations for the Postmortem Diagnosis of Alzheimer Disease from the National Institute on Aging and the Reagan Institute

Working Group on Diagnositic Criteria for the Neuropathological Assessment of Alzheimr Disease. *J Neuropathol Exp Neurol*. 1997;56(10):1095-1097.

14. Folstein MF, Folstein SE, McHugh PR. "Mini-Mental State" A Practical Method for Grading the Cognitive State of Patients for the Clinician. *J Psychiatr Res*. 1975;12:189-198. doi:10.3744/snak.2003.40.2.021

15. Roccaforte WH, Burke WJ, Bayer BL, Wengel SP. Validation of a Telephone Version of the Mini-Mental State Examination. *J Am Geriatr Soc*. 1992;40:697-702.

16. McKhann GM, Knopman DS, Chertkow H, et al. The diagnosis of dementia due to Alzheimer's disease: Recommendations from the National Institute on Aging- Alzheimer's Association workgroups on diagnostic guidelines for Alzheimer's disease. *Alzheimers Dement*. 2011;7(3):263-269. doi:10.1016/j.jalz.2011.03.005.

17. Albert MS, DeKosky ST, Dickson D, et al. The diagnosis of mild cognitive impairment due to Alzheimer's disease: Recommendations from the National Institute on Aging- Alzheimer's Association workgroups on diagnostic guidelines for Alzheimer's disease. *Alzheimers Dement*. 2011;7(3):270-279. doi:10.1016/j.jalz.2011.03.008.

18. Hyman BT, Phelps CH, Beach TG, et al. National Institute on Aging – Alzheimer's Association guidelines for the neuropathologic assessment of Alzheimer's disease. *Alzheimer's & Dementia*. 2012;8(1):1-13. doi:10.1016/j.jalz.2011.10.007

19. Weintraub S, Salmon D, Mercaldo N, et al. The Alzheimer's Disease Centers' Uniform Data Set (UDS) The Neuropsychologic Test Battery. *Alzheimer Dis Assoc Disord*. 2009;23(2):91-101.

20. Beekly DL, Ramos EM, Lee WW, et al. The National Alzheimer's Coordinating Center (NACC) Database: The Uniform Data Set. *Alzheimer Dis Assoc Disord*. 2007;21(3):249-258. doi:10.1097/WAD.0b013e318142774e

21. Besser LM, Kukull WA, Teylan MA, et al. The revised national Alzheimer's coordinating center's neuropathology form-available data and new analyses. *J Neuropathol Exp Neurol*. 2018;77(8):717-726. doi:10.1093/jnen/nly049

22. Mirra SS, Heyman A, McKeel D, et al. The Consortium to Establish a Registry for Alzheimer's Disease (CERAD). Part II. Standardization of the neuropathologic assessment of Alzheimer's disease. *Neurology*. 1991;41(4):479-486. doi:10.1212/WNL.41.4.479

23. Morris JC, Weintraub S, Chui HC, et al. The Uniform Data Set (UDS): Clinical and cognitive variables and descriptive data from Alzheimer disease centers. *Alzheimer Dis Assoc Disord*. 2006;20(4):210-216. doi:10.1097/01.WAD.0000213865.09806.92

24. Weintraub S, Besser L, Dodge HH, et al. Version 3 of the Alzheimer Disease Centers' Neuropsychological Test Battery in the Uniform Data Set (UDS). *Alzheimer Dis Assoc Disord*. 2018;32(1):10-17. doi:10.1097/WAD.0000000000000223

25. Bennett DA, Schneider JA, Buchman AS, De Leon CM, Bienias JL, Wilson RS. The rush memory and aging project: Study design and baseline characteristics of the study cohort. *Neuroepidemiology*. 2005;25(4):163-175. doi:10.1159/000087446

26. Bennett DA, Wilson RS, Schneider JA, et al. Natural history of mild cognitive impairment in older persons. *Neurology*. 2002;59(2):198-205. doi:10.1212/WNL.59.2.198

27. Schneider JA, Arvanitakis Z, Bang W, Bennett DA. Mixed brain pathologies account for most dementia cases in community-dwelling older persons. *Neurology*. 2007;69(24):2197-2204. doi:10.1212/01.wnl.0000271090.28148.24

28. Hinrichs AS, Karolchik D, Baertsch R, et al. The UCSC Genome Browser Database: update 2006. *Nucleic Acids Res*. 2006;34:590-598. doi:10.1093/nar/gkj144

29. Chang CC, Chow CC, Tellier LCAM, Vattikuti S, Purcell SM, Lee JJ. Second-generation PLINK: Rising to the challenge of larger and richer datasets. *Gigascience*. 2015;4(7). doi:10.1186/s13742-015-0047-8

30. Le Guen Y, Belloy ME, Napolioni V, et al. A novel age-informed approach for genetic association analysis in Alzheimer's disease. *Alzheimers Res Ther*. 2021;13:72. 10.1186/s13195-021-00808-5

31. Karczewski KJ, Francioli LC, Tiao G, et al. The mutational constraint spectrum quantified from variation in 141,456 humans. *Nature*. 2020;581(7809):434-443. doi:10.1038/s41586-020-2308-7

32. Leung YY, Valladares O, Chou YF, et al. VCPA: Genomic variant calling pipeline and data management tool for Alzheimer's Disease Sequencing Project. *Bioinformatics*. 2019;35(10):1768-1770. doi:10.1093/bioinformatics/bty894

33. GATK team. GATK Best Practices Workflows. Accessed January 31, 2021. https://gatk.broadinstitute.org/hc/en-us/articles/360035894751

34. Belloy ME, Le Guen Y, Eger SJ, Napolioni V, Greicius MD, He Z. A fast and robust strategy to remove variant-level artifacts in Alzheimer disease sequencing project data. *Neurol Genet*. 2022;8(5). doi:10.1212/NXG.0000000000200012

35. Manichaikul A, Mychaleckyj JC, Rich SS, Daly K, Sale M, Chen WM. Robust relationship inference in genome-wide association studies. *Bioinformatics*. 2010;26(22):2867-2873. doi:10.1093/BIOINFORMATICS/BTQ559

36. Chen CY, Pollack S, Hunter DJ, Hirschhorn JN, Kraft P, Price AL. Improved ancestry inference using weights from external reference panels. *Bioinformatics*. 2013;29(11):1399-1406. doi:10.1093/bioinformatics/btt144

37. Auton A, Abecasis GR, Altshuler DM, et al. A global reference for human genetic variation. *Nature*. 2015;526(7571):68-74. doi:10.1038/nature15393

38. Lambert CA, Connelly CF, Madeoy J, Qiu R, Olson M V., Akey JM. Highly punctuated patterns of population structure on the X chromosome and implications for African evolutionary history. *Am J Hum Genet*. 2010;86(1):34-44. doi:10.1016/J.AJHG.2009.12.002

39. Conomos MP, Miller MB, Thornton TA. Robust inference of population structure for ancestry prediction and correction of stratification in the presence of relatedness. *Genet Epidemiol*. 2015;39(4):276-293. doi:10.1002/gepi.21896

40. Taliun D, Harris DN, Kessler MD, et al. Sequencing of 53,831 diverse genomes from the NHLBI TOPMed Program. *Nature*. 2021;590(7845):290-299. doi:10.1038/s41586-021-03205-y

41. Das S, Forer L, Schönherr S, et al. Next-generation genotype imputation service and methods. *Nat Genet*. 2016;48(10):1284-1287. doi:10.1038/ng.3656

42. Belloy ME, Andrews SJ, Yann ;, et al. APOE Genotype and Alzheimer Disease Risk Across Age, Sex, and Population Ancestry. *JAMA Neurol*. Published online November 6, 2023. doi:10.1001/JAMANEUROL.2023.3599

43. Loh P ru, Kichaev G, Gazal S, Schoech AP, Price AL. Mixed-model association for biobank-scale datasets. *Nat Genet*. 2018;50:906-908. doi:10.1038/s41588-018-0144-6

44. Wattmo C, Londos E, Minthon L. Risk factors that affect life expectancy in alzheimer's disease: A 15-year follow-up. *Dement Geriatr Cogn Disord*. 2014;38(5-6):286-299. doi:10.1159/000362926

45. Kuzma A, Valladares O, Cweibel R, et al. NIAGADS: The NIA Genetics of Alzheimer's Disease Data Storage Site. *Alzheimer's & Dementia*. 2016;12(11):1200-1203. doi:10.1016/j.jalz.2016.08.018

46. Bycroft C, Freeman C, Petkova D, et al. The UK Biobank resource with deep phenotyping and genomic data. *Nature*. 2018;562:203-209. doi:10.1038/s41586-018-0579-z

47. Schwartzentruber J, Cooper S, Liu JZ, et al. Genome-wide meta-analysis, fine-mapping and integrative prioritization implicate new Alzheimer's disease risk genes. *Nat Genet*. 2021;53(3):392-402. doi:10.1038/s41588-020-00776-w

48. Kurki MI, Karjalainen J, Palta P, et al. FinnGen provides genetic insights from a well-phenotyped isolated population. *Nature*. 2023;613(7944):508-518. doi:10.1038/S41586-022-05473-8

49. Mbatchou J, Barnard L, Backman J, et al. Computationally efficient whole-genome regression for quantitative and binary traits. *Nature Genetics 2021 53:7*. 2021;53(7):1097-1103. doi:10.1038/s41588-021-00870-7

50. Sherva R, Zhang R, Sahelijo N, et al. African ancestry GWAS of dementia in a large military cohort identifies significant risk loci. *Mol Psychiatry*. 2023;28(3):1293-1302. doi:10.1038/s41380-022-01890-3

51. Nguyen XMT, Whitbourne SB, Li Y, et al. Data Resource Profile: Self-reported data in the Million Veteran Program: survey development and insights from the first 850 736 participants. *Int J Epidemiol*. 2023;52(1):e1-e17. doi:10.1093/IJE/DYAC133

52. Hunter-Zinck H, Shi Y, Li M, et al. Genotyping Array Design and Data Quality Control in the Million Veteran Program. *The American Journal of Human Genetics*. 2020;106(4):535-548. doi:10.1016/J.AJHG.2020.03.004

53. Fang H, Hui Q, Lynch J, et al. Harmonizing Genetic Ancestry and Self-identified Race/Ethnicity in Genome-wide Association Studies. *Am J Hum Genet*. 2019;105(4):763-772. doi:10.1016/J.AJHG.2019.08.012

54. Mägi R, Morris AP. GWAMA: Software for genome-wide association meta-analysis. *BMC Bioinformatics*. 2010;11(1):1-6. doi:10.1186/1471-2105-11-288/TABLES/2

55. Sidorenko J, Kassam I, Kemper KE, et al. The effect of X-linked dosage compensation on complex trait variation. *Nat Commun*. 2019;10(1). doi:10.1038/s41467-019-10598-y

56. Tukiainen T, Villani AC, Yen A, et al. Landscape of X chromosome inactivation across human tissues. *Nature*. 2017;550(7675):244-248. doi:10.1038/nature24265

57. Garieri M, Stamoulis G, Blanc X, et al. Extensive cellular heterogeneity of X inactivation revealed by single-cell allele-specific expression in human fibroblasts. *Proc Natl Acad Sci U S A*. 2018;115(51):13015-13020. doi:10.1073/PNAS.1806811115

58. Aguet F, Barbeira AN, Bonazzola R, et al. The GTEx Consortium atlas of genetic regulatory effects across human tissues The Genotype Tissue Expression Consortium. *Science (1979)*. 2020;369:1318-1330. doi:10.1101/787903

59. Wingo AP, Liu Y, Gerasimov ES, et al. Sex differences in brain protein expression and disease. *Nat Med*. Published online September 1, 2023. doi:10.1038/s41591-023-02509-y

60. Kerimov N, Hayhurst JD, Peikova K, et al. A compendium of uniformly processed human gene expression and splicing quantitative trait loci. *Nat Genet*. 2021;53(September):1290-1299. doi:10.1038/s41588-021-00924-w

61. Bellenguez C, Küçükali F, Jansen IE, et al. New insights into the genetic etiology of Alzheimer's disease and related dementias. *Nat Genet*. 2022;54(4):412-436. doi:10.1038/S41588-022-01024-Z

62. Giambartolomei C, Vukcevic D, Schadt EE, et al. Bayesian Test for Colocalisation between Pairs of Genetic Association Studies Using Summary Statistics. *PLoS Genet*. 2014;10(5):1-15. doi:10.1371/journal.pgen.1004383

63. Liu JZ, Erlich Y, Pickrell JK. Case-control association mapping by proxy using family history of disease. *Nat Genet*. 2017;49(3):325-331. doi:10.1038/ng.3766

64. Mohammadi P, Castel SE, Brown AA, Lappalainen T. Quantifying the regulatory effect size of cis-acting genetic variation using allelic fold change. *Genome Res*. 2017;27(11):1872-1884. doi:10.1101/gr.216747.116
